# Right Hemisphere Language Network Plasticity in Aphasia

**DOI:** 10.1101/2025.04.11.25325701

**Authors:** Peter E. Turkeltaub, Kelly C. Martin, Alycia B. Laks, Andrew T. DeMarco

**Affiliations:** Center for Brain Plasticity and Recovery, Center for Aphasia Research and Rehabilitation, Departments of Neurology and Rehabilitation Medicine, Georgetown University Medical Center, Washington, DC, USA; Research Division, MedStar National Rehabilitation Hospital, Washington, DC, USA; Department of Ophthalmology, University of Pittsburgh, Pittsburgh, PA, USA

**Keywords:** stroke recovery, neuroplasticity, brain plasticity, neurorehabilitation, remapping

## Abstract

The role of the right hemisphere in aphasia recovery has been controversial since the 19th century. Imaging studies have sometimes found increased activation in right hemisphere regions homotopic to canonical left hemisphere language regions, but these results have been questioned due to small sample sizes, unreliable imaging tasks, and task performance confounds that affect right hemisphere activation levels even in neurologically healthy adults. Several principles of right hemisphere language recruitment in aphasia have been proposed based on these studies: that the right hemisphere is recruited primarily by individuals with severe left hemisphere damage, that transcallosal disinhibition results in recruitment of right hemisphere nodes homotopic to the lesion, and that increased right hemisphere activation diminishes to baseline levels over time. It is debated whether engagement of language homotopes reflects upregulation of weakly active right hemisphere nodes in a bihemispheric language network, versus recruitment of new nodes into the network.

Here, we address these issues in 76 chronic left hemisphere stroke survivors and 69 neurologically healthy older adults using a semantic decision fMRI paradigm that elicits reliable and strongly left-lateralized individual-participant language activation and adapts to require effortful performance irrespective of participant ability levels.

Right hemisphere activation was greater in stroke survivors than controls, and related to younger age, left-handedness, and higher education. Right hemisphere activation magnitude was modest compared to left hemisphere activation. In contrast to prior proposals, right hemisphere activation was unrelated to lesion size and greater with longer time-since-stroke. Right ventral inferior frontal and mid-anterior temporal nodes were weakly engaged in language processing in controls and co-activated with their homotopic left hemisphere counterparts. Lesions to those left hemisphere counterparts resulted in increased homotopic activation that contributed to naming and word reading outcomes. In contrast, the right dorsal inferior frontal cortex was not engaged during language processing in controls and did not co-activate with its left hemisphere counterpart. After stroke, it exhibited the largest increase in group-level activation due to complex lesion-activation interactions, but the activation was unrelated to aphasia outcomes.

In sum, right hemisphere language homotopes are recruited in the chronic phase of aphasia recovery, consistent with both upregulation of weakly active nodes in a bihemispheric language network and recruitment of the dorsal inferior frontal gyrus as a new node. These findings clarify the mechanisms of, and constraints on, right hemisphere language network plasticity in adults and may guide selection of candidates likely to benefit from neuromodulatory aphasia treatments.

## Introduction

Left hemisphere (LH) strokes often disrupt language network function, causing aphasia. Recovery from aphasia is thought to rely in part on resolution of this dysfunction, reflected by partial normalization of LH language network activity.^1–3^ Right hemisphere (RH) regions homotopic to canonical LH language nodes (hereafter, the “RH Language Network”) may also be recruited by people with aphasia.^4^ The role of the RH Language Network in aphasia recovery has been debated since the 19th century and remains controversial.^5,6^

Evidence of RH involvement in aphasia recovery began with Barlow’s 1877 case of a boy who became aphasic after a stroke of the left inferior frontal gyrus (IFG), then recovered, but worsened again after a stroke of the right IFG.^7^ The validity of this case has been questioned,^8,9^ but later cases of sequential LH and RH strokes in adults reported similar findings.^10,11^ Other lines of evidence have also suggested RH compensation for aphasia, for example, worsening of aphasia after right carotid anesthesia.^12^ These findings suggest that, overall, the RH can contribute to language ability after damage to the LH language network, although they lack the anatomical precision needed to implicate specific RH structures.

Functional neuroimaging has provided that anatomical precision spurred an explosion of aphasia recovery research.^4^ An early meta-analysis of this literature found that chronic LH stroke survivors consistently activated RH language homotopes whereas controls did not, but did not compare the groups directly.^13^ A subsequent meta-analysis found greater activity in some RH Language Network regions compared to controls.^14^ A third meta-analysis found evidence for increased activity in stroke survivors with aphasia compared to controls exclusively in the right IFG, but questioned the reliability of this finding due to methodological limitations of the studies contributing to it.^15^ Indeed, imaging studies of aphasia have often been plagued by task performance confounds. People with aphasia have difficulty performing language tasks, and RH language activation can increase during challenging language processing.^16,17^ As a result, increased RH activation may simply reflect poor in-scanner task performance by people with aphasia rather than neuroplastic changes in language processing. Prior studies have also generally used small sample sizes and fMRI tasks that do not produce reliable activation patterns in individual people.^18^ These two issues are critical because RH Language Network recruitment may vary from person to person based on the size and location of the stroke and also demographic factors related to pre-morbid network organization.^19^

Whether RH recruitment helps or hinders aphasia recovery also remains debated. As noted above, several older lines of evidence suggest the RH as a whole contributes to aphasia recovery. Some neuroimaging findings also support this conclusion for specific RH language nodes.^20–22^ In contrast, inhibiting the right IFG using repetitive transcranial magnetic stimulation (rTMS) improves language outcomes in people with aphasia.^23^ Prominent frameworks of aphasia recovery have suggested the RH is ineffective or even maladaptive in aphasia recovery.^24,25^ We have previously argued that individual nodes of the RH Language Network may be recruited based on different neuroplastic mechanisms and contribute differentially to recovery.^5,6,11,13,19^

Hypotheses regarding neuroplastic mechanisms of RH recruitment must consider the status of the RH Language Network in typical adults and its developmental origins. At birth, the two hemispheres are thought to be equipotential for language,^26^ albeit with a LH bias for speech and language processing.^27^ When a large LH stroke occurs around the time of birth, children can successfully acquire language using the RH Language Network.^28,29^ In typical development, RH language activity diminishes over the course of childhood,^30^ but a developmental remnant, recently dubbed the “weak shadow” of early life RH language, remains relatively unchanged in its localization when subthreshold activation is examined.^31^ Some of these weakly active RH Language Network nodes are recruited in relation to effort required for language processing.^16,17^ This recruitment likely reflects their role as nodes in a bihemispheric, but left-lateralized, language network.^32,33^ The RH weak shadow is not a perfect mirror of the LH language network^29^ and so some RH regions homotopic to LH language nodes may not be part of the bihemispheric language network.

Thus, we arrive at two hypotheses regarding RH language in aphasia recovery. The first is that recruitment of homotopic RH language nodes reflects increased use of homotopic processors that are already weakly engaged in language prior to stroke (i.e., the RH “weak shadow”). This represents a modest recalibration of weights in residual network nodes, potentially through modification of existing synapses.^3,32^ Some stroke survivors may therefore recruit homotopic RH nodes more successfully than others because their language networks were less strongly left-lateralized before the stroke.^34^ This might, for example, relate to age or left-handedness.^35^ The second hypothesis is that changes in RH activation occur in regions that are homotopic to LH language nodes, but not engaged, even weakly, in language processing prior to stroke. This would suggest a more dramatic form of neuroplastic reorganization, sometimes called remapping or vicariation.^6,36–38^ This might occur, for example, due to competition for shared axonal projection targets or a release of inhibition on nodes of a different network. Testing these hypotheses requires a systematic examination of the factors driving recruitment of individual RH language nodes and their relationships with aphasia outcomes. These factors may include personal factors, like age and handedness as discussed above, or lesion factors, which we will discuss below.

Lesion location is a key factor that may drive RH Language Network recruitment. For example, the interhemispheric inhibition hypothesis argues for lesion-dependent recruitment of RH nodes.^39^ Under this proposal, homotopic LH and RH processors are said to inhibit each other via transcallosal fibers, achieving a balance in which the dominant LH suppresses the RH. A LH lesion is thought to release homotopic RH processors from inhibition, resulting in increased activation. Some studies have indeed found that recruitment of right IFG or right ventral motor regions relates to homotopic left frontal lesions.^13,20,40^ However, posterior RH language nodes do not seem to follow this pattern.^2,21^ Further, the interhemispheric inhibition hypothesis predicts that RH language activity should relate not only to homotopic LH lesions, but also to deactivation of those lesioned nodes. This prediction has never been systematically tested, to our knowledge.

Overall lesion size may also determine RH recruitment. Individuals with large LH strokes are often said to rely more on the RH Language Network than those with small strokes,^3,24,25,39^ although direct evidence for this claim is lacking. If RH Language Network recruitment reflects reweighting of nodes in a bihemispheric network, only lesions large enough to severely disrupt LH processing may induce network reweighting toward the spared RH.^3,33^ One prior study did find that larger lesions related to greater RH activation during picture naming, but the pattern of findings suggested that in-scanner effort due to severe anomia drove the effect, not true reorganization of the language network.^20^

RH recruitment also varies over time after stroke. Longitudinal studies have suggested that RH language engagement peaks a few weeks after stroke and then normalizes over time, returning to neurotypical levels by six months.^1,2^ These effects too could be driven by effort for fMRI task performance as aphasia improves over time. Further, mechanisms of RH plasticity may differ longer after stroke, and prior studies have not examined the relationship of RH activation to time-since-stroke during the chronic phase of recovery.

Here, we aim to address the key questions raised above about RH language recruitment in aphasia, using a large sample of stroke survivors and controls, and an fMRI task that produces reliable single-participant activation maps and adapts to require effortful performance irrespective of ability level.^18,41^ Our results provide clear evidence for contributions of some RH Language Network nodes to aphasia recovery and identify personal and lesion factors related RH recruitment. Different neuroplastic mechanisms appear to account for recruitment of different RH nodes, with evidence both for increased reliance on weakly active RH language nodes, and recruitment of homotopic RH nodes not previously engaged in language processing, at least as measured by our fMRI task. These results may serve as the basis for individualized approaches to optimize language network organization to improve aphasia recovery.

## Materials and methods

### Participants

Participants were recruited for a cross-sectional study of language outcomes after LH stroke (clinicaltrials.gov NCT04991519). From an initial sample of 103 stroke survivors and 70 neurologically healthy controls, 76 participants with LH stroke and history of aphasia, and 69 age-matched controls were included in the final analyses (Table 1). Participants were native English speakers without stroke outside the LH or history of significant learning disability, psychiatric or other central neurological conditions. See Supplementary Materials for details. Study procedures were approved by the Georgetown University IRB and participants’ consent was obtained according to the Declaration of Helsinki.

**Table 1.** Participant Characteristics.

|  | Stroke | In-Sample Controls | Held-Out Controls used to define Language Network | Stroke vs. In-Sample Controls |
| --- | --- | --- | --- | --- |
| Sample Size | 76 | 49 | 20 |  |
| Age (years) | 60.2 (11.6) | 60.4 (10.7) | 60.9 (13.3) | $t_{(123)} = 0.11, P = .91$ |
| Gender (M:F) | 43:33 | 23:26 | 10:10 | $\chi^2_{(2)} = 0.94, P = .63$ |
| Race (Black:White:More-Than-One) | 30:45:1 | 22:27:0 | 1:19:0 | $\chi^2_{(2)} = 0.13, P = .72$ |
| Ethnicity (Hispanic:Not-Hispanic) | 5:71 | 0:49 | 0:20 | $\chi^2_{(2)} = 1.86, P = .17$ |
| Education (years by highest degree) | 16.3 (2.9) | 17.1 (2.7) | 17.4 (2.1) | $t_{(123)} = 1.64, P = .10$ |
| Handedness Index | 69.6 (53.3) | 70.8 (41.4) | 69.4 (55.3) | $U_{(123)} = 1596, P = .17$ |
| Handedness (Left:Ambi:Right) | 9:0:67 | 3:1:45 | 2:0:18 | $\chi^2_{(2)} = 2.61, P = .27$ |
| Time-Since-Stroke (months) | 44.4 (52.2) |  |  |  |
| Lesion Size (cubic cm) | 89.5 (88.6) |  |  |  |
| WAB Spontaneous Speech (/20) | 15.7 (4.3) |  |  |  |
| WAB Naming and Word Finding (/10) | 7.9 (2.2) |  |  |  |
| WAB Repetition (/10) | 7.7 (2.4) |  |  |  |
| WAB Auditory-Verbal Comprehension (/10) | 8.9 (1.3) |  |  |  |
| Oral Word Reading (%correct) | 71.7 (29.6) |  |  |  |
| Semantic Judgment Composite (%correct) | 88.9 (7.6) |  |  |  |

Values are Mean and (standard deviation). Categorical handedness is determined by Edinburgh Handedness Index > or < +/-20.

### MRI acquisition

Participants were scanned on a Siemens 3T Prisma Fit with 20-channel head coil, including a high-resolution T1-weighted scan (MPRAGE), a T2-weighted fluid-attenuated inversion recovery (FLAIR) scan, and a BOLD T2*-weighted functional MRI scan.

See the Supplementary Materials for details of MRI acquisition and analyses described below.

### Lesion delineation

Lesions were manually segmented on each participant’s MPRAGE and FLAIR images by author P.E.T. in ITK-SNAP. Lesion masks were warped to the Clinical Toolbox Older Adult Template using Advanced Normalization Tools^42^ with a custom pipeline.^43^ A lesion overlap map is shown in Supplementary Fig. 1.

### fMRI methods

Language activation was measured used an adaptive semantic decision task validated in people with aphasia.^41^ Participants viewed word pairs and indicated via button press if they were related in meaning (e.g., shark–whale). During a control condition, participants indicated via button press if pseudofont pairs (e.g., LDLLD–LLLLD) were identical. A staircase procedure adjusted the difficulty of each task based on accuracy.^41^ Fifteen stroke participants from the original sample were excluded for inability to perform the semantic decision task (binomial *P* < 0.05). Standard preprocessing was performed in AFNI.^44^ A whole-brain general linear model assessed the contrast of semantic decision vs. pseudofont matching. Resulting single-subject statistical maps were warped to MNI space.

### Defining networks and nodes

To identify RH homotopes to canonical language regions, we first mapped the active LH regions in 20 held-out control participants using a voxelwise one-sample t-test *(P <* 0.0001, clusters > 200 voxels). We then employed a procedure to break up clusters into individual subregions (hereafter “nodes”) surrounding the peaks of activity. To focus analyses on RH homotopes of canonical LH language regions, we grouped lateral fronto-temporo-parietal nodes as the “Language Network” and other nodes as the “Semantic/Executive Network.” Nodes were labeled by hemisphere (R, L), network (Lang, Sem), and an arbitrary number (e.g., RLang05).

Lesion load was calculated for each stroke participant for each LH node as the proportion of voxels in the node that intersected with the participant’s lesion mask. fMRI activation for each participant for each node was computed as the mean t-value of all voxels in the node. Analyses comparing Stroke participants to Control participants used these nodewise mean t-values. Analyses exclusively examining the Stroke group used these nodewise mean t-values Z-scored against the in-sample control group (i.e., excluding the 20 held-out controls) to interpret results in terms of nodewise over- vs. under-activation compared to controls.

### Behavioral measures

Participants completed a behavioral battery for the larger project. Here, we use the four subscores from the Western Aphasia Battery-Revised^45^ (WAB-R; Spontaneous Speech, Naming and Word Finding, Repetition, and Auditory-Verbal Comprehension), and two measures relevant to the fMRI task demands: oral single-word reading accuracy, and a composite semantic measure combining accuracies on a shortened Pyramids and Palm Trees test^46^ and a picture-based category judgment task.^47^

### Statistical analyses

Statistical tests were implemented in JASP (Version 0.19.2)^48^ (https://jasp-stats.org/). Linear mixed-effects models used the maximal random effects structure justified by the design.^49^ Random slopes were removed to retain the maximal random effects structure that did not result in singular fits. Demographic covariates included in all analyses were age, education (years by degree), gender, and handedness (Edinburgh Handedness Inventory Handedness Index^50^). The specific models are described in the Results section, with model formulae in the Supplementary Tables.

### Lesion-symptom mapping

To identify lesion locations associated with increased RH activity, we conducted voxelwise support vector regression lesion-symptom mapping (SVR-LSM)^51^ using the SVR-LSM MATLAB toolbox.^52^ The dependent measures were Z-scored nodewise activation for RH Language Network nodes. Lesion volume, age, education, handedness, log-transformed time-since-stroke, gender, homotopic LH node activation, and semantic decision accuracy were regressed out of the lesion and behavioral data.^52^ Analyses examined voxels lesioned in at least 10% of participants. Significance was tested using 5000 permutations (voxelwise *P* < 0.005, family-wise error cluster *P* < 0.05).

### Data availability

Data will be made available upon reasonable request to the authors.

## Results

### The Language Network

The group fMRI map from the 20 held-out controls (Fig. 1a) included 25 nodes that were mirrored to the RH to define the RH homotopic networks. Of the 25 nodes, 11 were grouped as the Language Network (Fig. 1b), and 14 were grouped as the Semantic/Executive Network based on their location in structures typically associated with semantic cognition and/or “domain general” executive functions (Supplementary Fig. 2).

**Figure 1.**
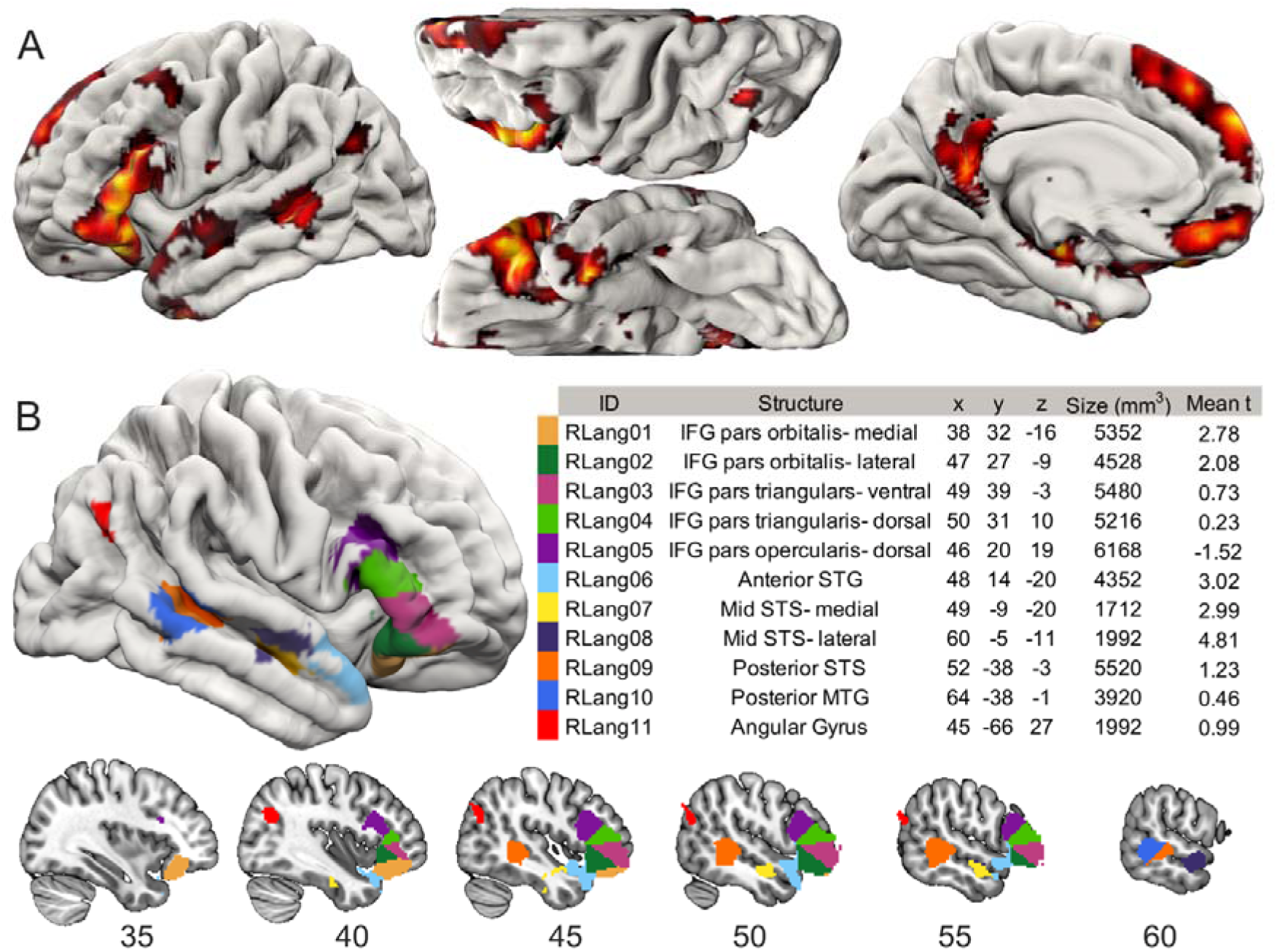
The language network. (**A**) The group fMRI map from the 20 held-out controls that was used to define the Language and Semantic/Executive Networks, voxelwise *P* < 0.0001. (**B**) The homotopic RH Language Network derived by clustering the fMRI group map and flipping over the midsagittal plane. The x y z coordinates are in MNI space. “Mean t” refers to the mean group-level activation (mean of voxelwise t-scores in the group map) in the node in the held-out controls. IFG: inferior frontal gyrus, STG: superior temporal gyrus, STS: superior temporal sulcus, MTG: middle temporal gyrus.

Activation levels of homotopic RH Language Network nodes ranged in the held-out controls (Fig. 1b). The IFG pars orbitalis (RLang01, RLang02), the anterior superior temporal gyrus (STG; RLang06) and the medial mid-superior temporal sulcus (STS; RLang07) were weakly activated (mean *t* = 2 to 3). The lateral mid-STS (RLang08) was activated above the voxelwise threshold used to define the LH Language Network (mean *t* = 4.81). The dorsal IFG pars opercularis (RLang05) was marginally deactivated (semantic decision < pseudofont control, mean *t* = −1.51).

### fMRI task performance

We next asked whether the adaptive semantic decision task achieved equivalent in-scanner performance across participants (Supplementary Fig. 3). As expected, control participants achieved a higher difficulty level than stroke survivors on the semantic decision task (mean level 5.3 vs. 3.8, *t*(122.6) = 7.29, *P* < 0.001), but not on the pseudofont control task (mean level 4.7 vs. 4.5, *t*(123) = 1.22, *P* = 0.23). Despite the adaptive procedure, semantic decision accuracy was higher in controls than stroke participants (86.9% vs. 80.7%, *t*(122.9) = 7.00, *P* < 0.001), and was inversely correlated with lesion size (*rho* = −0.245, *P* = 0.03). Accuracy on the pseudofont control condition was not different between groups (84.6% vs. 84.0%, *t*(123) = 0.96, *P* = 0.18), and was unrelated to lesion size (*rho* = −0.161, *P* = 0.17). To ameliorate residual performance confounds, we controlled for semantic decision accuracy in subsequent analyses.

### Does RH Language Network activity increase after LH stroke?

We tested whether overall activation of the homotopic RH Language Network increases after LH stroke using a linear mixed-effects model examining the fixed effect of Group (Stroke vs. Control) on nodewise activity, covarying for demographic variables and semantic decision accuracy. The effect of Group was significant (*F*(1,118) = 4.44, *P* = 0.037; Fig. 2; Supplementary Table 1), with Stroke activation greater than Controls (estimated marginal mean activity: Stroke = 0.856, vs. zero *P* < 0.001; Control = 0.388, vs. zero *P* = 0.066). Higher RH language activation was also related to younger age *(P =* 0.005), greater education *(P =* 0.005), and left-handedness (*P* < 0.001). Adding interactions of these variables with Group revealed no significant effects (all *P* > 0.10). The distribution of overall mean activation revealed that 14 LH stroke participants had greater RH Language Network activity than every control participant except two right-lateralized controls (RH > LH overall activity).

**Figure 2.**
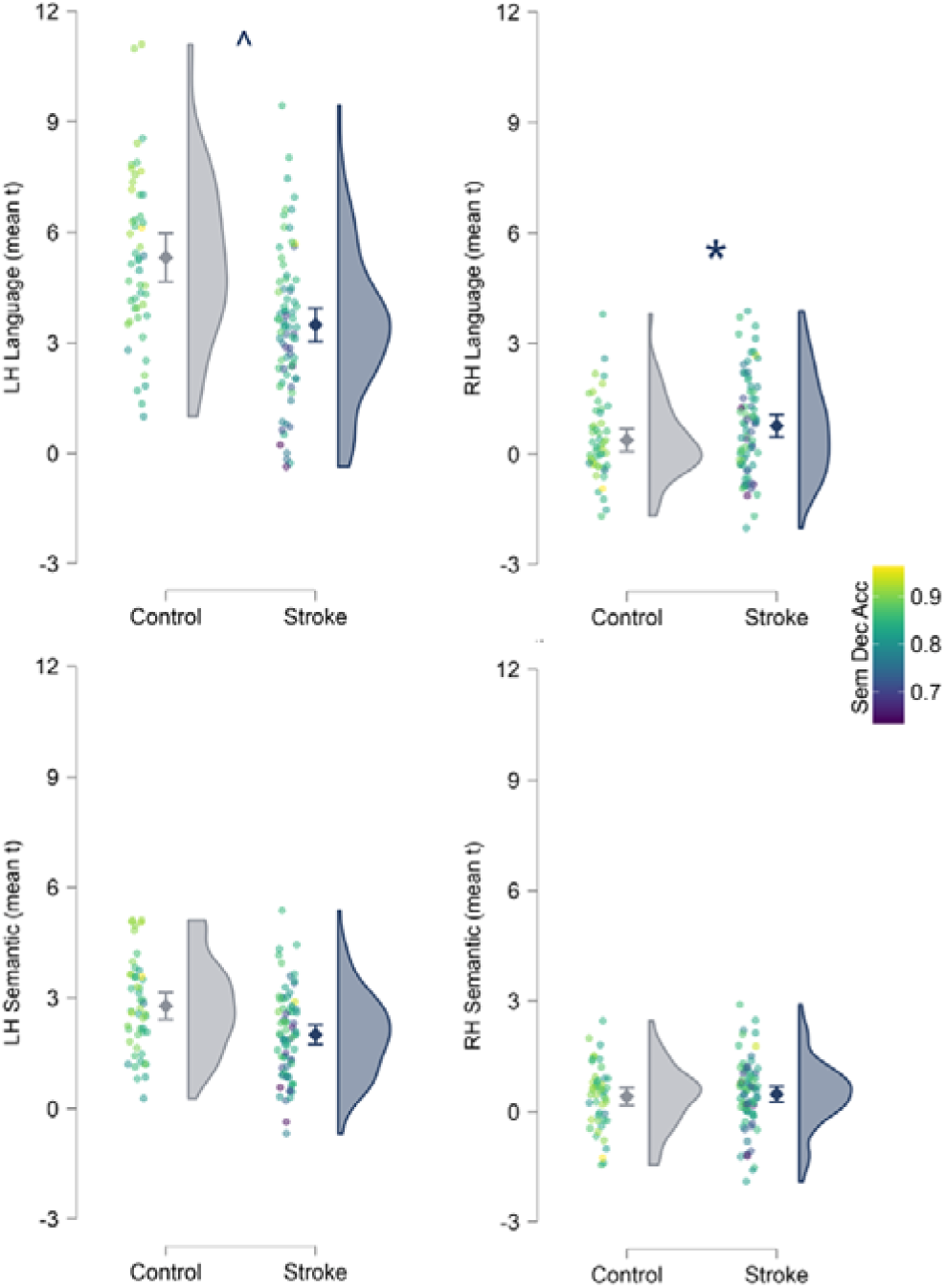
Overall activation of the LH and RH Language and Semantic/Executive Networks in Stroke and Control groups. Raw mean t-values for the entire network in each hemisphere are shown without accounting for covariates. The diamond is the mean and error bars are the 95% confidence interval. \**P* < 0.05, ^*P* < 0.10

For comparison, we performed similar linear mixed-effects models examining activation of the LH Language Network and the LH and RH Semantic/Executive Networks (Supplementary Tables 2-4). Distribution plots showed that the magnitude of RH language activity in the stroke group was modest compared to LH language activity (Fig. 2). No between-group differences were found in these three networks (LH Language: *F*(1,118) = 2.96, *P* = 0.088; LH Semantic/Executive *F*(1,118) = 1.16, *P* = 0.285; RH Semantic/Executive *F*(1,118) = 2.01, *P* = 0.159), but the distribution of raw activation values in both LH networks appeared reduced in stroke participants. The effect of semantic decision accuracy on activity was strong in both of these networks (LH Language: *F*(1,118) = 29.14, *P* < 0.001; LH Semantic/Executive: *F*(1,118) = 18.40, *P* < 0.001), suggesting that apparently lower activation in stroke participants related primarily to poor fMRI task performance.

Next, we used ANCOVAs to test for group activation differences in each of the 11 RH language nodes covarying for the same predictors as above (Supplementary Tables 5-15). Only activation of the dorsal pars triangularis (RLang04) was different between the groups after multiple-comparisons correction (Stroke > Control, *F*(1,118) = 8.37, *P* = 0.0045, Fig. 3). The dorsal IFG pars opercularis (RLang05) showed a group difference at an uncorrected threshold (Stroke > Control, *F*(1,118) = 5.85, *P* = 0.017), an effect that was larger without controlling for semantic decision accuracy (*F*(1,118) = 11.91, *P* < 0.001). Notably, neither RLang04 or RLang05 were activated even weakly by the control group. Control group activation of RLang04 was near zero (mean *t*-score= 0.2, single group t-test vs. zero: *t*(48) = 0.07, *P* = 0.94), and RLang05 was deactivated relative to the symbol matching task in controls (mean *t*-score = −0.94, single group t-test vs. zero: *t*(48) = −2.76, *P* = 0.008). No other node exhibited a group difference in activity at an uncorrected *P* < 0.05, although there were trending effects and every node showed numerically greater marginal mean activity in the stroke group than controls. For comparison, we ran ANCOVAs on the RH semantic/executive network nodes, which revealed no group differences (all *uncorrected P* > 0.10, Supplementary Tables 16-29).

**Figure 3.**
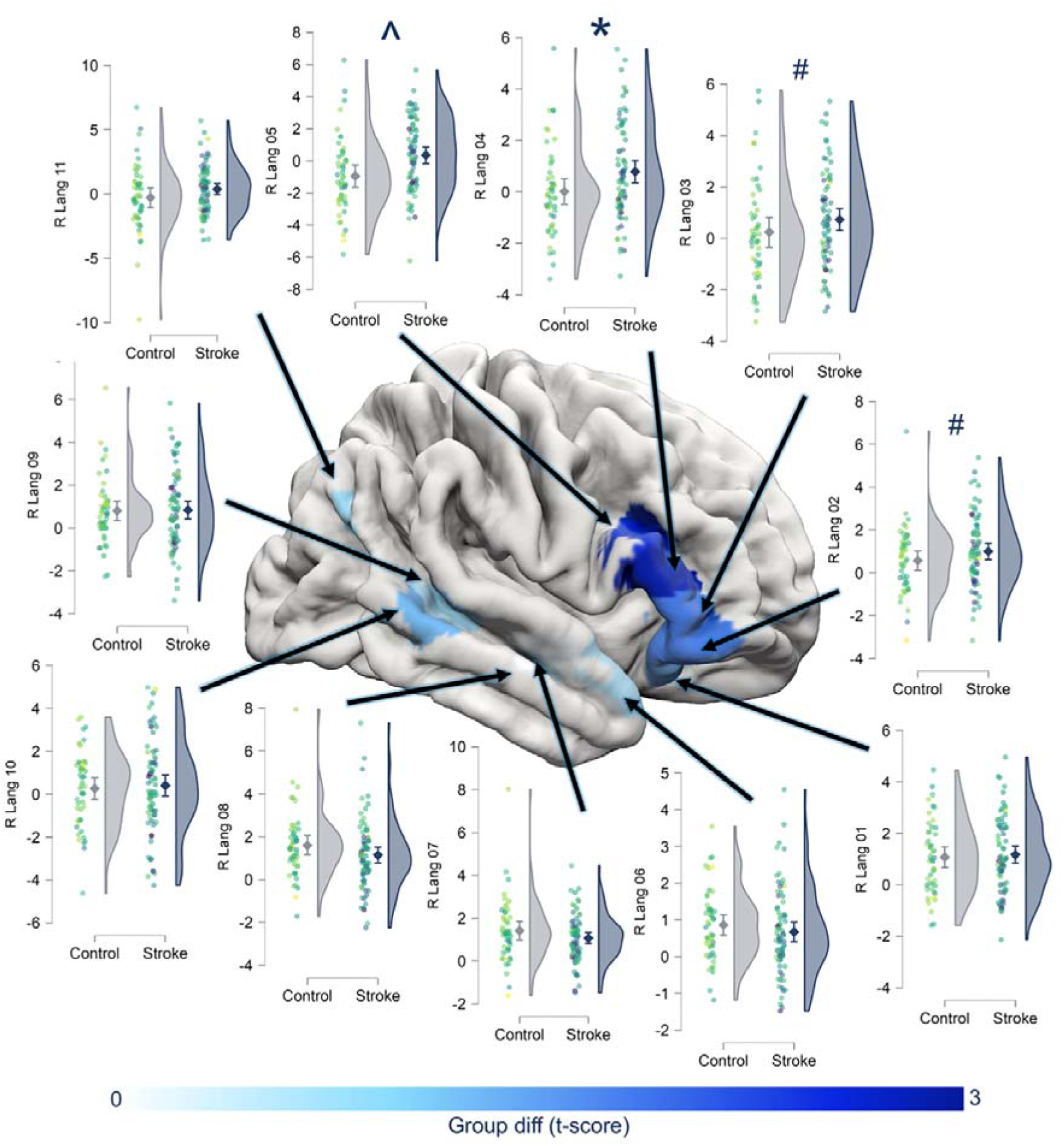
Group differences in activation at each RH Language Network node. The brain rendering depicts the t-score of the effect of group in the regressions examining activity of the 11 RH Language Network nodes. Plots show raw activation values not accounting for covariates, with individual participant points colored based on semantic decision accuracy. \**P* < 0.05 corrected for 11 comparisons, ^*P* < 0.05 uncorrected, #*P* < 0.10 uncorrected

### Is RH activity related to larger lesions and earlier time-since stroke?

To test if RH recruitment occurs primarily in larger strokes and normalizes over time, we used a linear mixed-effects model examining fixed effects of log-transformed lesion size and log-transformed time-since-stroke on nodewise RH language activity in the stroke group, including the same covariates as the models above (Supplementary Fig. 4; Supplementary Table 30). RH language activity was related to time-since-stroke (*F*(1,64.4) = 4.45, *P* = 0.039), but not lesion size (*F*(1,68) = 0.04, *P* = 0.85). In contrast to the predicted direction of effect, RH activity increased rather than decreased with greater time-since-stroke. RH language activity was also greater with younger age (*P* = 0.006), higher education (*P* = 0.035), and left-handedness (*P* = 0.017).

### Does transcallosal disinhibition account for RH engagement?

Next, we addressed the hypothesis that damage or dysfunction in a LH language node causes a loss of transcallosal left-to-right inhibition, resulting in increased activation of the homotopic RH node. To test this, we added fixed effects of homotopic LH nodewise lesion load and activation to the mixed-effects model above (Supplementary Table 31). This analysis revealed the predicted effect of nodewise LH lesion load on homotopic RH nodewise activity (*F*(1,13.4) = 21.39, *P* < 0.001). However, in contrast to the prediction that reduced LH activation drives homotopic RH recruitment, activity in LH nodes and homotopic RH nodes were positively related (*F*(1,44.2) = 42.64, *P* < 0.001). This result seemed paradoxical so we next confirmed that lesions reduce activation of LH nodes using another linear mixed-effects model examining the fixed effect of nodewise lesion load on nodewise LH activity.

This confirmed that lesions reduce activation at the lesioned LH node (*F*(1,15.5) = 31.20, *P* < 0.001, Supplementary Table 32). To confirm that covariance between nodewise LH lesion load and activity did not cause the seemingly paradoxical effects on homotopic RH activity, we ran two additional models including only one of the predictors at a time, which confirmed both effects independently (LH lesion load: *F*(1,44.9) = 5.19, *P* = 0.028; LH activity: *F*(1,29.9) = 16.74, *P* < 0.001; Supplementary Tables 33 and 34).

To localize the effects, we ran a series of linear regressions examining activation of each RH language node using the same predictors as above (Fig. 4, Supplementary Tables 35-45). Activity of the right lateral mid-STS (RLang08) related both to lesions and activation of its LH homotope after correction for 22 comparisons (two variables x 11 nodes). Effects of homotopic LH lesion load (Fig. 4a) and activation (Fig. 4c) were strongest in the mid-anterior temporal and ventral IFG nodes. We reasoned that the relationship between LH and RH activity might reflect continued engagement of RH nodes that were already weakly engaged in language before the stroke. Indeed, the nodewise effects of homotopic LH lesions and activity were related to the nodewise activation levels in controls (correlation between nodewise mean t-score in controls and effect of LH homotopic lesion load: *r* = 0.740, *P* = 0.009, LH homotopic activation: *r* = 0.738, *P* = 0.010; Fig. 4b). Another mixed effects model confirmed that RH nodes tend to coactivate with their homotopic LH counterparts in control participants (*F*(1,78.9) = 15.98, *P* < 0.001, Supplementary Table 46). Plots of nodewise LH activity vs. RH activity in controls showed that relationships between LH and RH activity were strongest in the mid-anterior temporal nodes (RLang06, RLang07, RLang08). Adding the stroke group to these plots, stratified by lesion status at each LH node, demonstrated that lesions to LH nodes resulted in reduced LH activation and increased RH activation in many nodes (Fig. 4c). Further, for most RH nodes, the coactivation relationships between the LH and RH appeared similar between stroke survivors and controls when the LH node was spared by the lesion. In the mid-anterior temporal lobe (RLang06, RLang07, RLang08), this coactivation relationship remained intact even when the LH node was lesioned. In sum, in RH nodes that are already engaged to some degree in the language network in controls, lesions that damage their homotopic LH counterparts result in increased activation without affecting their prior relationship to the network.

**Figure 4.**
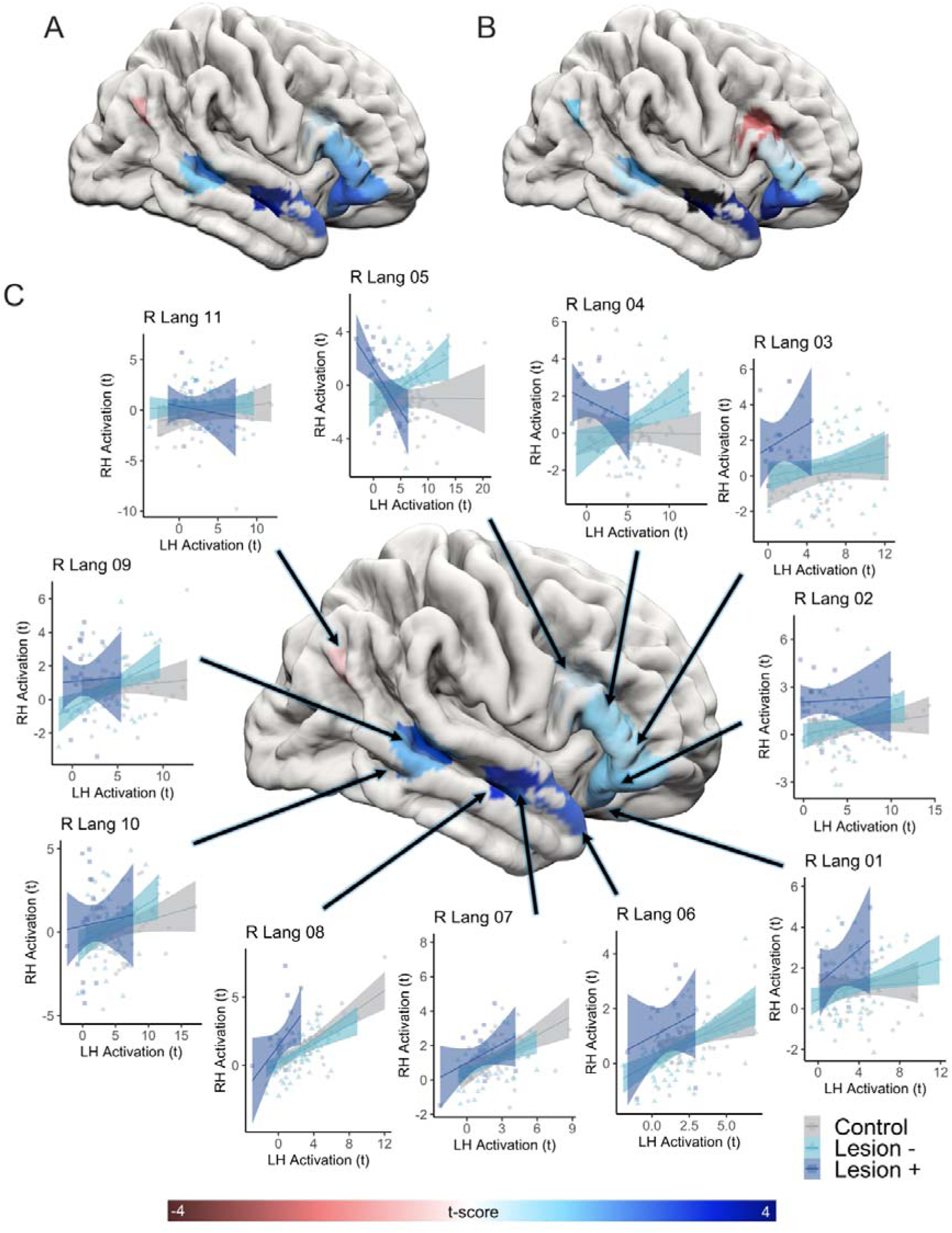
Effects of homotopic LH lesions and activation on nodewise RH activity. (**A**) Nodewise effects of homotopic LH lesions on RH activation are shown with darker blue indicating greater effects and darker red indicating greater inverse effects. (**B**) Nodewise activation levels in controls are shown to demonstrate that nodewise effects of homotopic LH lesions and activation are related to activation levels in controls, indicating that the RH language nodes that increase in activation in relation to homotopic lesions and activation tend to be those that were engaged in the language network to some degree prior to stroke. Darker blue indicates greater activation in controls, and darker red indicates greater deactivation in controls. (**C**) Nodewise effects of homotopic LH activation are shown on the rendered hemisphere using the same color scale used in **A**. Surrounding the rendered hemisphere are plots showing the relationship between homotopic LH and RH activation at each node in three groups: Controls (gray), Stroke survivors without lesions involving the homotopic LH node (light blue), and Stroke survivors with lesions involving the homotopic LH node (dark blue). LH nodes were categorized as lesioned if >=25% of the voxels in the node were in the participant’s lesion mask. These plots illustrate that: LH activation tends to decrease in many nodes in stroke survivors, particularly when the node itself is lesioned; LH and RH activation are correlated in all groups in many nodes, particularly those that activated to some degree in controls (compare to panel **B**); lesions to the homotopic LH node results in a shift upward in RH activation but does not substantially change the relationship between LH and RH activation in most nodes; in the dorsal IFG nodes (RLang04 and especially RLang05) behave differently than all other nodes in that LH and RH activation are unrelated in controls, directly related in stroke survivors with lesions elsewhere in the LH, and inversely related when lesions affect the corresponding LH node. The interaction of homotopic LH lesions and homotopic LH activation on RH activation is only significant for RLang05.

The lesion-activation plots revealed a different pattern in the right dorsal IFG nodes (Fig. 4c, RLang04 and RLang05). Activity of these nodes was unrelated to their homotopic LH counterparts in control participants, but related in the stroke group. When the left dorsal IFG nodes were unlesioned, the left and right dorsal IFG nodes co-activated (i.e., a positive relationship between homotopic LH and RH activity). In contrast, when the left dorsal IFG nodes were lesioned, activation was inversely related, such that right dorsal IFG activity was greatest in individuals who did not activate the left dorsal IFG. Adding an interaction term to the regressions confirmed a significant interaction between LH lesion and LH activity for dorsal pars opercularis (RLang05 *P* = 0.013, Supplementary Table 39), but not dorsal pars triangulars (RLang04 *P* = 0.53, Supplementary Table 38). This pattern of findings is partly compatible with the hypothesis that loss of transcallosal left-to-right inhibition drives recruitment of right dorsal IFG.

### Do lesions to specific LH structures drive RH activation?

We then used SVR-LSM analyses to identify lesions associated with greater activity in the RH language nodes, homotopic or not. This revealed two distinct lesion-activation relationships (Fig. 5, slices shown in Supplementary Fig. 5, Supplementary Table 47). First, lesions in the left IFG and anterior insula related to greater activity in the right ventral IFG (RLang01, RLang02, RLang03) and also the posterior STS (RLang09) and middle temporal gyrus (MTG; RLang10). Second, lesions of the left anterior and mid-temporal lobe related to greater activity of the right anterior and mid-temporal nodes (RLang06, RLang07, RLang08). No lesion-activation relationships were observed for the right dorsal IFG or the angular gyrus (RLang04, RLang05, RLang11, all cluster *P* > 0.10).

**Figure 5.**
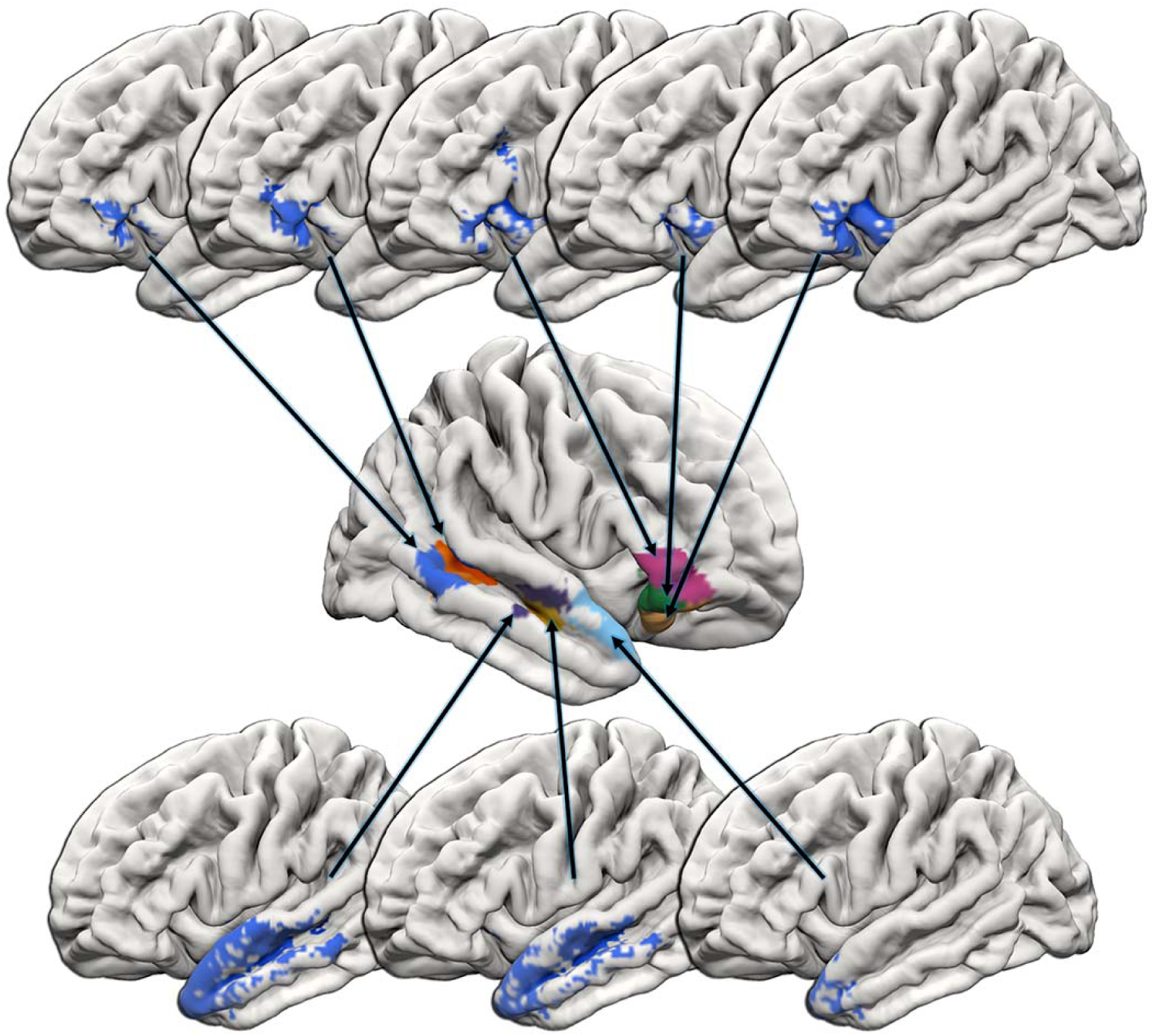
Lesions related to greater RH language activity. Significant SVR-LSM results for the individual RH language nodes. Arrows connect the SVR-LSM maps to the RH language nodes where activity is increased by the lesions. The top row shows results for the nodes where RH activation increases with left inferior frontal lesions - left to right: RLang10, RLang09, RLang03, RLang02, RLang01. The bottom row shows results for the nodes where RH activation relates to anterior temporal lobe lesions-left to right: RLang08, RLang07, RLang06. All analyses control for lesion size, age, education, handedness, gender, log-time-since-stroke, semantic decision accuracy, and LH activity in the homotopic LH language nodes. Maps are thresholded at voxelwise *P* < 0.005, cluster corrected *P* < 0.05.

### Does RH language activation contribute to aphasia outcomes?

Next, we tested if nodewise RH activation predicted aphasia outcomes. Identifying the contribution of RH activation to outcomes requires accounting for demographic and lesion factors that might contribute to aphasia outcomes. Therefore, we used nonparametric partial correlations controlling for age, education, handedness, gender, log-time-since-stroke, log-lesion size, and lesion load in each LH language node and the three LH semantic nodes with adequate lesion coverage (>25% lesion load in at least 10% of participants). Even after accounting for these other predictors, activity of the lateral mid-STS (RLang08) was related to oral word reading (*ρ* = 0.448, *P* = 0.0006) after correction for 66 comparisons (11 nodes x 6 behaviors; Fig. 6; Supplementary Table 48). Correcting only for 11 nodes, but not 6 behaviors, activity of the right medial IFG pars orbitalis (RLang01) related to WAB-R Naming and Word Finding. Several relationships were observed at uncorrected thresholds, including RLang01 and RLang10 (posterior MTG) with oral reading, RLang01 with semantic judgment, and RLang08 with Naming and Word Finding (all *P* < 0.01, uncorrected).

**Figure 6.**
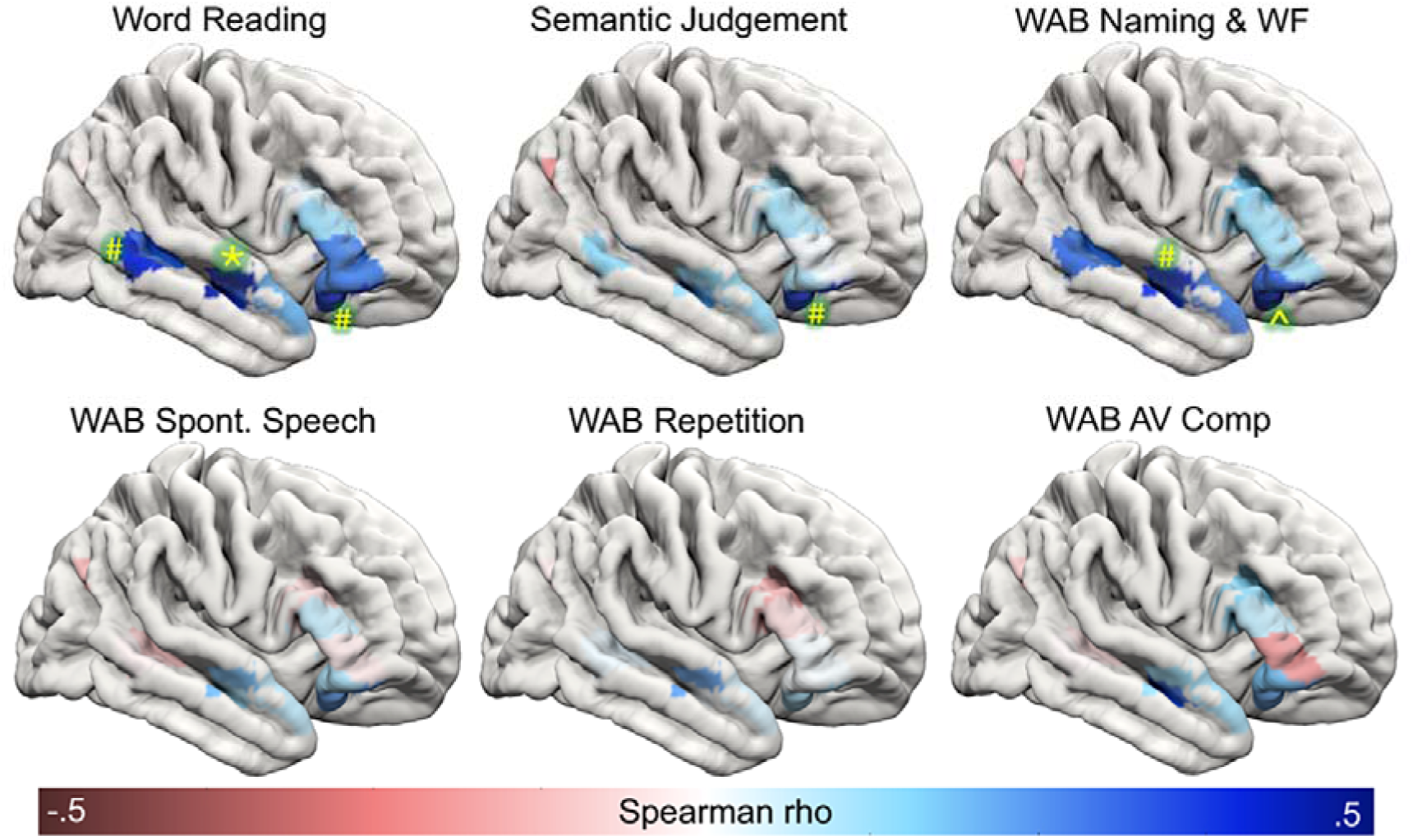
Relationships between RH language activity and aphasia outcomes. The color scale shows the effects at each RH Language Network node based on nonparametric partial correlations between activity and the behavioral score, controlling for log-transformed lesion size, lesion load in LH nodes with adequate lesion coverage (>25% lesion load in at least 10% of stroke participants; 14 nodes in total: all 11 LH Language Network nodes plus LSem04, LSem05, LSem12), log-transformed time-since-stroke, age, education, gender, and handedness. * *P* < 0.05 corrected for 66 comparisons, ^*P* < 0.05 corrected for 11 comparisons, #*P* < 0.01 uncorrected

For comparison, we used similar correlations to examine behavioral relationships for the LH Language Network and LH and RH Semantic/Executive network nodes (Supplementary Table 49). As expected, activity of LH Language Network nodes showed the strongest behavioral relationships overall, with several significant relationships surviving full correction for multiple comparisons. No correlations met the same corrected threshold in either the LH or RH Semantic/Executive Networks.

## Discussion

This study provides clear evidence that activation in RH language homotopes increases beyond typical levels in the chronic phase of aphasia recovery, a finding that has been questioned due to limitations of prior imaging studies.^15^ We confirmed this effect in a large sample of stroke survivors and controls using a reliable, well-validated fMRI task that partially addresses the task performance confounds that plague imaging research on aphasia.^41^ Remaining performance confounds were addressed statistically, so the findings likely reflect stable changes in network organization, not transient effects related to effortful task performance.^16,17^

The overall magnitude of RH activity was modest compared to typical LH activity for this task. This reflects the limited plasticity available to adults, unlike the wholesale shift of language processing to the RH that can support acquisition of normal language after early-life LH strokes.^28,29^ Our finding that younger age relates to greater RH activity might suggest that early life plasticity persists to some degree in adults and declines gradually throughout the lifespan. Future research should aim to understand the biological constraints on language network plasticity that preclude greater use of the RH to support language in adults. This is essential to develop treatments that remove these constraints in a controlled way to allow stroke survivors to relearn language using the intact RH Language Network, like young children can.

It is perhaps unsurprising that RH activation related to left handedness, given the relationship between handedness and native language lateralization^35^ and prior evidence that weaker left-lateralization provides resilience to transient LH lesions induced by rTMS.^34^ However, individual differences in native language lateralization related to handedness cannot explain the increased RH activity observed in stroke survivors because the groups were matched on handedness, handedness was covaried in all analyses, and many stroke survivors had greater RH language activation than all controls except two who were right-lateralized.

In contrast to prior longitudinal studies that suggest normalization (i.e., reduction) of RH activation over the first several months after stroke,^1^ we found tentative evidence that RH activation may gradually increase later after stroke. Longitudinal work in the chronic period will be required to confirm this finding. RH activity may increase over time as individuals strengthen bilateral lexical-semantic representations through years of language experience or ongoing aphasia therapy.

RH activation was unrelated to lesion size, contradicting a prominent theory that individuals with severe LH damage use the RH to support language more than individuals with milder damage.^3,24,25^ A prior finding that suggested a relationship between lesion size and RH activity^21^ was likely spurious due to task performance confounds. RH engagement does, however, relate to the location of LH damage and the degree to which homotopic LH nodes remain functional. Because these relationships differ between groups of RH nodes, we infer that different neuroplastic mechanisms underlie recruitment of different parts of the homotopic RH Language Network (Fig. 7). Below, we will discuss the main patterns of findings observed across groups of RH language nodes and the possible neuroplastic mechanisms underlying their recruitment.

**Figure 7.**
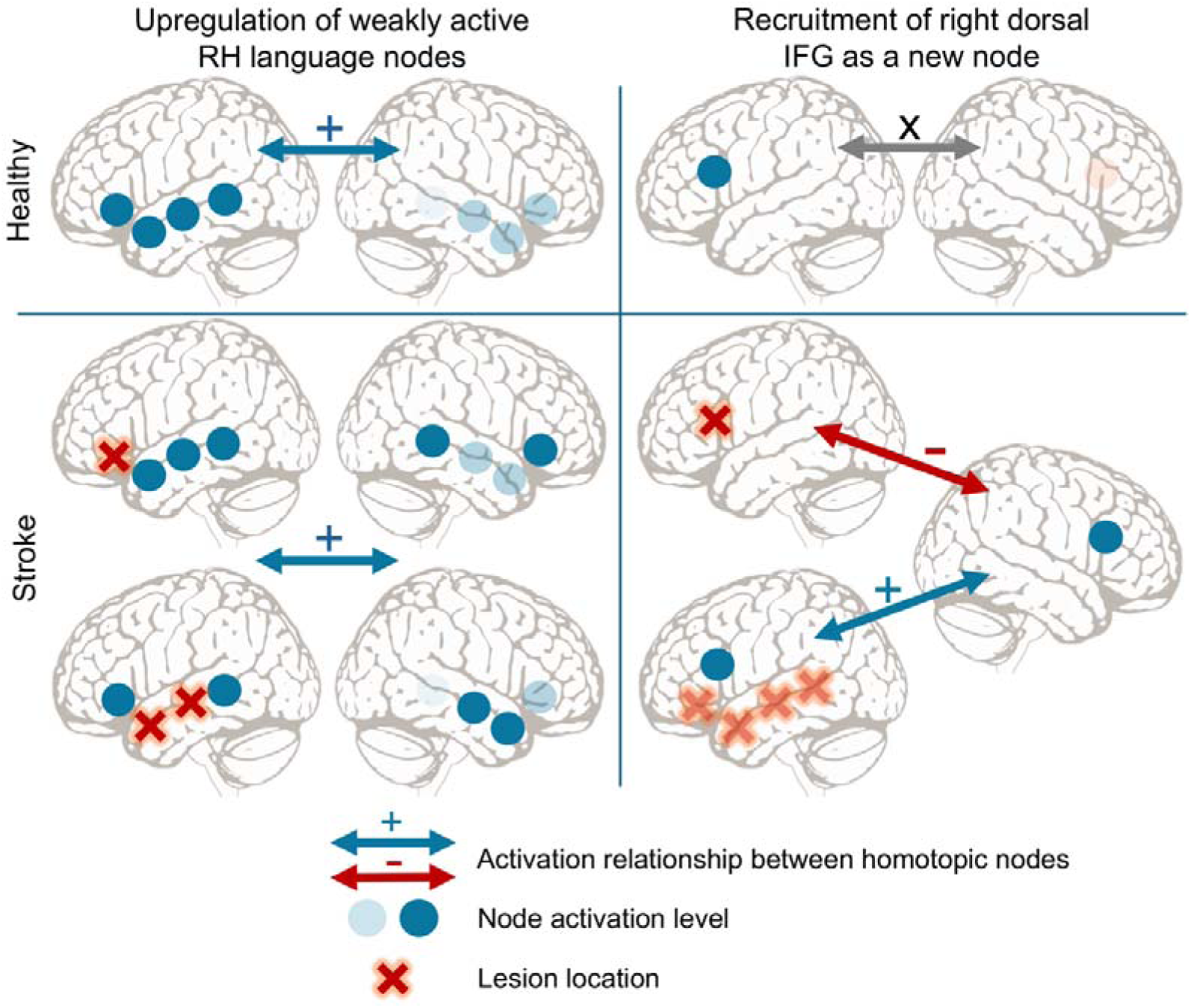
Schematic of two types of RH language plasticity. Left: In nodes that were already part of the language network based on subthreshold activation in controls (shown by the opaqueness of the dots) and correlated activation between homotopic LH and RH nodes (shown by the arrows between the hemispheres), LH lesions resulted in recruitment of homotopic RH nodes with continued coactivation with the residual activation of the injured LH nodes. Ventral frontal lesions also resulted in recruitment of the posterior STS/MTG. Activation of some of these nodes related to better reading and naming/word finding outcomes. Right: The right dorsal IFG was not engaged in language in neurological healthy adults based on null activation or slight deactivation and a null relationship between homotopic LH and RH activity. In contrast, strokes resulted in recruitment of dorsal IFG based on increased activation and new relationships between LH and RH activation. When lesions affecting the left dorsal IFG the right dorsal IFG activated in proportion to the dysfunction (inactivation) of the left dorsal IFG. When lesions spared the left dorsal IFG, the homotopic LH and RH dorsal IFGs became yoked, such that the RH activation increased in relation to LH activation. These changes were unrelated to aphasia outcomes.

The first pattern of RH recruitment was evident in the ventral IFG (RLang01, 02, to some degree 03) and mid-anterior temporal lobe nodes (RLang06, 07, 08). In controls, these nodes were weakly activated and their activation levels related to that of their LH homotopes. This suggests that they are part of the RH “weak shadow” language network that diminishes over development^30^ but may remain engaged weakly in language processing in adulthood, albeit at subthreshold activation levels.^31^ Activity in these regions increased in response to homotopic LH lesions and remained correlated with activation of the LH homotopes. Overall these findings suggests that the right ventral IFG and mid-anterior temporal lobe are nodes in a bihemispheric, albeit left-lateralized, language network, that are upregulated due to damage to parallel LH nodes that are likely engaged in similar computations.^3,32,53^ The co-activation relationship between homotopic LH and RH nodes indicates that RH upregulation does not occur through transcallosal disinhibition. Direct stimulation to enhance processing in the right ventral IFG and mid-anterior temporal nodes may help to improve aphasia, particularly for individuals who underactivate these nodes despite homotopic LH lesions. In the future, developing methods to focally facilitate the cellular and synaptic changes underlying network reweighting may allow for more dramatic enhancement of language processing in the RH.^37^

In the right posterior STS and MTG (RLang09, 10), activation increased not in relation to homotopic lesions, but rather in relation to the same frontal lesions that drove recruitment of the right ventral IFG. This finding replicates two prior studies that found that left frontal, but not temporal, lesions drive RH language recruitment, including posterior temporal cortical areas.^2,21^ These prior studies used picture naming and sentence comprehension tasks, so the replication of this lesion-activation relationship across three different tasks establishes it as a general principle of language network plasticity in adults. Some have argued that recruitment of right ventral IFG and posterior temporal nodes does not reflect a change in language processing, but rather activation of the ventral attention network (VAN),^54^ a right-lateralized network involved in stimulus-driven direction of attention.^55–57^ Our findings cannot easily be explained based on attentional demands since we addressed performance confounds as described above. Activation of these posterior temporal nodes also related (modestly) to reading and naming abilities, suggesting a role in language rather than attention. Moreover, these posterior temporal nodes are ventral and anterior to the VAN temporoparietal node.^55,57^ Notably, lesions not just to the left IFG, but also the anterior insula related to recruitment of these nodes. The right anterior insula is involved in executive control and coordinates switching between large-scale brain networks involved in attention and executive function,^58,59^ but the behavioral and network roles of left anterior insula are less well understood.^58^ Targeted research on the network effects of left anterior insula lesions might reveal an analogous role to its RH counterpart in network switching that could be relevant to its relationship to RH engagement for language processing.

Importantly, activity in several of the above ventral RH nodes corresponded with better aphasia outcomes, even after accounting for demographic factors, lesion size, and lesion location. These associations were evident for naming and word reading, without effects on repetition or auditory comprehension, suggesting these RH nodes may support lexical retrieval. The relationship of RH activation to education may then reflect strengthened bilateral lexical-semantic representations^53^ associated with higher education, postgraduate careers, or access to comprehensive aphasia treatment services. The semantic decision task does not tax syntactic or phonological processing, so this study may have been insensitive to RH contributions to comprehension or repetition.

A distinct pattern of findings was observed in the right dorsal IFG (RLang04, 05), which exhibited the largest group-level increases in activity. Recent reviews have argued that neuroplasticity allows for quantitative changes to preexisting circuits like those observed in the ventral nodes above, but not qualitative changes in function, sometimes referred to as remapping, reorganization, or vicariation.^36,37^ The pattern of results observed here in the dorsal IFG might suggest a qualitative change in processing, and deserves further characterization.

The right dorsal IFG nodes are near a region associated with the “multidemand network” which is involved in cognitive control and attention across a variety of tasks.^60^ Consistent with this, the dorsal pars triangularis (RLang04) was not activated even weakly by control participants during the semantic decision task, and the right pars opercularis (RLang05) was deactivated by controls relative to pseudofont matching. Activation levels in the left and right dorsal IFG were unrelated in controls, again suggesting these RH nodes were not engaged in language processing, at least as measured by the semantic decision fMRI task, prior to stroke. After LH lesions, however, many individuals activated the dorsal IFG, but in different ways depending on lesion location. A LH lesion sparing the dorsal pars opercularis resulted in recruitment of the right pars operularis into the language network with activation levels yoked to its LH counterpart. This relationship inverted when lesions directly damaged the left pars opercularis: reduced LH activity in this case related to increased RH activity.

How can this reversal be explained? Yoking of LH and RH activation levels might suggest transcallosal left-to-right facilitation. A partially damaged left dorsal pars opercularis may then provide noisy transcallosal input to the right pars opercularis that disrupts processing.^39^ Alternatively, the group-level deactivation of right pars opercularis in control participants could reflect left-to-right transcallosal suppression with a nonlinear response function that masks the expected relationship between the nodes. In this case, left pars opercularis lesions that cause frank node dysfunction may result in a release of inhibition on the RH, as is often suggested in the literature.^5,39,61,62^ This interpretation seems hard to reconcile with the coactivation of the left and right pars opercularis in the presence of lesions elsewhere in the LH, but evidence from the motor system, where interhemispheric interactions can be directly measured physiologically, has demonstrated reversals of interhemispheric inhibition depending on lesions and task states.^63^ The complex interaction between the hemispheres observed here in the dorsal IFG aligns with effective connectivity studies in aphasia, which suggest that network effects are more complex than a simple release of inhibition between homotopic nodes.^64,65^

Notably, increased right dorsal IFG activity did not relate to the aphasia outcomes examined here. A prior meta-analysis similarly found that the same part of right IFG demonstrated no functional homology with its LH counterpart across tasks, suggesting it served a different role.^13^ The right dorsal IFG may not contribute meaningfully to language outcomes, or it may contribute to behaviors not assessed here. Since the dorsal IFG interacts differently with the LH Language Network depending on the location of the lesion, analyses that account for LH activation levels in subgroups separated by lesion location may be needed to identify the roles of the dorsal IFG in aphasia outcomes, if any.

Collectively, our findings suggest that the right dorsal IFG is unlike other RH Language Network nodes in aphasia. These differences may be relevant to rTMS treatments for aphasia, which typically aim to inhibit the right IFG pars triangularis, particularly in people with nonfluent aphasia.^23^ The mechanisms underlying right IFG inhibition for aphasia remain unclear,^5,39^ making it difficult to select patients most likely to benefit from treatment. Our findings suggest that the effects of TMS might differ depending on the combination of lesion location and RH activation. While suppressing an overactive right dorsal IFG may be beneficial when the left dorsal IFG is lesioned and dysfunctional, the same treatment might have opposite effects when the left dorsal IFG is spared the right IFG coactivates with it.

In sum, this study provides perhaps the most comprehensive view yet of RH Language Network recruitment during aphasia recovery, confirming modest neuroplastic engagement of some RH nodes that varies between individuals based on age, handedness, education, and lesion location. The interactions between LH lesions, LH language activity, and RH language activity observed here suggest that different neuroplastic mechanisms account for recruitment of different RH nodes. These findings may guide current treatment approaches aiming to modulate RH engagement to improve aphasia recovery. Ultimately, a deeper understanding of these neuroplastic mechanisms and the constraints precluding more dramatic engagement of RH language homotopes after strokes in adults will be needed to develop new methods aiming to unlock more dramatic plasticity in the RH to improve aphasia recovery.

## Acknowledgements

We thank the valuable contributions of our participants, and our data collectors, in alphabetical order: Elizabeth Dvorak, Trini Kelly, Elizabeth Lacey, Sachi Paul, and Candace van der Stelt.

## Supplementary material

Supplementary material is available at *Brain* online.

