## Supplemental Materials for "Right Hemisphere Language Network Plasticity in Aphasia"

### Supplementary Materials

#### Supplementary Methods

##### *Participants*

Left hemisphere stroke survivors were recruited from the following sources: author P.E.T.'s outpatient Aphasia Clinic, the Speech and Stroke services at MedStar National Rehabilitation Hospital (NRH), the Stroke National Capital Area Network for Research, the MedStar Georgetown University Hospital Stroke service, regional aphasia and stroke recovery organizations, local stroke support groups, referrals from local clinicians, ClinicalTrials.gov, advertisements in the community, and word of mouth. Neurotypical control participants were recruited from the local community, ResearchMatch.org, as well as from families and friends of the stroke participants. Stroke survivors were recruited on the basis left hemisphere stroke at least six months prior to enrollment, native English language (learned earlier than age eight), no other history of significant psychiatric or neurological conditions that would affect interpretation of data, and no history of learning disorder requiring educational intervention. From an initial sample of 103 stroke survivors, six were excluded due to additional lesions outside of the left hemisphere, four were excluded because they did not complete the needed MRI scans, 15 were excluded due to performance below chance on the functional language mapping task as outlined in Functional Language Mapping Procedures section, and one was excluded for being earlier than six months since stroke. From an initial sample of 70 neurologically healthy controls, one was excluded for having a Montreal Cognitive Assessment (MoCA)[1] score below the cutoff, as determined by education, ethnicity, and race.[2]

##### *Neuroimaging*

Participants completed a neuroimaging session on a Siemens 3T Prisma Fit with 20-channel head coil. Sequences included a high-resolution T1-weighted magnetization-prepared rapid gradient echo (MPRAGE) scan (TR 1900 ms, TE 2.98, 176 1-mm sagittal slices, FOV 256, matrix 256 × 256, FA 9°, SMS 4), a T2-weighted fluid-attenuated inversion recovery (FLAIR) scan (TR 5000 ms, TE 38.2, 192 1-mm sagittal slices, FOV 256, matrix 256 × 256, FA 120°), and a BOLD T2*-weighted scan (TR 794 ms, 48 2.6-mm slices with 10% gap, 2.9 mm voxels, FOV 211 mm, matrix 74 × 74, FA 50°, SMS 4) consisting of 504 volumes lasting 6:40.

##### *Image Preprocessing and Statistical Analysis*

Standard preprocessing was performed in AFNI[3], including slice timing correction, realignment for head motion, despiking, smoothing with a 5-mm full width at half maximum kernel, temporal high-pass filtering at 0.01 Hz, and detrending. A whole-brain general linear model was estimated using the *fmrilm* function from FMRISTAT[4] with covariates including the time-course of a white matter and CSF seed, and the 6 head-motion parameters not convolved with the hemodynamic response function (HRF). The task was modeled using two alternating boxcar functions (corresponding to the language and control conditions), convolved with the HRF. The contrast of interest was semantic decision greater than pseudofont matching control (1 -1). The resulting statistical maps were then warped to MNI space based on the transformation estimated from the MPRAGE. Finally, images were resliced to 2-mm isotropic voxels.

##### *Procedure to divide clusters into subregions*

We employed a procedure to break up clusters into individual subregions surrounding the peaks of activity. All clustering procedures were performed in MATLAB 2023a (MathWorks, Natick, MA) using the Image Processing toolbox functionality. First, we applied a 3D Gaussian filter (via *imgaussfilt3*, $\sigma$=1.25) to the thresholded group t-map to attenuate noisy peaks. Local maxima (i.e., peaks of activity) were then isolated, via *imregionalmax*, on the resulting image using the default 3D 26-connected neighborhood criterion. Finally, all suprathreshold voxels were assigned to their nearest local peak based on Euclidean distance, such that clusters were divided into subregions surrounding the local peaks. This procedure assigned some anterior temporal lobe voxels to a subregion centered on an inferior frontal activation peak, so these voxels were reassigned to a contiguous anterior temporal subregion to respect the Sylvian fissure as an anatomical boundary.

##### *Demographic Variables*

Years of education were determined based on the highest degree-level attained in the United States (high school = 12, college = 16, master’s = 18, JD = 19, MD = 20, PhD = 21). Time-since-stroke was measured in months and was log transformed for statistical analyses due to a skewed distribution. Handedness prior to stroke was assessed using the Edinburgh Handedness Inventory handedness index.[5] Gender was coded as binary based on self-report (male, female; no participants endorsed a non-binary gender).

##### *Visualization*

Brain renderings were created in Surf Ice 1.0.20211006+ with overlay smoothing off. Brain slices were created in MRIcroGL with overlay smoothing off. Raincloud distribution plots were created in JASP 0.19.2. Linear trend plots were created using ggplot in R version 4.4.2 using RStudio Version 2024.12.0+467.

#### Supplementary Figures

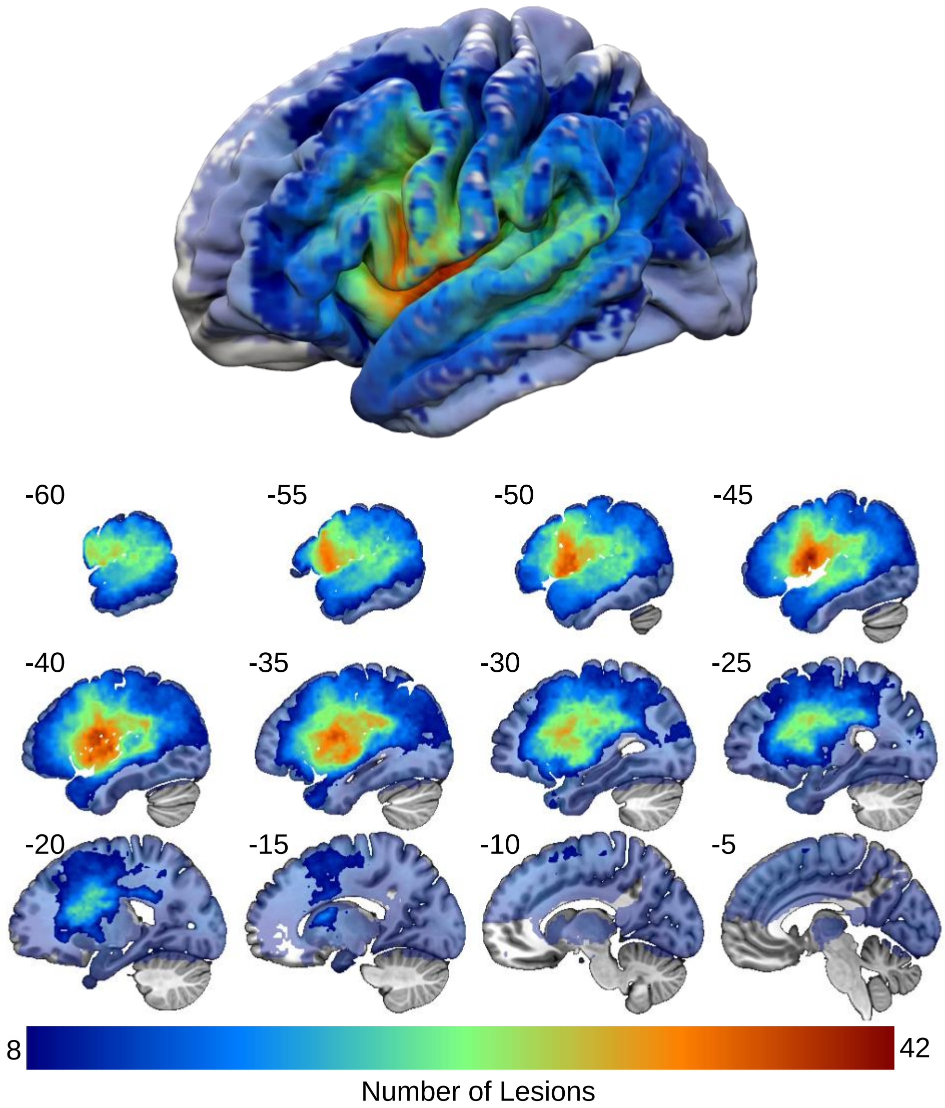


**Supplementary Figure 1.** Lesion overlap map. Voxels with sufficient lesion overlap to be included in SVR-LSM analyses (i.e., >= 8 lesions) are shown in the opaque overlay. Voxels with lesion overlap below the threshold for inclusion in SVR-LSM analyses are shown in translucent colors. The maximal lesion overlap was 42 lesions.

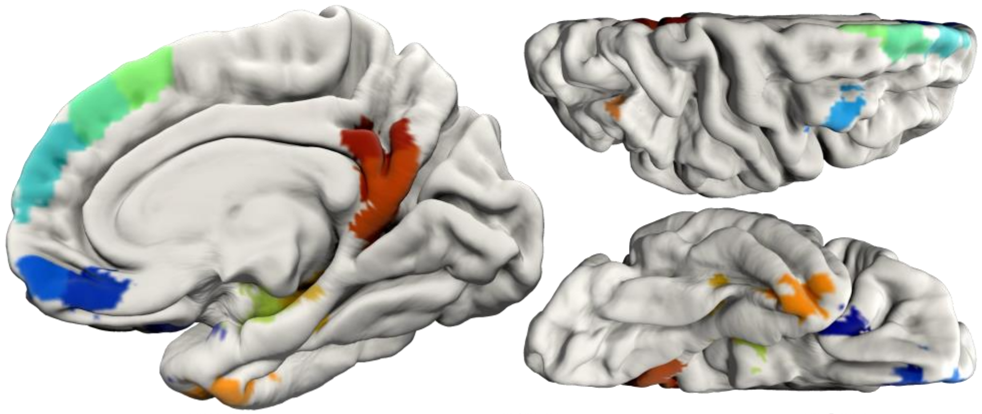

| ID | Structure | x | y | z | Size ($mm^3$) | Mean t |
|---|---|---|---|---|---|---|
| RSem01 | Orbitofrontal cortex- lateral | 24 | 15 | -22 | 2104 | 1.99 |
| RSem02 | Gyrus rectus- posterior | 5 | 41 | -19 | 3232 | 4.96 |
| RSem03 | Gyrus rectus- anterior | 4 | 57 | -15 | 2240 | 4.45 |
| RSem04 | Posterior MFG- ventral | 43 | 10 | 47 | 1840 | -3.58 |
| RSem05 | Posterior MFG- dorsal | 36 | 19 | 51 | 1904 | -2.82 |
| RSem06 | Medial SFG- polar | 8 | 57 | 26 | 6336 | 2.36 |
| RSem07 | Medial SFG- anterior | 8 | 40 | 45 | 5936 | 1.10 |
| RSem08 | Medial SFG- mid | 5 | 25 | 56 | 3464 | 1.73 |
| RSem09 | Hippocampus- anterior | 22 | -11 | -20 | 5744 | 3.27 |
| RSem10 | Hippocampus- postrior | 30 | -27 | -16 | 3360 | 1.43 |
| RSem11 | Inferior temporal pole | 36 | 2 | -42 | 3544 | 1.38 |
| RSem12 | Intraparietal sulcus-posterior | 35 | -67 | 43 | 1048 | -3.30 |
| RSem13 | Precuneus-ventral | 7 | -53 | 12 | 4432 | 3.55 |
| RSem14 | Precuneus/Posterior Cingulate | 4 | -50 | 31 | 2656 | 3.44 |

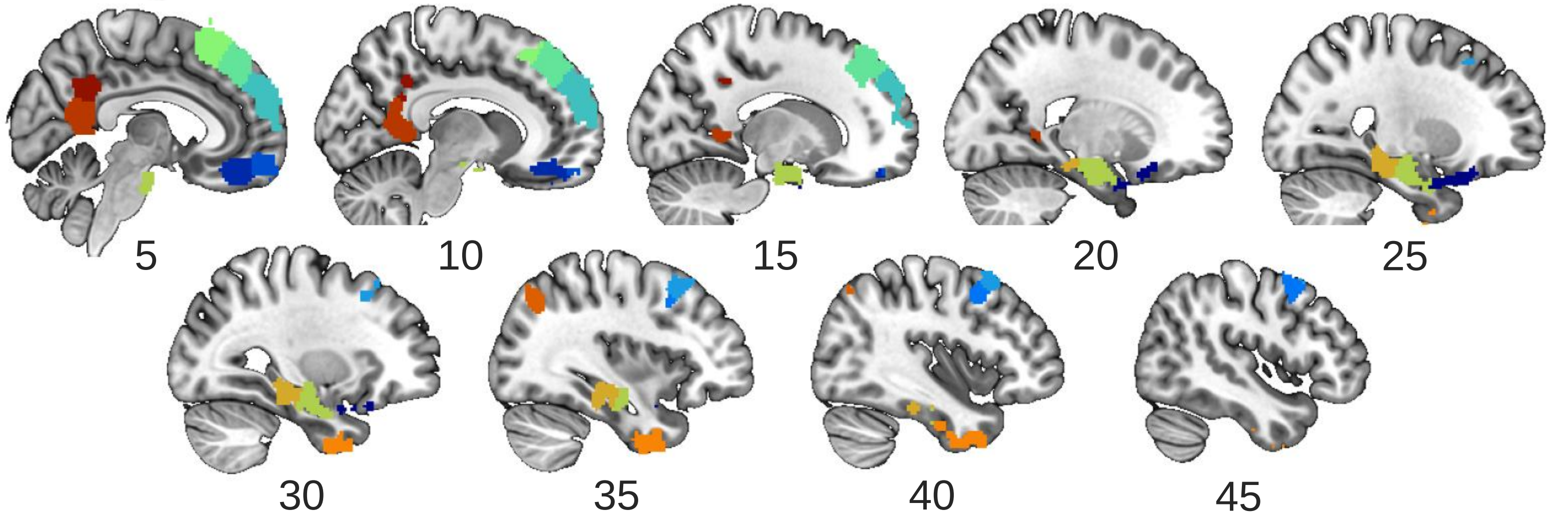


**Supplementary Figure 2.** The RH Semantic/Executive Network. The homotopic RH Semantic/Executive Network derived by clustering the fMRI group map and flipping over the midsagittal plane. All clusters not in the IFG or lateral temporoparietal regions canonically associated with language processing were included in the Semantic/Executive Network. Renderings are medial sagittal view on the left, and top view (top), and bottom view (bottom) with the front of the brain to the right. The x y z coordinates are in MNI space. “Mean t” refers to the mean t-score of activation in the node in the held-out controls. MFG = middle frontal gyrus, SFG = superior frontal gyrus.

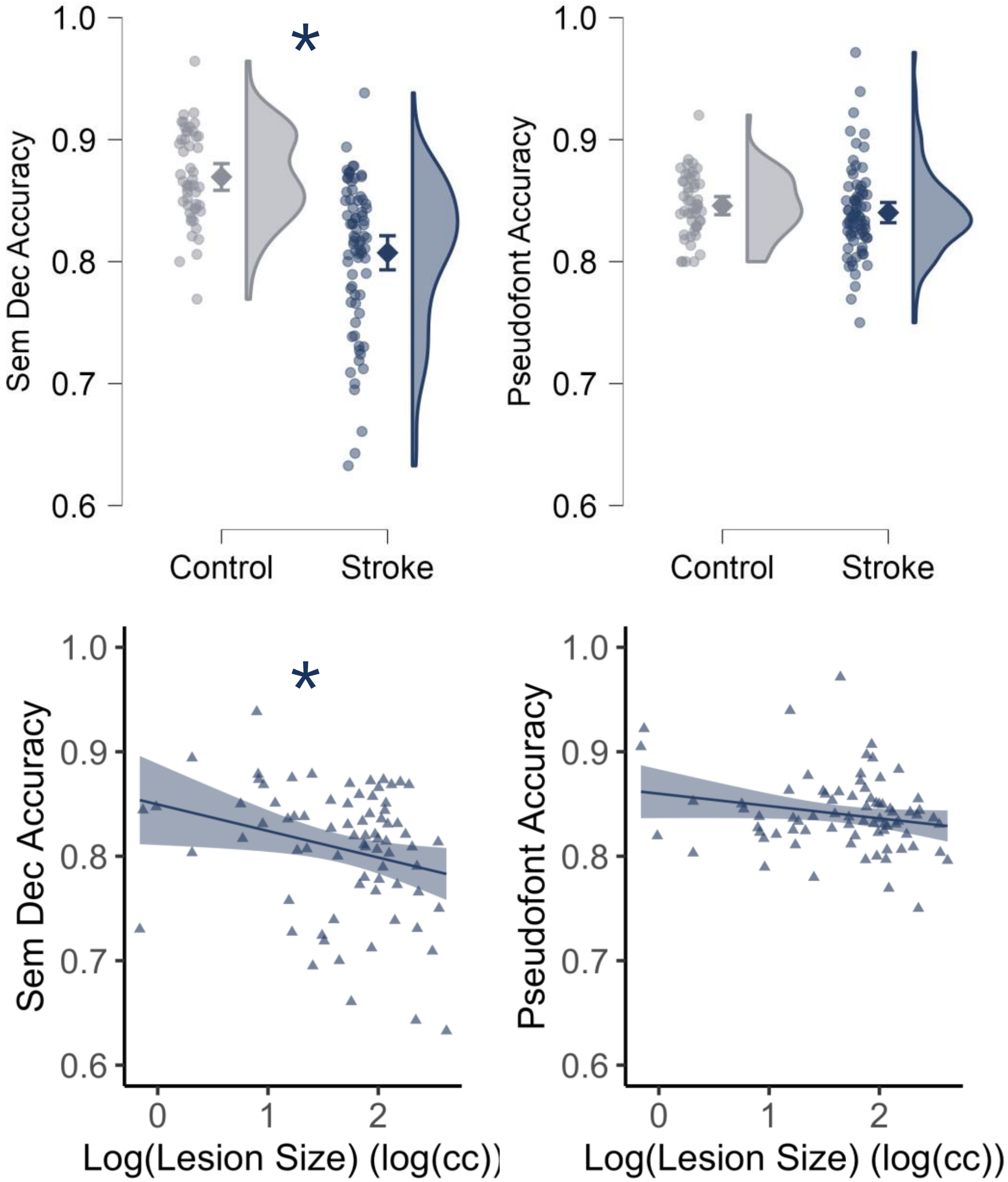


**Supplementary Figure 3.** fMRI task performance. The top row shows accuracy by group on the semantic decision task (left) and the pseudofont matching control task (right). The diamond is the mean and error bars are the 95% confidence interval. These figures were created in JASP. The bottom row shows the correlation between log-transformed lesion size and accuracy on the tasks (semantic decision on the left and pseudofont matching on the right). Best fit lines for the linear relationship are shown with 95% confidence interval. These figures were created using *ggplot* in R. *$p < .05$

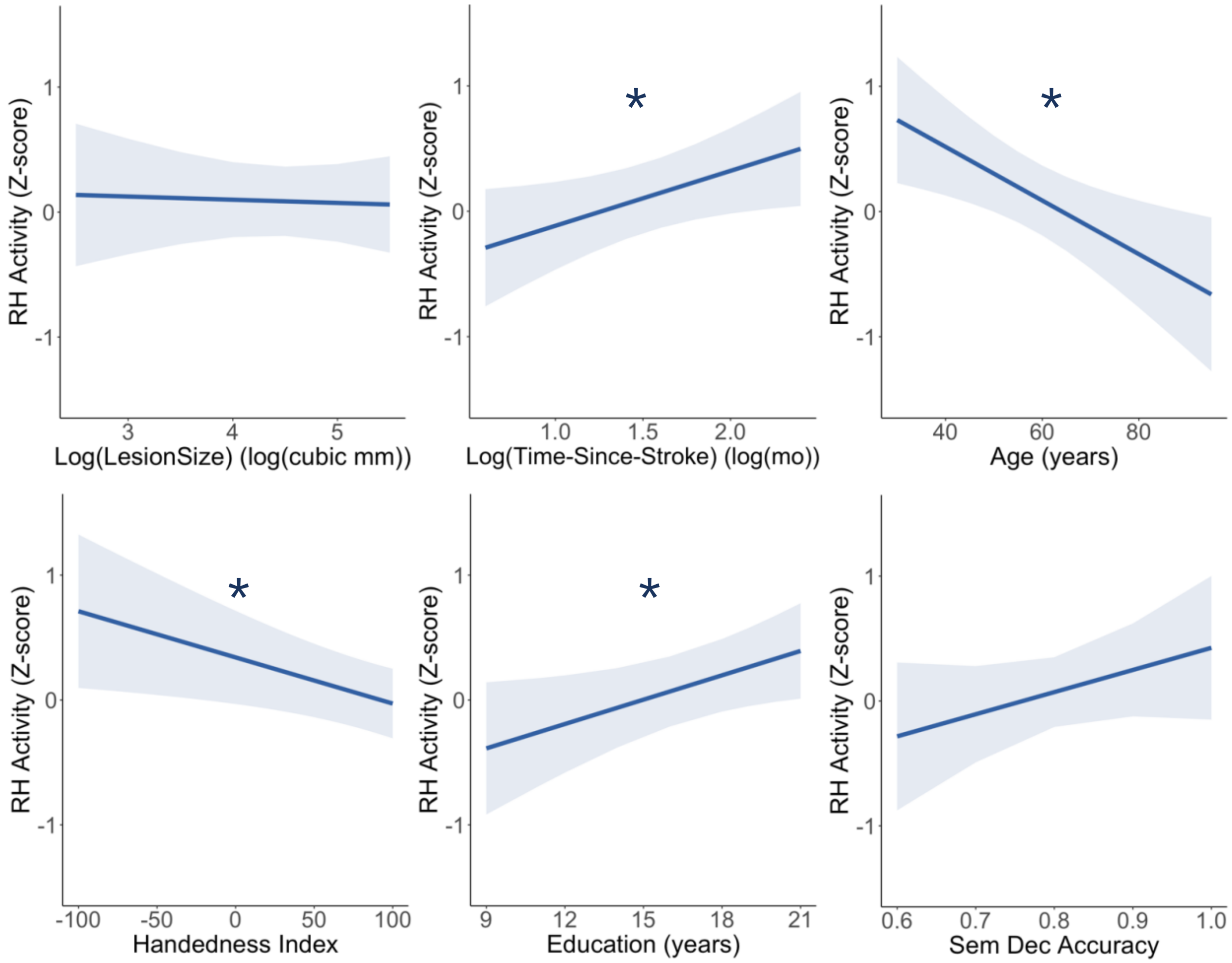


**Supplementary Figure 4.** Effects of lesion size, time-since-stroke, and covariates on RH Language Network activity. The plots show estimated marginal effects from the linear mixed-effects model, visualized using plot_model in R. *$P < 0.05$

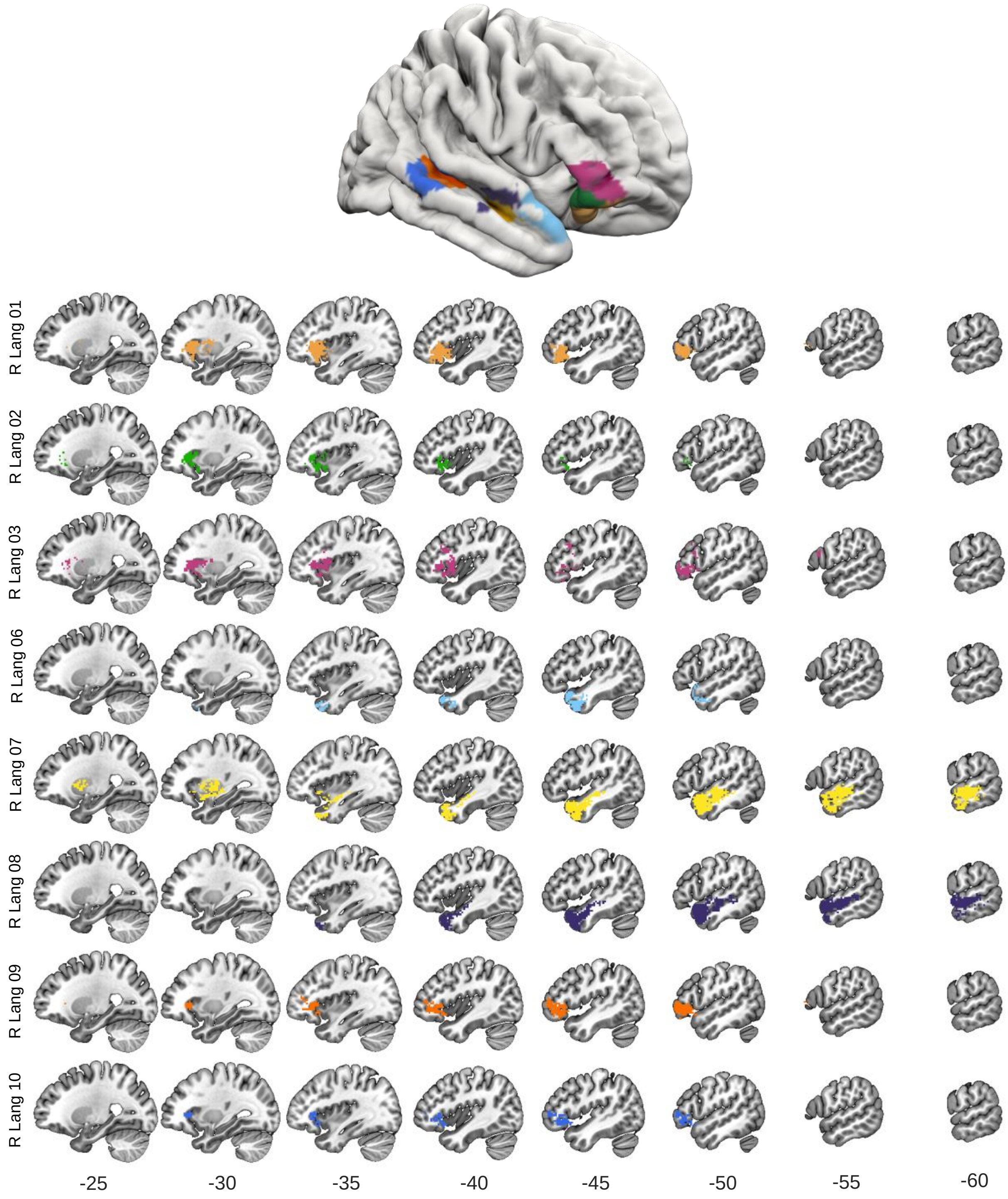


**Supplementary Figure 5.** Lesions associated with greater RH Language Network activity. The rendering shows the nodes of the RH Language Network where greater activity was associated with specific lesion locations based on SVR-LSM analyses. Each row shows the SVR-LSM results for one of the RH Language nodes, as labeled on the left of the row. The color of each overlay matches the color of the associated RH node on the rendering. Two lesion-activation patterns emerged from these analyses. Left inferior frontal and anterior insula lesions are associated with greater activation of right ventral IFG (RLang01, 02, 03) and posterior STS (RLang09) and MTG (RLang10). In contrast, left anterior and mid-temporal lobe lesions are associated with greater activation of right anterior and mid-temporal lobe nodes (RLang06, 07, 08). SVR-LSM analyses were thresholded at voxelwise $p<.005$, cluster family-wise error rate corrected $p<.05$.

### Supplementary Tables

#### Supplementary Table 1- Mixed-effects model comparing RH language network activity between the Stroke and Control groups

```
jaspMixedModels::MixedModelsLMM(
        version = "0.19.2",
        formula = RHActivity ~ Group + Age + Handedness + Gender + Sem_Dec_Acc +
Education + (1 + Gender | Node) + (1 | Participant),
        contrasts = NULL,
        fixedEffectEstimate = TRUE,
        marginalMeansComparison = TRUE,
        marginalMeansTerms = ~ Group,
        modelSummary = TRUE,
        trendsContrasts = NULL)
```

*ANOVA Summary*

| Effect | df | F | p |
|---|---|---|---|
| Group | 1, 118.01 | 4.436 | 0.037 |
| Age | 1, 118.01 | 8.088 | 0.005 |
| Handedness | 1, 118.01 | 11.892 | $7.822 \times 10^{-4}$ |
| Gender | 1, 87.87 | 0.003 | 0.954 |
| Sem_Dec_Acc | 1, 118.01 | 1.433 | 0.234 |
| Education | 1, 118.01 | 8.027 | 0.005 |

*Note.* Model terms tested with Satterthwaite testMethod.
*Note.* The following variables are used as random effects grouping factors: 'Node', 'Participant'.
*Note.* Type III Sum of Squares

##### Model summary

*Fit statistics*

| Deviance (REML) | log Lik. | df | AIC | BIC |
|---|---|---|---|---|
| 5162.218 | -2581.109 | 12 | 5186.218 | 5248.932 |

*Note.* The model was fitted using restricted maximum likelihood. Please note that models with different fixed effects cannot be compared when REML is used. To use ML, switch 'Test method' to 'Likelihood ratio tests'.

*Sample sizes*

| | Levels of RE grouping factors | |
|---|---|---|
| Observations | Participant | Node |
| 1375 | 125 | 11 |

*Fixed Effects Estimates*

| Term | Estimate | SE | df | t | p |
|---|---|---|---|---|---|
| Intercept | -0.815 | 1.603 | 119.569 | -0.508 | 0.612 |
| Group (1) | 0.234 | 0.111 | 118.014 | 2.106 | 0.037 |
| Age | -0.025 | 0.009 | 118.014 | -2.844 | 0.005 |
| Handedness | -0.007 | 0.002 | 118.014 | -3.449 | $7.822 \times 10^{-4}$ |
| Gender (1) | 0.006 | 0.096 | 87.870 | 0.058 | 0.954 |
| Sem_Dec_Acc | 2.128 | 1.778 | 118.014 | 1.197 | 0.234 |
| Education | 0.098 | 0.035 | 118.014 | 2.833 | 0.005 |

*Fit statistics*

| Deviance (REML) | log Lik. | df | AIC | BIC |
|---|---|---|---|---|

*Note.* The intercept corresponds to the (unweighted) grand mean; for each factor with k levels, k - 1 parameters are estimated with sum contrast coding. Consequently, the estimates cannot be directly mapped to factor levels. Use estimated marginal means for obtaining estimates for each factor level/design cell or their differences.

*Estimated Marginal Means*

| | | | 95% CI | | | |
|---|---|---|---|---|---|---|
| Group | Estimate | SE | Lower | Upper | z | p† |
| Stroke | 0.856 | 0.185 | 0.494 | 1.218 | 4.639 | $3.501 \times 10^{-6}$ |
| Control | 0.388 | 0.211 | -0.026 | 0.801 | 1.838 | 0.066 |

*Note.* Results are averaged over the levels of: Gender.

† P-values correspond to test of null hypothesis against 0.

#### Supplementary Table 2- Mixed-effects model comparing LH language network activity between the Stroke and Control groups

```
jaspMixedModels::MixedModelsLMM(
        version = "0.19.2",
        formula = LHActivity ~ Group + Age + Handedness + Gender + Sem_Dec_Acc +
Education + (1 | Node) + (1 | Participant),
        contrasts = NULL,
        fixedEffectEstimate = TRUE,
        marginalMeansComparison = TRUE,
        marginalMeansTerms = ~ Group,
        modelSummary = TRUE,
        trendsContrasts = NULL)
```

*ANOVA Summary*

| Effect | df | F | p |
|---|---|---|---|
| Group | 1, 118.00 | 2.962 | 0.088 |
| Age | 1, 118.00 | 0.071 | 0.791 |
| Handedness | 1, 118.00 | 0.133 | 0.716 |
| Gender | 1, 118.00 | $7.424 \times 10^{-4}$ | 0.978 |
| Sem_Dec_Acc | 1, 118.00 | 29.139 | $3.534 \times 10^{-7}$ |
| Education | 1, 118.00 | 2.979 | 0.087 |

*Note.* Model terms tested with Satterthwaite testMethod.
*Note.* The following variables are used as random effects grouping factors: 'Node', 'Participant'.
*Note.* Type III Sum of Squares

##### Model summary

*Fit statistics*

| Deviance (REML) | log Lik. | df | AIC | BIC |
|---|---|---|---|---|
| 6267.068 | -3133.534 | 10 | 6287.068 | 6339.330 |

*Note.* The model was fitted using restricted maximum likelihood. Please note that models with different fixed effects cannot be compared when REML is used. To use ML, switch 'Test method' to 'Likelihood ratio tests'.

*Sample sizes*

| | Levels of RE grouping factors | |
|---|---|---|
| Observations | Participant | Node |
| 1375 | 125 | 11 |

*Fixed Effects Estimates*

| Term | Estimate | SE | df | t | p |
|---|---|---|---|---|---|
| Intercept | -11.579 | 2.734 | 122.513 | -4.235 | $4.440 \times 10^{-5}$ |
| Group (1) | -0.324 | 0.188 | 117.999 | -1.721 | 0.088 |
| Age | 0.004 | 0.015 | 117.999 | 0.266 | 0.791 |
| Handedness | 0.001 | 0.003 | 117.999 | 0.365 | 0.716 |
| Gender (1) | 0.004 | 0.161 | 117.999 | 0.027 | 0.978 |
| Sem_Dec_Acc | 16.241 | 3.009 | 117.999 | 5.398 | $3.534 \times 10^{-7}$ |
| Education | 0.101 | 0.059 | 117.999 | 1.726 | 0.087 |

*Note.* The intercept corresponds to the (unweighted) grand mean; for each factor with k levels, k - 1 parameters are estimated with sum contrast coding. Consequently, the estimates cannot be directly mapped to factor levels. Use estimated marginal means for obtaining estimates for each factor level/design cell or their differences.

*Estimated Marginal Means*

| | | | 95% CI | | | |
|---|---|---|---|---|---|---|
| Group | Estimate | SE | Lower | Upper | z | p† |
| Stroke | 3.603 | 0.456 | 2.710 | 4.496 | 7.906 | $2.664 \times 10^{-15}$ |
| Control | 4.251 | 0.487 | 3.295 | 5.206 | 8.720 | $2.774 \times 10^{-18}$ |

*Note.* Results are averaged over the levels of: Gender.

† P-values correspond to test of null hypothesis against 0.

#### Supplementary Table 3- Mixed-effects model comparing RH semantic network activity between the Stroke and Control groups

```
jaspMixedModels::MixedModelsLMM(
        version = "0.19.2",
        formula = RHactivity ~ Group + Age + Education + Handedness + Gender +
Sem_Dec_Acc + (1 + Sem_Dec_Acc | Node) + (1 | Participant),
        contrasts = NULL,
        fixedEffectEstimate = TRUE,
        marginalMeansComparison = TRUE,
        marginalMeansTerms = ~ Group,
        modelSummary = TRUE,
        trendsContrasts = NULL)
```

*ANOVA Summary*

| Effect | df | F | p |
|---|---|---|---|
| Group | 1, 118.00 | 2.009 | 0.159 |
| Age | 1, 118.00 | 0.949 | 0.332 |
| Education | 1, 118.00 | 1.987 | 0.161 |
| Handedness | 1, 118.00 | 13.456 | $3.677\times10^{-4}$ |
| Gender | 1, 118.00 | 1.554 | 0.215 |
| Sem_Dec_Acc | 1, 23.24 | 0.118 | 0.734 |

*Note.* Model terms tested with Satterthwaite testMethod.
*Note.* The following variables are used as random effects grouping factors: 'Node', 'Participant'.
*Note.* Type III Sum of Squares

##### Model summary

*Fit statistics*

| Deviance (REML) | log Lik. | df | AIC | BIC |
|---|---|---|---|---|
| 7128.762 | -3564.381 | 12 | 7152.762 | 7218.371 |

*Note.* The model was fitted using restricted maximum likelihood. Please note that models with different fixed effects cannot be compared when REML is used. To use ML, switch 'Test method' to 'Likelihood ratio tests'.

*Sample sizes*

| | Levels of RE grouping factors | |
|---|---|---|
| Observations | Participant | Node |
| 1750 | 125 | 14 |

*Fixed Effects Estimates*

| Term | Estimate | SE | df | t | p |
|---|---|---|---|---|---|
| Intercept | -0.445 | 1.903 | 33.341 | -0.234 | 0.816 |
| Group (1) | 0.133 | 0.094 | 117.999 | 1.417 | 0.159 |
| Age | -0.007 | 0.007 | 117.999 | -0.974 | 0.332 |
| Education | 0.041 | 0.029 | 117.999 | 1.410 | 0.161 |
| Handedness | -0.006 | 0.002 | 117.999 | -3.668 | $3.677\times10^{-4}$ |
| Gender (1) | 0.100 | 0.081 | 117.999 | 1.247 | 0.215 |
| Sem_Dec_Acc | 0.868 | 2.524 | 23.237 | 0.344 | 0.734 |

*Note.* The intercept corresponds to the (unweighted) grand mean; for each factor with k levels, k - 1 parameters are estimated with sum contrast coding. Consequently, the estimates cannot be directly mapped to factor levels. Use estimated marginal means for obtaining estimates for each factor level/design cell or their differences.

*Estimated Marginal Means*

| Group | Estimate | SE | 95% CI Lower | 95% CI Upper | z | p† |
|---|---|---|---|---|---|---|
| Stroke | 0.241 | 0.363 | -0.471 | 0.953 | 0.663 | 0.507 |
| Control | -0.025 | 0.373 | -0.757 | 0.706 | -0.068 | 0.946 |

*Note.* Results are averaged over the levels of: Gender.
† P-values correspond to test of null hypothesis against 0.

### Supplementary Table 4- Mixed-effects model comparing LH semantic network activity between the Stroke and Control groups

```
jaspMixedModels::MixedModelsLMM(
        version = "0.19.2",
        formula = LHactivity ~ Group + Age + Education + Handedness + Gender +
Sem_Dec_Acc + (1 + Group | Node) + (1 | Participant),
        contrasts = NULL,
        fixedEffectEstimate = TRUE,
        marginalMeansComparison = TRUE,
        marginalMeansTerms = ~ Group,
        modelSummary = TRUE,
        trendsContrasts = NULL)
```

*ANOVA Summary*

| Effect | df | F | p |
|---|---|---|---|
| Group | 1, 84.87 | 1.156 | 0.285 |
| Age | 1, 117.99 | 0.017 | 0.895 |
| Education | 1, 117.99 | 4.502 | 0.036 |
| Handedness | 1, 117.99 | 0.465 | 0.496 |
| Gender | 1, 117.99 | 1.114 | 0.293 |
| Sem_Dec_Acc | 1, 117.99 | 18.397 | $3.689 \times 10^{-5}$ |

*Note.* Model terms tested with Satterthwaite testMethod.
*Note.* The following variables are used as random effects grouping factors: 'Node', 'Participant'.
*Note.* Type III Sum of Squares

#### Model summary

*Fit statistics*

| Deviance (REML) | log Lik. | df | AIC | BIC |
|---|---|---|---|---|
| 7328.293 | -3664.147 | 12 | 7352.293 | 7417.902 |

*Note.* The model was fitted using restricted maximum likelihood. Please note that models with different fixed effects cannot be compared when REML is used. To use ML, switch 'Test method' to 'Likelihood ratio tests'.

*Sample sizes*

| | Levels of RE grouping factors | |
|---|---|---|
| Observations | Participant | Node |
| 1750 | 125 | 14 |

*Fixed Effects Estimates*

| Term | Estimate | SE | df | t | p |
|---|---|---|---|---|---|
| Intercept | -5.884 | 1.712 | 122.524 | -3.438 | $8.013 \times 10^{-4}$ |
| Group (1) | -0.137 | 0.128 | 84.866 | -1.075 | 0.285 |
| Age | 0.001 | 0.009 | 117.995 | 0.132 | 0.895 |
| Education | 0.078 | 0.037 | 117.995 | 2.122 | 0.036 |
| Handedness | 0.001 | 0.002 | 117.995 | 0.682 | 0.496 |
| Gender (1) | -0.107 | 0.101 | 117.995 | -1.055 | 0.293 |
| Sem_Dec_Acc | 8.082 | 1.884 | 117.995 | 4.289 | $3.689 \times 10^{-5}$ |

*Note.* The intercept corresponds to the (unweighted) grand mean; for each factor with k levels, k - 1 parameters are estimated with sum contrast coding. Consequently, the estimates cannot be directly mapped to factor levels. Use estimated marginal means for obtaining estimates for each factor level/design cell or their differences.

*Estimated Marginal Means*

| Group | Estimate | SE | 95% CI Lower | 95% CI Upper | z | p† |
|---|---|---|---|---|---|---|
| Stroke | 2.164 | 0.245 | 1.684 | 2.644 | 8.838 | $9.747\times10^{-19}$ |
| Control | 2.439 | 0.340 | 1.773 | 3.105 | 7.179 | $7.006\times10^{-13}$ |

*Note.* Results are averaged over the levels of: Gender.
† P-values correspond to test of null hypothesis against 0.

### Supplementary Table 5- RLang01 ANCOVA comparing Stroke vs Control groups

```
jaspAnova::Ancova(
        version = "0.19.2",
        formula = RLang01 ~ Group.nominal + Gender + Age + Education + Handedness +
Sem_Dec_Acc,
        covariates = list("Age", "Education", "Handedness", "Sem_Dec_Acc"),
        descriptivePlotErrorBar = TRUE,
        effectSizeEstimates = TRUE,
        effectSizeOmegaSquared = FALSE,
        effectSizePartialEtaSquared = TRUE,
        homogeneityTests = TRUE,
        marginalMeanComparedToZero = TRUE,
        marginalMeanTerms = ~ Group.nominal,
        restrictedModelComparison = "unconstrained",
        restrictedModels = list(list(informedHypothesisTest = FALSE, marginalMean =
FALSE, name = "Model 1", summary = FALSE, syntax = "")))
```

*ANCOVA - RLang01*

| Cases | Sum of Squares | df | Mean Square | F | p | $\eta^2_p$ |
|---|---|---|---|---|---|---|
| Group | 3.766 | 1 | 3.766 | 2.086 | 0.151 | 0.017 |
| Gender | 1.424 | 1 | 1.424 | 0.789 | 0.376 | 0.007 |
| Age | 12.283 | 1 | 12.283 | 6.803 | 0.010 | 0.055 |
| Education | 6.986 | 1 | 6.986 | 3.870 | 0.052 | 0.032 |
| Handedness | 13.783 | 1 | 13.783 | 7.634 | 0.007 | 0.061 |
| Sem_Dec_Acc | 4.865 | 1 | 4.865 | 2.695 | 0.103 | 0.022 |
| Residuals | 213.039 | 118 | 1.805 | | | |

*Note.* Type III Sum of Squares

#### Assumption Checks

*Test for Equality of Variances (Levene's)*

| F | df1 | df2 | p |
|---|---|---|---|
| 0.282 | 3.000 | 121.000 | 0.838 |

#### Marginal Means

*Marginal Means - Group*

| | | 95% CI for Mean Difference | | | | | |
|---|---|---|---|---|---|---|---|
| Group | Marginal Mean | Lower | Upper | SE | t | df | p |
| Control | 0.887 | 0.470 | 1.305 | 0.211 | 4.206 | 118 | $5.096\times10^{-5}$ |
| Stroke | 1.301 | 0.974 | 1.627 | 0.165 | 7.882 | 118 | $1.785\times10^{-12}$ |

#### Supplementary Table 6- RLang02 ANCOVA comparing Stroke vs Control groups

```
jaspAnova::Ancova(
        version = "0.19.2",
        formula = RLang02 ~ Group.nominal + Gender + Age + Education + Handedness +
Sem_Dec_Acc,
        covariates = list("Age", "Education", "Handedness", "Sem_Dec_Acc"),
        descriptivePlotErrorBar = TRUE,
        effectSizeEstimates = TRUE,
        effectSizeOmegaSquared = FALSE,
        effectSizePartialEtaSquared = TRUE,
        homogeneityTests = TRUE,
        marginalMeanComparedToZero = TRUE,
        marginalMeanTerms = ~ Group.nominal,
        restrictedModelComparison = "unconstrained",
        restrictedModels = list(list(informedHypothesisTest = FALSE, marginalMean =
FALSE, name = "Model 1", summary = FALSE, syntax = "")))
```

*ANCOVA - RLang02*

| Cases | Sum of Squares | df | Mean Square | F | p | $\eta^2_p$ |
|---|---|---|---|---|---|---|
| Group | 6.955 | 1 | 6.955 | 3.079 | 0.082 | 0.025 |
| Gender | 0.513 | 1 | 0.513 | 0.227 | 0.635 | 0.002 |
| Age | 21.936 | 1 | 21.936 | 9.712 | 0.002 | 0.076 |
| Education | 13.556 | 1 | 13.556 | 6.001 | 0.016 | 0.048 |
| Handedness | 21.970 | 1 | 21.970 | 9.727 | 0.002 | 0.076 |
| Sem_Dec_Acc | 0.105 | 1 | 0.105 | 0.047 | 0.829 | $3.949\times10^{-4}$ |
| Residuals | 266.533 | 118 | 2.259 | | | |

*Note.* Type III Sum of Squares

##### Assumption Checks

*Test for Equality of Variances (Levene's)*

| F | df1 | df2 | p |
|---|---|---|---|
| 1.123 | 3.000 | 121.000 | 0.342 |

##### Marginal Means

*Marginal Means - Group*

| | | 95% CI for Mean Difference | | | | | |
|---|---|---|---|---|---|---|---|
| Group | Marginal Mean | Lower | Upper | SE | t | df | p |
| Control | 0.492 | 0.025 | 0.959 | 0.236 | 2.085 | 118 | 0.039 |
| Stroke | 1.054 | 0.688 | 1.419 | 0.185 | 5.709 | 118 | $8.630\times10^{-8}$ |

#### Supplementary Table 7- RLang03 ANCOVA comparing Stroke vs Control groups

```
jaspAnova::Ancova(
        version = "0.19.2",
        formula = RLang03 ~ Group.nominal + Gender + Age + Education + Handedness + 
Sem_Dec_Acc,
        covariates = list("Age", "Education", "Handedness", "Sem_Dec_Acc"),
        descriptivePlotErrorBar = TRUE,
        effectSizeEstimates = TRUE,
        effectSizeOmegaSquared = FALSE,
        effectSizePartialEtaSquared = TRUE,
        homogeneityTests = TRUE,
        marginalMeanComparedToZero = TRUE,
        marginalMeanTerms = ~ Group.nominal,
        restrictedModelComparison = "unconstrained",
        restrictedModels = list(list(informedHypothesisTest = FALSE, marginalMean = 
FALSE, name = "Model 1", summary = FALSE, syntax = "")))
```

*ANCOVA - RLang03*

| Cases | Sum of Squares | df | Mean Square | F | p | $\eta^2_p$ |
|---|---|---|---|---|---|---|
| Group | 11.328 | 1 | 11.328 | 3.322 | 0.071 | 0.027 |
| Gender | 10.113 | 1 | 10.113 | 2.965 | 0.088 | 0.025 |
| Age | 22.978 | 1 | 22.978 | 6.738 | 0.011 | 0.054 |
| Education | 23.816 | 1 | 23.816 | 6.983 | 0.009 | 0.056 |
| Handedness | 6.799 | 1 | 6.799 | 1.994 | 0.161 | 0.017 |
| Sem_Dec_Acc | 0.227 | 1 | 0.227 | 0.067 | 0.797 | $5.638 \times 10^{-4}$ |
| Residuals | 402.430 | 118 | 3.410 | | | |

*Note.* Type III Sum of Squares

##### Assumption Checks

*Test for Equality of Variances (Levene's)*

| F | df1 | df2 | p |
|---|---|---|---|
| 0.446 | 3.000 | 121.000 | 0.721 |

##### Marginal Means

*Marginal Means - Group*

| | | 95% CI for Mean Difference | | | | | |
|---|---|---|---|---|---|---|---|
| Group | Marginal Mean | Lower | Upper | SE | t | df | p |
| Control | 0.117 | -0.458 | 0.691 | 0.290 | 0.402 | 118 | 0.688 |
| Stroke | 0.833 | 0.384 | 1.283 | 0.227 | 3.675 | 118 | $3.593 \times 10^{-4}$ |

### Supplementary Table 8- RLang04 ANCOVA comparing Stroke vs Control groups

```
jaspAnova::Ancova(
        version = "0.19.2",
        formula = RLang04 ~ Group.nominal + Gender + Age + Education + Handedness +
Sem_Dec_Acc,
        covariates = list("Age", "Education", "Handedness", "Sem_Dec_Acc"),
        descriptivePlotErrorBar = TRUE,
        effectSizeEstimates = TRUE,
        effectSizeOmegaSquared = FALSE,
        effectSizePartialEtaSquared = TRUE,
        homogeneityTests = TRUE,
        marginalMeanComparedToZero = TRUE,
        marginalMeanTerms = ~ Group.nominal,
        restrictedModelComparison = "unconstrained",
        restrictedModels = list(list(informedHypothesisTest = FALSE, marginalMean =
FALSE, name = "Model 1", summary = FALSE, syntax = "")))
```

*ANCOVA - RLang04*

| Cases | Sum of Squares | df | Mean Square | F | p | $\eta^2_p$ |
|---|---|---|---|---|---|---|
| Group | 23.959 | 1 | 23.959 | 8.374 | 0.005 | 0.066 |
| Gender | 0.091 | 1 | 0.091 | 0.032 | 0.858 | $2.708\times10^{-4}$ |
| Age | 28.082 | 1 | 28.082 | 9.815 | 0.002 | 0.077 |
| Education | 13.434 | 1 | 13.434 | 4.695 | 0.032 | 0.038 |
| Handedness | 29.049 | 1 | 29.049 | 10.153 | 0.002 | 0.079 |
| Sem_Dec_Acc | 3.124 | 1 | 3.124 | 1.092 | 0.298 | 0.009 |
| Residuals | 337.601 | 118 | 2.861 | | | |

*Note.* Type III Sum of Squares

#### Assumption Checks

*Test for Equality of Variances (Levene's)*

| F | df1 | df2 | p |
|---|---|---|---|
| 0.580 | 3.000 | 121.000 | 0.629 |

#### Marginal Means

*Marginal Means - Group*

| | | 95% CI for Mean Difference | | | | | |
|---|---|---|---|---|---|---|---|
| Group | Marginal Mean | Lower | Upper | SE | t | df | p |
| Control | -0.146 | -0.672 | 0.379 | 0.266 | -0.552 | 118 | 0.582 |
| Stroke | 0.896 | 0.485 | 1.307 | 0.208 | 4.313 | 118 | $3.361\times10^{-5}$ |

### Supplementary Table 9- RLang05 ANCOVA comparing Stroke vs Control groups

```
jaspAnova::Ancova(
        version = "0.19.2",
        formula = RLang05 ~ Group.nominal + Gender + Age + Education + Handedness +
Sem_Dec_Acc,
        covariates = list("Age", "Education", "Handedness", "Sem_Dec_Acc"),
        descriptivePlotErrorBar = TRUE,
        effectSizeEstimates = TRUE,
        effectSizeOmegaSquared = FALSE,
        effectSizePartialEtaSquared = TRUE,
        homogeneityTests = TRUE,
        marginalMeanComparedToZero = TRUE,
        marginalMeanTerms = ~ Group.nominal,
        restrictedModelComparison = "unconstrained",
        restrictedModels = list(list(informedHypothesisTest = FALSE, marginalMean =
FALSE, name = "Model 1", summary = FALSE, syntax = "")))
```

*ANCOVA - RLang05*

| Cases | Sum of Squares | df | Mean Square | F | p | $\eta^2_p$ |
|---|---|---|---|---|---|---|
| Group | 26.904 | 1 | 26.904 | 5.855 | 0.017 | 0.047 |
| Gender | 0.128 | 1 | 0.128 | 0.028 | 0.868 | $2.363\times10^{-4}$ |
| Age | 27.579 | 1 | 27.579 | 6.001 | 0.016 | 0.048 |
| Education | 16.642 | 1 | 16.642 | 3.621 | 0.059 | 0.030 |
| Handedness | 64.919 | 1 | 64.919 | 14.127 | $2.672\times10^{-4}$ | 0.107 |
| Sem_Dec_Acc | 7.112 | 1 | 7.112 | 1.548 | 0.216 | 0.013 |
| Residuals | 542.259 | 118 | 4.595 | | | |

*Note.* Type III Sum of Squares

#### Assumption Checks

*Test for Equality of Variances (Levene's)*

| F | df1 | df2 | p |
|---|---|---|---|
| 2.205 | 3.000 | 121.000 | 0.091 |

#### Marginal Means

*Marginal Means - Group*

| | | 95% CI for Mean Difference | | | | | |
|---|---|---|---|---|---|---|---|
| Group | Marginal Mean | Lower | Upper | SE | t | df | p |
| Control | -0.826 | -1.492 | -0.159 | 0.337 | -2.453 | 118 | 0.016 |
| Stroke | 0.279 | -0.243 | 0.800 | 0.263 | 1.059 | 118 | 0.292 |

### Supplementary Table 10- RLang06 ANCOVA comparing Stroke vs Control groups

```
jaspAnova::Ancova(
        version = "0.19.2",
        formula = RLang06 ~ Group.nominal + Gender + Age + Education + Handedness + 
Sem_Dec_Acc,
        covariates = list("Age", "Education", "Handedness", "Sem_Dec_Acc"),
        descriptivePlotErrorBar = TRUE,
        effectSizeEstimates = TRUE,
        effectSizeOmegaSquared = FALSE,
        effectSizePartialEtaSquared = TRUE,
        homogeneityTests = TRUE,
        marginalMeanComparedToZero = TRUE,
        marginalMeanTerms = ~ Group.nominal,
        restrictedModelComparison = "unconstrained",
        restrictedModels = list(list(informedHypothesisTest = FALSE, marginalMean = 
FALSE, name = "Model 1", summary = FALSE, syntax = "")))
```

*ANCOVA - RLang06*

| Cases | Sum of Squares | df | Mean Square | F | p | $\eta^2_p$ |
|---|---|---|---|---|---|---|
| Group | 0.392 | 1 | 0.392 | 0.372 | 0.543 | 0.003 |
| Gender | 2.194 | 1 | 2.194 | 2.082 | 0.152 | 0.017 |
| Age | 0.495 | 1 | 0.495 | 0.470 | 0.494 | 0.004 |
| Education | 8.442 | 1 | 8.442 | 8.009 | 0.005 | 0.064 |
| Handedness | 2.453 | 1 | 2.453 | 2.327 | 0.130 | 0.019 |
| Sem_Dec_Acc | 6.559 | 1 | 6.559 | 6.222 | 0.014 | 0.050 |
| Residuals | 124.379 | 118 | 1.054 | | | |

*Note.* Type III Sum of Squares

#### Assumption Checks

*Test for Equality of Variances (Levene's)*

| F | df1 | df2 | p |
|---|---|---|---|
| 0.950 | 3.000 | 121.000 | 0.419 |

#### Marginal Means

*Marginal Means - Group*

| | | 95% CI for Mean Difference | | | | | |
|---|---|---|---|---|---|---|---|
| Group | Marginal Mean | Lower | Upper | SE | t | df | p |
| Control | 0.657 | 0.337 | 0.976 | 0.161 | 4.074 | 118 | $8.423\times10^{-5}$ |
| Stroke | 0.790 | 0.540 | 1.040 | 0.126 | 6.267 | 118 | $6.231\times10^{-9}$ |

### Supplementary Table 11- RLang07 ANCOVA comparing Stroke vs Control groups

```
jaspAnova::Ancova(
        version = "0.19.2",
        formula = RLang07 ~ Group.nominal + Gender + Age + Education + Handedness + 
Sem_Dec_Acc,
        covariates = list("Age", "Education", "Handedness", "Sem_Dec_Acc"),
        descriptivePlotErrorBar = TRUE,
        effectSizeEstimates = TRUE,
        effectSizeOmegaSquared = FALSE,
        effectSizePartialEtaSquared = TRUE,
        homogeneityTests = TRUE,
        marginalMeanComparedToZero = TRUE,
        marginalMeanTerms = ~ Group.nominal,
        restrictedModelComparison = "unconstrained",
        restrictedModels = list(list(informedHypothesisTest = FALSE, marginalMean = 
FALSE, name = "Model 1", summary = FALSE, syntax = "")))
```

*ANCOVA - RLang07*

| Cases | Sum of Squares | df | Mean Square | F | p | $\eta^2_p$ |
|---|---|---|---|---|---|---|
| Group | 0.104 | 1 | 0.104 | 0.071 | 0.790 | $6.021\times10^{-4}$ |
| Gender | 1.197 | 1 | 1.197 | 0.819 | 0.367 | 0.007 |
| Age | 0.013 | 1 | 0.013 | 0.009 | 0.925 | $7.490\times10^{-5}$ |
| Education | 7.799 | 1 | 7.799 | 5.336 | 0.023 | 0.043 |
| Handedness | 3.386 | 1 | 3.386 | 2.317 | 0.131 | 0.019 |
| Sem_Dec_Acc | 11.528 | 1 | 11.528 | 7.887 | 0.006 | 0.063 |
| Residuals | 172.483 | 118 | 1.462 | | | |

*Note.* Type III Sum of Squares

#### Assumption Checks

*Test for Equality of Variances (Levene's)*

| F | df1 | df2 | p |
|---|---|---|---|
| 0.791 | 3.000 | 121.000 | 0.501 |

#### Marginal Means

*Marginal Means - Group*

| | | 95% CI for Mean Difference | | | | | |
|---|---|---|---|---|---|---|---|
| Group | Marginal Mean | Lower | Upper | SE | t | df | p |
| Control | 1.154 | 0.778 | 1.530 | 0.190 | 6.079 | 118 | $1.530\times10^{-8}$ |
| Stroke | 1.223 | 0.929 | 1.517 | 0.148 | 8.235 | 118 | $2.770\times10^{-13}$ |

### Supplementary Table 12- RLang08 ANCOVA comparing Stroke vs Control groups

```
jaspAnova::Ancova(
        version = "0.19.2",
        formula = RLang08 ~ Group.nominal + Gender + Age + Education + Handedness +
Sem_Dec_Acc,
        covariates = list("Age", "Education", "Handedness", "Sem_Dec_Acc"),
        descriptivePlotErrorBar = TRUE,
        effectSizeEstimates = TRUE,
        effectSizeOmegaSquared = FALSE,
        effectSizePartialEtaSquared = TRUE,
        homogeneityTests = TRUE,
        marginalMeanComparedToZero = TRUE,
        marginalMeanTerms = ~ Group.nominal,
        restrictedModelComparison = "unconstrained",
        restrictedModels = list(list(informedHypothesisTest = FALSE, marginalMean =
FALSE, name = "Model 1", summary = FALSE, syntax = "")))
```

*ANCOVA - RLang08*

| Cases | Sum of Squares | df | Mean Square | F | p | $\eta^2_p$ |
|---|---|---|---|---|---|---|
| Group | 0.009 | 1 | 0.009 | 0.004 | 0.952 | $3.071 \times 10^{-5}$ |
| Gender | 1.388 | 1 | 1.388 | 0.550 | 0.460 | 0.005 |
| Age | 0.516 | 1 | 0.516 | 0.205 | 0.652 | 0.002 |
| Education | 3.366 | 1 | 3.366 | 1.335 | 0.250 | 0.011 |
| Handedness | 0.104 | 1 | 0.104 | 0.041 | 0.839 | $3.505 \times 10^{-4}$ |
| Sem_Dec_Acc | 18.328 | 1 | 18.328 | 7.267 | 0.008 | 0.058 |
| Residuals | 297.611 | 118 | 2.522 | | | |

*Note.* Type III Sum of Squares

#### Assumption Checks

*Test for Equality of Variances (Levene's)*

| F | df1 | df2 | p |
|---|---|---|---|
| 1.631 | 3.000 | 121.000 | 0.186 |

#### Marginal Means

*Marginal Means - Group*

| | | 95% CI for Mean Difference | | | | | |
|---|---|---|---|---|---|---|---|
| Group | Marginal Mean | Lower | Upper | SE | t | df | p |
| Control | 1.301 | 0.807 | 1.795 | 0.249 | 5.218 | 118 | $7.827 \times 10^{-7}$ |
| Stroke | 1.322 | 0.935 | 1.708 | 0.195 | 6.777 | 118 | $5.120 \times 10^{-10}$ |

#### Supplementary Table 13- RLang09 ANCOVA comparing Stroke vs Control groups

```
jaspAnova::Ancova(
        version = "0.19.2",
        formula = RLang09 ~ Group.nominal + Gender + Age + Education + Handedness +
Sem_Dec_Acc,
        covariates = list("Age", "Education", "Handedness", "Sem_Dec_Acc"),
        descriptivePlotErrorBar = TRUE,
        effectSizeEstimates = TRUE,
        effectSizeOmegaSquared = FALSE,
        effectSizePartialEtaSquared = TRUE,
        homogeneityTests = TRUE,
        marginalMeanComparedToZero = TRUE,
        marginalMeanTerms = ~ Group.nominal,
        restrictedModelComparison = "unconstrained",
        restrictedModels = list(list(informedHypothesisTest = FALSE, marginalMean =
FALSE, name = "Model 1", summary = FALSE, syntax = "")))
```

*ANCOVA - RLang09*

| Cases | Sum of Squares | df | Mean Square | F | p | $\eta^2_p$ |
|---|---|---|---|---|---|---|
| Group | 1.033 | 1 | 1.033 | 0.371 | 0.544 | 0.003 |
| Gender | 0.194 | 1 | 0.194 | 0.070 | 0.793 | $5.888\times10^{-4}$ |
| Age | 16.951 | 1 | 16.951 | 6.081 | 0.015 | 0.049 |
| Education | 4.103 | 1 | 4.103 | 1.472 | 0.227 | 0.012 |
| Handedness | 10.711 | 1 | 10.711 | 3.843 | 0.052 | 0.032 |
| Sem_Dec_Acc | 1.974 | 1 | 1.974 | 0.708 | 0.402 | 0.006 |
| Residuals | 328.913 | 118 | 2.787 | | | |

*Note.* Type III Sum of Squares

##### Assumption Checks

*Test for Equality of Variances (Levene's)*

| F | df1 | df2 | p |
|---|---|---|---|
| 0.836 | 3.000 | 121.000 | 0.476 |

##### Marginal Means

*Marginal Means - Group*

| | | 95% CI for Mean Difference | | | | | |
|---|---|---|---|---|---|---|---|
| Group | Marginal Mean | Lower | Upper | SE | t | df | p |
| Control | 0.690 | 0.171 | 1.209 | 0.262 | 2.632 | 118 | 0.010 |
| Stroke | 0.906 | 0.500 | 1.312 | 0.205 | 4.421 | 118 | $2.197\times10^{-5}$ |

#### Supplementary Table 14- RLang10 ANCOVA comparing Stroke vs Control groups

```
jaspAnova::Ancova(
        version = "0.19.2",
        formula = RLang10 ~ Group.nominal + Gender + Age + Education + Handedness +
Sem_Dec_Acc,
        covariates = list("Age", "Education", "Handedness", "Sem_Dec_Acc"),
        descriptivePlotErrorBar = TRUE,
        effectSizeEstimates = TRUE,
        effectSizeOmegaSquared = FALSE,
        effectSizePartialEtaSquared = TRUE,
        homogeneityTests = TRUE,
        marginalMeanComparedToZero = TRUE,
        marginalMeanTerms = ~ Group.nominal,
        restrictedModelComparison = "unconstrained",
        restrictedModels = list(list(informedHypothesisTest = FALSE, marginalMean =
FALSE, name = "Model 1", summary = FALSE, syntax = "")))
```

*ANCOVA - RLang10*

| Cases | Sum of Squares | df | Mean Square | F | p | $\eta^2_p$ |
|---|---|---|---|---|---|---|
| Group | 5.922 | 1 | 5.922 | 1.585 | 0.211 | 0.013 |
| Gender | 3.470 | 1 | 3.470 | 0.929 | 0.337 | 0.008 |
| Age | 6.299 | 1 | 6.299 | 1.686 | 0.197 | 0.014 |
| Education | 7.569 | 1 | 7.569 | 2.026 | 0.157 | 0.017 |
| Handedness | 20.723 | 1 | 20.723 | 5.546 | 0.020 | 0.045 |
| Sem_Dec_Acc | 10.599 | 1 | 10.599 | 2.837 | 0.095 | 0.023 |
| Residuals | 440.890 | 118 | 3.736 | | | |

*Note.* Type III Sum of Squares

##### Assumption Checks

*Test for Equality of Variances (Levene's)*

| F | df1 | df2 | p |
|---|---|---|---|
| 0.843 | 3.000 | 121.000 | 0.473 |

##### Marginal Means

*Marginal Means - Group*

| | | 95% CI for Mean Difference | | | | | |
|---|---|---|---|---|---|---|---|
| Group | Marginal Mean | Lower | Upper | SE | t | df | p |
| Control | 0.026 | -0.575 | 0.627 | 0.304 | 0.086 | 118 | 0.931 |
| Stroke | 0.544 | 0.074 | 1.015 | 0.237 | 2.294 | 118 | 0.024 |

### Supplementary Table 15- RLang11 ANCOVA comparing Stroke vs Control groups

```
jaspAnova::Ancova(
        version = "0.19.2",
        formula = RLang11 ~ Group.nominal + Gender + Age + Education + Handedness + 
Sem_Dec_Acc,
        covariates = list("Age", "Education", "Handedness", "Sem_Dec_Acc"),
        descriptivePlotErrorBar = TRUE,
        effectSizeEstimates = TRUE,
        effectSizeOmegaSquared = FALSE,
        effectSizePartialEtaSquared = TRUE,
        homogeneityTests = TRUE,
        marginalMeanComparedToZero = TRUE,
        marginalMeanTerms = ~ Group.nominal,
        restrictedModelComparison = "unconstrained",
        restrictedModels = list(list(informedHypothesisTest = FALSE, marginalMean = 
FALSE, name = "Model 1", summary = FALSE, syntax = "")))
```

*ANCOVA - RLang11*

| Cases | Sum of Squares | df | Mean Square | F | p | $\eta^2_p$ |
|---|---|---|---|---|---|---|
| Group | 2.800 | 1 | 2.800 | 0.567 | 0.453 | 0.005 |
| Gender | 0.959 | 1 | 0.959 | 0.194 | 0.660 | 0.002 |
| Age | 0.944 | 1 | 0.944 | 0.191 | 0.663 | 0.002 |
| Education | 1.651 | 1 | 1.651 | 0.334 | 0.564 | 0.003 |
| Handedness | 12.983 | 1 | 12.983 | 2.630 | 0.108 | 0.022 |
| Sem_Dec_Acc | 10.597 | 1 | 10.597 | 2.146 | 0.146 | 0.018 |
| Residuals | 582.590 | 118 | 4.937 | | | |

*Note.* Type III Sum of Squares

#### Assumption Checks

*Test for Equality of Variances (Levene's)*

| F | df1 | df2 | p |
|---|---|---|---|
| 1.692 | 3.000 | 121.000 | 0.172 |

#### Marginal Means

*Marginal Means - Group*

| | | 95% CI for Mean Difference | | | | | |
|---|---|---|---|---|---|---|---|
| Group | Marginal Mean | Lower | Upper | SE | t | df | p |
| Control | -0.087 | -0.778 | 0.604 | 0.349 | -0.249 | 118 | 0.804 |
| Stroke | 0.270 | -0.271 | 0.810 | 0.273 | 0.988 | 118 | 0.325 |

### Supplementary Table 16- RSem01 ANCOVA comparing Stroke vs Control groups

```
jaspAnova::Ancova(
        version = "0.19.2",
        formula = RSem01 ~ Group.nominal + Gender + Age + Education + Handedness +
Sem_Dec_Acc,
        covariates = list("Age", "Education", "Handedness", "Sem_Dec_Acc"),
        descriptivePlotErrorBar = TRUE,
        effectSizeEstimates = TRUE,
        effectSizeOmegaSquared = FALSE,
        effectSizePartialEtaSquared = TRUE,
        homogeneityTests = TRUE,
        marginalMeanComparedToZero = TRUE,
        marginalMeanTerms = ~ Group.nominal,
        restrictedModelComparison = "unconstrained",
        restrictedModels = list(list(informedHypothesisTest = FALSE, marginalMean =
FALSE, name = "Model 1", summary = FALSE, syntax = "")))
```

*ANCOVA - RSem01*

| Cases | Sum of Squares | df | Mean Square | F | p | $\eta^2_p$ |
|---|---|---|---|---|---|---|
| Group | 1.199 | 1 | 1.199 | 1.647 | 0.202 | 0.014 |
| Gender | 0.087 | 1 | 0.087 | 0.119 | 0.731 | 0.001 |
| Age | 0.001 | 1 | 0.001 | 0.002 | 0.965 | $1.638\times10^{-5}$ |
| Education | 4.796 | 1 | 4.796 | 6.589 | 0.012 | 0.053 |
| Handedness | 4.641 | 1 | 4.641 | 6.375 | 0.013 | 0.051 |
| Sem_Dec_Acc | 3.396 | 1 | 3.396 | 4.665 | 0.033 | 0.038 |
| Residuals | 85.896 | 118 | 0.728 | | | |

*Note.* Type III Sum of Squares

#### Assumption Checks

*Test for Equality of Variances (Levene's)*

| F | df1 | df2 | p |
|---|---|---|---|
| 0.474 | 3.000 | 121.000 | 0.701 |

#### Marginal Means

*Marginal Means - Group*

| | | 95% CI for Mean Difference | | | | | |
|---|---|---|---|---|---|---|---|
| Group | Marginal Mean | Lower | Upper | SE | t | df | p |
| Control | 0.170 | -0.095 | 0.435 | 0.134 | 1.268 | 118 | 0.207 |
| Stroke | 0.403 | 0.196 | 0.611 | 0.105 | 3.848 | 118 | $1.941\times10^{-4}$ |

#### Supplementary Table 17- RSem02 ANCOVA comparing Stroke vs Control groups

```
jaspAnova::Ancova(
        version = "0.19.2",
        formula = RSem02 ~ Group.nominal + Gender + Age + Education + Handedness + 
Sem_Dec_Acc,
        covariates = list("Age", "Education", "Handedness", "Sem_Dec_Acc"),
        descriptivePlotErrorBar = TRUE,
        effectSizeEstimates = TRUE,
        effectSizeOmegaSquared = FALSE,
        effectSizePartialEtaSquared = TRUE,
        homogeneityTests = TRUE,
        marginalMeanComparedToZero = TRUE,
        marginalMeanTerms = ~ Group.nominal,
        restrictedModelComparison = "unconstrained",
        restrictedModels = list(list(informedHypothesisTest = FALSE, marginalMean = 
FALSE, name = "Model 1", summary = FALSE, syntax = "")))
```

*ANCOVA - RSem02*

| Cases | Sum of Squares | df | Mean Square | F | p | $\eta^2_p$ |
|---|---|---|---|---|---|---|
| Group | 4.589 | 1 | 4.589 | 1.732 | 0.191 | 0.014 |
| Gender | 1.650 | 1 | 1.650 | 0.623 | 0.431 | 0.005 |
| Age | 1.250 | 1 | 1.250 | 0.472 | 0.493 | 0.004 |
| Education | 13.180 | 1 | 13.180 | 4.976 | 0.028 | 0.040 |
| Handedness | 1.660 | 1 | 1.660 | 0.627 | 0.430 | 0.005 |
| Sem_Dec_Acc | 30.876 | 1 | 30.876 | 11.657 | $8.776\times10^{-4}$ | 0.090 |
| Residuals | 312.557 | 118 | 2.649 | | | |

*Note.* Type III Sum of Squares

##### Assumption Checks

*Test for Equality of Variances (Levene's)*

| F | df1 | df2 | p |
|---|---|---|---|
| 0.785 | 3.000 | 121.000 | 0.505 |

##### Marginal Means

*Marginal Means - Group*

| | | 95% CI for Mean Difference | | | | | |
|---|---|---|---|---|---|---|---|
| Group | Marginal Mean | Lower | Upper | SE | t | df | p |
| Control | 1.102 | 0.596 | 1.608 | 0.256 | 4.311 | 118 | $3.389\times10^{-5}$ |
| Stroke | 1.558 | 1.162 | 1.954 | 0.200 | 7.795 | 118 | $2.819\times10^{-12}$ |

#### Supplementary Table 18- RSem03 ANCOVA comparing Stroke vs Control groups

```
jaspAnova::Ancova(
        version = "0.19.2",
        formula = RSem03 ~ Group.nominal + Gender + Age + Education + Handedness + 
Sem_Dec_Acc,
        covariates = list("Age", "Education", "Handedness", "Sem_Dec_Acc"),
        descriptivePlotErrorBar = TRUE,
        effectSizeEstimates = TRUE,
        effectSizeOmegaSquared = FALSE,
        effectSizePartialEtaSquared = TRUE,
        homogeneityTests = TRUE,
        marginalMeanComparedToZero = TRUE,
        marginalMeanTerms = ~ Group.nominal,
        restrictedModelComparison = "unconstrained",
        restrictedModels = list(list(informedHypothesisTest = FALSE, marginalMean = 
FALSE, name = "Model 1", summary = FALSE, syntax = "")))
```

*ANCOVA - RSem03*

| Cases | Sum of Squares | df | Mean Square | F | p | $\eta^2_p$ |
|---|---|---|---|---|---|---|
| Group | 6.809 | 1 | 6.809 | 2.294 | 0.133 | 0.019 |
| Gender | 0.177 | 1 | 0.177 | 0.060 | 0.808 | $5.049 \times 10^{-4}$ |
| Age | 0.576 | 1 | 0.576 | 0.194 | 0.660 | 0.002 |
| Education | 17.440 | 1 | 17.440 | 5.875 | 0.017 | 0.047 |
| Handedness | 3.281 | 1 | 3.281 | 1.105 | 0.295 | 0.009 |
| Sem_Dec_Acc | 32.859 | 1 | 32.859 | 11.068 | 0.001 | 0.086 |
| Residuals | 350.309 | 118 | 2.969 | | | |

*Note.* Type III Sum of Squares

##### Assumption Checks

*Test for Equality of Variances (Levene's)*

| F | df1 | df2 | p |
|---|---|---|---|
| 0.908 | 3.000 | 121.000 | 0.439 |

##### Marginal Means

*Marginal Means - Group*

| | | 95% CI for Mean Difference | | | | | |
|---|---|---|---|---|---|---|---|
| Group | Marginal Mean | Lower | Upper | SE | t | df | p |
| Control | 0.956 | 0.421 | 1.492 | 0.271 | 3.535 | 118 | $5.829 \times 10^{-4}$ |
| Stroke | 1.512 | 1.093 | 1.931 | 0.212 | 7.147 | 118 | $7.972 \times 10^{-11}$ |

#### Supplementary Table 19- RSem04 ANCOVA comparing Stroke vs Control groups

```
jaspAnova::Ancova(
        version = "0.19.2",
        formula = RSem04 ~ Group.nominal + Gender + Age + Education + Handedness +
Sem_Dec_Acc,
        covariates = list("Age", "Education", "Handedness", "Sem_Dec_Acc"),
        descriptivePlotErrorBar = TRUE,
        effectSizeEstimates = TRUE,
        effectSizeOmegaSquared = FALSE,
        effectSizePartialEtaSquared = TRUE,
        homogeneityTests = TRUE,
        marginalMeanComparedToZero = TRUE,
        marginalMeanTerms = ~ Group.nominal,
        restrictedModelComparison = "unconstrained",
        restrictedModels = list(list(informedHypothesisTest = FALSE, marginalMean =
FALSE, name = "Model 1", summary = FALSE, syntax = "")))
```

*ANCOVA - RSem04*

| Cases | Sum of Squares | df | Mean Square | F | p | $\eta^2_p$ |
|---|---|---|---|---|---|---|
| Group | 17.284 | 1 | 17.284 | 2.588 | 0.110 | 0.021 |
| Gender | 11.563 | 1 | 11.563 | 1.731 | 0.191 | 0.014 |
| Age | 13.082 | 1 | 13.082 | 1.958 | 0.164 | 0.016 |
| Education | 14.465 | 1 | 14.465 | 2.165 | 0.144 | 0.018 |
| Handedness | 106.111 | 1 | 106.111 | 15.886 | $1.170\times10^{-4}$ | 0.119 |
| Sem_Dec_Acc | 4.603 | 1 | 4.603 | 0.689 | 0.408 | 0.006 |
| Residuals | 788.202 | 118 | 6.680 | | | |

*Note.* Type III Sum of Squares

##### Assumption Checks

*Test for Equality of Variances (Levene's)*

| F | df1 | df2 | p |
|---|---|---|---|
| 1.573 | 3.000 | 121.000 | 0.199 |

##### Marginal Means

*Marginal Means - Group*

| | | 95% CI for Mean Difference | | | | | |
|---|---|---|---|---|---|---|---|
| Group | Marginal Mean | Lower | Upper | SE | t | df | p |
| Control | -2.388 | -3.191 | -1.584 | 0.406 | -5.884 | 118 | $3.836\times10^{-8}$ |
| Stroke | -1.502 | -2.131 | -0.874 | 0.317 | -4.734 | 118 | $6.178\times10^{-6}$ |

### Supplementary Table 20- RSem05 ANCOVA comparing Stroke vs Control groups

```
jaspAnova::Ancova(
        version = "0.19.2",
        formula = RSem05 ~ Group.nominal + Gender + Age + Education + Handedness + 
Sem_Dec_Acc,
        covariates = list("Age", "Education", "Handedness", "Sem_Dec_Acc"),
        descriptivePlotErrorBar = TRUE,
        effectSizeEstimates = TRUE,
        effectSizeOmegaSquared = FALSE,
        effectSizePartialEtaSquared = TRUE,
        homogeneityTests = TRUE,
        marginalMeanComparedToZero = TRUE,
        marginalMeanTerms = ~ Group.nominal,
        restrictedModelComparison = "unconstrained",
        restrictedModels = list(list(informedHypothesisTest = FALSE, marginalMean = 
FALSE, name = "Model 1", summary = FALSE, syntax = "")))
```

*ANCOVA - RSem05*

| Cases | Sum of Squares | df | Mean Square | F | p | $\eta^2_p$ |
|---|---|---|---|---|---|---|
| Group | 2.393 | 1 | 2.393 | 0.480 | 0.490 | 0.004 |
| Gender | 7.567 | 1 | 7.567 | 1.519 | 0.220 | 0.013 |
| Age | 1.857 | 1 | 1.857 | 0.373 | 0.543 | 0.003 |
| Education | 7.637 | 1 | 7.637 | 1.533 | 0.218 | 0.013 |
| Handedness | 25.805 | 1 | 25.805 | 5.181 | 0.025 | 0.042 |
| Sem_Dec_Acc | 7.595 | 1 | 7.595 | 1.525 | 0.219 | 0.013 |
| Residuals | 587.704 | 118 | 4.981 | | | |

*Note.* Type III Sum of Squares

#### Assumption Checks

*Test for Equality of Variances (Levene's)*

| F | df1 | df2 | p |
|---|---|---|---|
| 3.880 | 3.000 | 121.000 | 0.011 |

#### Marginal Means

*Marginal Means - Group*

| | | 95% CI for Mean Difference | | | | | |
|---|---|---|---|---|---|---|---|
| Group | Marginal Mean | Lower | Upper | SE | t | df | p |
| Control | -1.188 | -1.882 | -0.494 | 0.350 | -3.389 | 118 | $9.538 \times 10^{-4}$ |
| Stroke | -0.858 | -1.401 | -0.316 | 0.274 | -3.132 | 118 | 0.002 |

### Supplementary Table 21- RSem06 ANCOVA comparing Stroke vs Control groups

```
jaspAnova::Ancova(
        version = "0.19.2",
        formula = RSem06 ~ Group.nominal + Gender + Age + Education + Handedness + 
Sem_Dec_Acc,
        covariates = list("Age", "Education", "Handedness", "Sem_Dec_Acc"),
        descriptivePlotErrorBar = TRUE,
        effectSizeEstimates = TRUE,
        effectSizeOmegaSquared = FALSE,
        effectSizePartialEtaSquared = TRUE,
        homogeneityTests = TRUE,
        marginalMeanComparedToZero = TRUE,
        marginalMeanTerms = ~ Group.nominal,
        restrictedModelComparison = "unconstrained",
        restrictedModels = list(list(informedHypothesisTest = FALSE, marginalMean = 
FALSE, name = "Model 1", summary = FALSE, syntax = "")))
```

*ANCOVA - RSem06*

| Cases | Sum of Squares | df | Mean Square | F | p | $\eta^2_p$ |
|---|---|---|---|---|---|---|
| Group | 4.965 | 1 | 4.965 | 1.832 | 0.178 | 0.015 |
| Gender | 0.916 | 1 | 0.916 | 0.338 | 0.562 | 0.003 |
| Age | 0.347 | 1 | 0.347 | 0.128 | 0.721 | 0.001 |
| Education | 14.587 | 1 | 14.587 | 5.381 | 0.022 | 0.044 |
| Handedness | 14.329 | 1 | 14.329 | 5.287 | 0.023 | 0.043 |
| Sem_Dec_Acc | 8.944 | 1 | 8.944 | 3.300 | 0.072 | 0.027 |
| Residuals | 319.840 | 118 | 2.711 | | | |

*Note.* Type III Sum of Squares

#### Assumption Checks

*Test for Equality of Variances (Levene's)*

| F | df1 | df2 | p |
|---|---|---|---|
| 0.685 | 3.000 | 121.000 | 0.563 |

#### Marginal Means

*Marginal Means - Group*

| | | 95% CI for Mean Difference | | | | | |
|---|---|---|---|---|---|---|---|
| Group | Marginal Mean | Lower | Upper | SE | t | df | p |
| Control | 0.430 | -0.082 | 0.942 | 0.259 | 1.663 | 118 | 0.099 |
| Stroke | 0.905 | 0.504 | 1.305 | 0.202 | 4.474 | 118 | $1.778 \times 10^{-5}$ |

#### Supplementary Table 22- RSem07 ANCOVA comparing Stroke vs Control groups

```
jaspAnova::Ancova(
        version = "0.19.2",
        formula = RSem07 ~ Group.nominal + Gender + Age + Education + Handedness +
Sem_Dec_Acc,
        covariates = list("Age", "Education", "Handedness", "Sem_Dec_Acc"),
        descriptivePlotErrorBar = TRUE,
        effectSizeEstimates = TRUE,
        effectSizeOmegaSquared = FALSE,
        effectSizePartialEtaSquared = TRUE,
        homogeneityTests = TRUE,
        marginalMeanComparedToZero = TRUE,
        marginalMeanTerms = ~ Group.nominal,
        restrictedModelComparison = "unconstrained",
        restrictedModels = list(list(informedHypothesisTest = FALSE, marginalMean =
FALSE, name = "Model 1", summary = FALSE, syntax = "")))
```

*ANCOVA - RSem07*

| Cases | Sum of Squares | df | Mean Square | F | p | $\eta^2_p$ |
|---|---|---|---|---|---|---|
| Group | 0.720 | 1 | 0.720 | 0.315 | 0.576 | 0.003 |
| Gender | 2.173 | 1 | 2.173 | 0.951 | 0.331 | 0.008 |
| Age | 0.145 | 1 | 0.145 | 0.064 | 0.801 | $5.381\times10^{-4}$ |
| Education | 1.598 | 1 | 1.598 | 0.699 | 0.405 | 0.006 |
| Handedness | 14.634 | 1 | 14.634 | 6.403 | 0.013 | 0.051 |
| Sem_Dec_Acc | 3.200 | 1 | 3.200 | 1.400 | 0.239 | 0.012 |
| Residuals | 269.681 | 118 | 2.285 | | | |

*Note.* Type III Sum of Squares

##### Assumption Checks

*Test for Equality of Variances (Levene's)*

| F | df1 | df2 | p |
|---|---|---|---|
| 1.793 | 3.000 | 121.000 | 0.152 |

##### Marginal Means

*Marginal Means - Group*

| | | 95% CI for Mean Difference | | | | | |
|---|---|---|---|---|---|---|---|
| Group | Marginal Mean | Lower | Upper | SE | t | df | p |
| Control | 0.385 | -0.085 | 0.855 | 0.237 | 1.622 | 118 | 0.108 |
| Stroke | 0.566 | 0.198 | 0.933 | 0.186 | 3.047 | 118 | 0.003 |

#### Supplementary Table 23- RSem08 ANCOVA comparing Stroke vs Control groups

```
jaspAnova::Ancova(
        version = "0.19.2",
        formula = RSem08 ~ Group.nominal + Gender + Age + Education + Handedness +
Sem_Dec_Acc,
        covariates = list("Age", "Education", "Handedness", "Sem_Dec_Acc"),
        descriptivePlotErrorBar = TRUE,
        effectSizeEstimates = TRUE,
        effectSizeOmegaSquared = FALSE,
        effectSizePartialEtaSquared = TRUE,
        homogeneityTests = TRUE,
        marginalMeanComparedToZero = TRUE,
        marginalMeanTerms = ~ Group.nominal,
        restrictedModelComparison = "unconstrained",
        restrictedModels = list(list(informedHypothesisTest = FALSE, marginalMean =
FALSE, name = "Model 1", summary = FALSE, syntax = "")))
```

*ANCOVA - RSem08*

| Cases | Sum of Squares | df | Mean Square | F | p | $\eta^2_p$ |
|---|---|---|---|---|---|---|
| Group | 6.871 | 1 | 6.871 | 2.352 | 0.128 | 0.020 |
| Gender | 12.667 | 1 | 12.667 | 4.336 | 0.039 | 0.035 |
| Age | 9.272 | 1 | 9.272 | 3.174 | 0.077 | 0.026 |
| Education | 1.963 | 1 | 1.963 | 0.672 | 0.414 | 0.006 |
| Handedness | 46.813 | 1 | 46.813 | 16.025 | $1.096\times10^{-4}$ | 0.120 |
| Sem_Dec_Acc | 0.608 | 1 | 0.608 | 0.208 | 0.649 | 0.002 |
| Residuals | 344.699 | 118 | 2.921 | | | |

*Note.* Type III Sum of Squares

##### Assumption Checks

*Test for Equality of Variances (Levene's)*

| F | df1 | df2 | p |
|---|---|---|---|
| 1.521 | 3.000 | 121.000 | 0.213 |

##### Marginal Means

*Marginal Means - Group*

| | | 95% CI for Mean Difference | | | | | |
|---|---|---|---|---|---|---|---|
| Group | Marginal Mean | Lower | Upper | SE | t | df | p |
| Control | 0.658 | 0.127 | 1.190 | 0.268 | 2.453 | 118 | 0.016 |
| Stroke | 1.216 | 0.801 | 1.632 | 0.210 | 5.796 | 118 | $5.776\times10^{-8}$ |

### Supplementary Table 24- RSem09 ANCOVA comparing Stroke vs Control groups

```
jaspAnova::Ancova(
        version = "0.19.2",
        formula = RSem09 ~ Group.nominal + Gender + Age + Education + Handedness + 
Sem_Dec_Acc,
        covariates = list("Age", "Education", "Handedness", "Sem_Dec_Acc"),
        descriptivePlotErrorBar = TRUE,
        effectSizeEstimates = TRUE,
        effectSizeOmegaSquared = FALSE,
        effectSizePartialEtaSquared = TRUE,
        homogeneityTests = TRUE,
        marginalMeanComparedToZero = TRUE,
        marginalMeanTerms = ~ Group.nominal,
        restrictedModelComparison = "unconstrained",
        restrictedModels = list(list(informedHypothesisTest = FALSE, marginalMean = 
FALSE, name = "Model 1", summary = FALSE, syntax = "")))
```

*ANCOVA - RSem09*

| Cases | Sum of Squares | df | Mean Square | F | p | $\eta^2_p$ |
|---|---|---|---|---|---|---|
| Group | 0.073 | 1 | 0.073 | 0.154 | 0.695 | 0.001 |
| Gender | 0.506 | 1 | 0.506 | 1.070 | 0.303 | 0.009 |
| Age | 1.666 | 1 | 1.666 | 3.525 | 0.063 | 0.029 |
| Education | 3.903 | 1 | 3.903 | 8.258 | 0.005 | 0.065 |
| Handedness | 0.396 | 1 | 0.396 | 0.837 | 0.362 | 0.007 |
| Sem_Dec_Acc | 1.828 | 1 | 1.828 | 3.869 | 0.052 | 0.032 |
| Residuals | 55.764 | 118 | 0.473 | | | |

*Note.* Type III Sum of Squares

#### Assumption Checks

*Test for Equality of Variances (Levene's)*

| F | df1 | df2 | p |
|---|---|---|---|
| 0.418 | 3.000 | 121.000 | 0.741 |

#### Marginal Means

*Marginal Means - Group*

| | | 95% CI for Mean Difference | | | | | |
|---|---|---|---|---|---|---|---|
| Group | Marginal Mean | Lower | Upper | SE | t | df | p |
| Control | 0.617 | 0.403 | 0.831 | 0.108 | 5.716 | 118 | $8.351 \times 10^{-8}$ |
| Stroke | 0.674 | 0.507 | 0.842 | 0.084 | 7.989 | 118 | $1.016 \times 10^{-12}$ |

#### Supplementary Table 25- RSem10 ANCOVA comparing Stroke vs Control groups

```
jaspAnova::Ancova(
        version = "0.19.2",
        formula = RSem10 ~ Group.nominal + Gender + Age + Education + Handedness + 
Sem_Dec_Acc,
        covariates = list("Age", "Education", "Handedness", "Sem_Dec_Acc"),
        descriptivePlotErrorBar = TRUE,
        effectSizeEstimates = TRUE,
        effectSizeOmegaSquared = FALSE,
        effectSizePartialEtaSquared = TRUE,
        homogeneityTests = TRUE,
        marginalMeanComparedToZero = TRUE,
        marginalMeanTerms = ~ Group.nominal,
        restrictedModelComparison = "unconstrained",
        restrictedModels = list(list(informedHypothesisTest = FALSE, marginalMean = 
FALSE, name = "Model 1", summary = FALSE, syntax = "")))
```

*ANCOVA - RSem10*

| Cases | Sum of Squares | df | Mean Square | F | p | $\eta^2_p$ |
|---|---|---|---|---|---|---|
| Group | 0.215 | 1 | 0.215 | 0.331 | 0.566 | 0.003 |
| Gender | 0.036 | 1 | 0.036 | 0.055 | 0.815 | $4.635\times10^{-4}$ |
| Age | 0.238 | 1 | 0.238 | 0.367 | 0.546 | 0.003 |
| Education | 0.289 | 1 | 0.289 | 0.445 | 0.506 | 0.004 |
| Handedness | 2.483 | 1 | 2.483 | 3.826 | 0.053 | 0.031 |
| Sem_Dec_Acc | $2.943\times10^{-5}$ | 1 | $2.943\times10^{-5}$ | $4.535\times10^{-5}$ | 0.995 | $3.843\times10^{-7}$ |
| Residuals | 76.588 | 118 | 0.649 | | | |

*Note.* Type III Sum of Squares

##### Assumption Checks

*Test for Equality of Variances (Levene's)*

| F | df1 | df2 | p |
|---|---|---|---|
| 1.622 | 3.000 | 121.000 | 0.188 |

##### Marginal Means

*Marginal Means - Group*

| | | 95% CI for Mean Difference | | | | | |
|---|---|---|---|---|---|---|---|
| Group | Marginal Mean | Lower | Upper | SE | t | df | p |
| Control | 0.194 | -0.056 | 0.445 | 0.127 | 1.537 | 118 | 0.127 |
| Stroke | 0.293 | 0.097 | 0.489 | 0.099 | 2.964 | 118 | 0.004 |

#### Supplementary Table 26- RSem11 ANCOVA comparing Stroke vs Control groups

```
jaspAnova::Ancova(
        version = "0.19.2",
        formula = RSem11 ~ Group.nominal + Gender + Age + Education + Handedness + 
Sem_Dec_Acc,
        covariates = list("Age", "Education", "Handedness", "Sem_Dec_Acc"),
        descriptivePlotErrorBar = TRUE,
        effectSizeEstimates = TRUE,
        effectSizeOmegaSquared = FALSE,
        effectSizePartialEtaSquared = TRUE,
        homogeneityTests = TRUE,
        marginalMeanComparedToZero = TRUE,
        marginalMeanTerms = ~ Group.nominal,
        restrictedModelComparison = "unconstrained",
        restrictedModels = list(list(informedHypothesisTest = FALSE, marginalMean = 
FALSE, name = "Model 1", summary = FALSE, syntax = "")))
```

*ANCOVA - RSem11*

| Cases | Sum of Squares | df | Mean Square | F | p | $\eta^2_p$ |
|---|---|---|---|---|---|---|
| Group | 0.008 | 1 | 0.008 | 0.009 | 0.927 | $7.216\times10^{-5}$ |
| Gender | 0.600 | 1 | 0.600 | 0.636 | 0.427 | 0.005 |
| Age | 0.080 | 1 | 0.080 | 0.085 | 0.771 | $7.203\times10^{-4}$ |
| Education | 4.247 | 1 | 4.247 | 4.500 | 0.036 | 0.037 |
| Handedness | 0.286 | 1 | 0.286 | 0.304 | 0.583 | 0.003 |
| Sem_Dec_Acc | 3.517 | 1 | 3.517 | 3.726 | 0.056 | 0.031 |
| Residuals | 111.374 | 118 | 0.944 | | | |

*Note.* Type III Sum of Squares

##### Assumption Checks

*Test for Equality of Variances (Levene's)*

| F | df1 | df2 | p |
|---|---|---|---|
| 0.065 | 3.000 | 121.000 | 0.978 |

##### Marginal Means

*Marginal Means - Group*

| | | 95% CI for Mean Difference | | | | | |
|---|---|---|---|---|---|---|---|
| Group | Marginal Mean | Lower | Upper | SE | t | df | p |
| Control | 0.324 | 0.022 | 0.627 | 0.153 | 2.127 | 118 | 0.036 |
| Stroke | 0.344 | 0.107 | 0.580 | 0.119 | 2.880 | 118 | 0.005 |

### Supplementary Table 27- RSem12 ANCOVA comparing Stroke vs Control groups

```
jaspAnova::Ancova(
        version = "0.19.2",
        formula = RSem12 ~ Group.nominal + Gender + Age + Education + Handedness +
Sem_Dec_Acc,
        covariates = list("Age", "Education", "Handedness", "Sem_Dec_Acc"),
        descriptivePlotErrorBar = TRUE,
        effectSizeEstimates = TRUE,
        effectSizeOmegaSquared = FALSE,
        effectSizePartialEtaSquared = TRUE,
        homogeneityTests = TRUE,
        marginalMeanComparedToZero = TRUE,
        marginalMeanTerms = ~ Group.nominal,
        restrictedModelComparison = "unconstrained",
        restrictedModels = list(list(informedHypothesisTest = FALSE, marginalMean =
FALSE, name = "Model 1", summary = FALSE, syntax = "")))
```

*ANCOVA - RSem12*

| Cases | Sum of Squares | df | Mean Square | F | p | $\eta^2_p$ |
|---|---|---|---|---|---|---|
| Group | 2.345 | 1 | 2.345 | 0.161 | 0.689 | 0.001 |
| Gender | 22.253 | 1 | 22.253 | 1.529 | 0.219 | 0.013 |
| Age | 18.795 | 1 | 18.795 | 1.291 | 0.258 | 0.011 |
| Education | 4.711 | 1 | 4.711 | 0.324 | 0.571 | 0.003 |
| Handedness | 10.771 | 1 | 10.771 | 0.740 | 0.391 | 0.006 |
| Sem_Dec_Acc | 173.847 | 1 | 173.847 | 11.941 | $7.639\times10^{-4}$ | 0.092 |
| Residuals | 1717.952 | 118 | 14.559 | | | |

*Note.* Type III Sum of Squares

#### Assumption Checks

*Test for Equality of Variances (Levene's)*

| F | df1 | df2 | p |
|---|---|---|---|
| 4.141 | 3.000 | 121.000 | 0.008 |

#### Marginal Means

*Marginal Means - Group*

| | | 95% CI for Mean Difference | | | | | |
|---|---|---|---|---|---|---|---|
| Group | Marginal Mean | Lower | Upper | SE | t | df | p |
| Control | -3.049 | -4.236 | -1.863 | 0.599 | -5.089 | 118 | $1.374\times10^{-6}$ |
| Stroke | -3.375 | -4.303 | -2.447 | 0.469 | -7.203 | 118 | $5.976\times10^{-11}$ |

#### Supplementary Table 28- RSem13 ANCOVA comparing Stroke vs Control groups

```
jaspAnova::Ancova(
        version = "0.19.2",
        formula = RSem13 ~ Group.nominal + Gender + Age + Education + Handedness +
Sem_Dec_Acc,
        covariates = list("Age", "Education", "Handedness", "Sem_Dec_Acc"),
        descriptivePlotErrorBar = TRUE,
        effectSizeEstimates = TRUE,
        effectSizeOmegaSquared = FALSE,
        effectSizePartialEtaSquared = TRUE,
        homogeneityTests = TRUE,
        marginalMeanComparedToZero = TRUE,
        marginalMeanTerms = ~ Group.nominal,
        restrictedModelComparison = "unconstrained",
        restrictedModels = list(list(informedHypothesisTest = FALSE, marginalMean =
FALSE, name = "Model 1", summary = FALSE, syntax = "")))
```

*ANCOVA - RSem13*

| Cases | Sum of Squares | df | Mean Square | F | p | $\eta^2_p$ |
|---|---|---|---|---|---|---|
| Group | 2.186 | 1 | 2.186 | 0.982 | 0.324 | 0.008 |
| Gender | 1.736 | 1 | 1.736 | 0.780 | 0.379 | 0.007 |
| Age | 9.113 | 1 | 9.113 | 4.094 | 0.045 | 0.034 |
| Education | 11.112 | 1 | 11.112 | 4.991 | 0.027 | 0.041 |
| Handedness | 7.175 | 1 | 7.175 | 3.223 | 0.075 | 0.027 |
| Sem_Dec_Acc | 0.019 | 1 | 0.019 | 0.009 | 0.926 | $7.409\times10^{-5}$ |
| Residuals | 262.685 | 118 | 2.226 | | | |

*Note.* Type III Sum of Squares

##### Assumption Checks

*Test for Equality of Variances (Levene's)*

| F | df1 | df2 | p |
|---|---|---|---|
| 1.138 | 3.000 | 121.000 | 0.337 |

##### Marginal Means

*Marginal Means - Group*

| | | 95% CI for Mean Difference | | | | | |
|---|---|---|---|---|---|---|---|
| Group | Marginal Mean | Lower | Upper | SE | t | df | p |
| Control | 0.648 | 0.184 | 1.112 | 0.234 | 2.764 | 118 | 0.007 |
| Stroke | 0.962 | 0.600 | 1.325 | 0.183 | 5.253 | 118 | $6.724\times10^{-7}$ |

### Supplementary Table 29- RSem14 ANCOVA comparing Stroke vs Control groups

```
jaspAnova::Ancova(
        version = "0.19.2",
        formula = RSem14 ~ Group.nominal + Gender + Age + Education + Handedness + 
Sem_Dec_Acc,
        covariates = list("Age", "Education", "Handedness", "Sem_Dec_Acc"),
        descriptivePlotErrorBar = TRUE,
        effectSizeEstimates = TRUE,
        effectSizeOmegaSquared = FALSE,
        effectSizePartialEtaSquared = TRUE,
        homogeneityTests = TRUE,
        marginalMeanComparedToZero = TRUE,
        marginalMeanTerms = ~ Group.nominal,
        restrictedModelComparison = "unconstrained",
        restrictedModels = list(list(informedHypothesisTest = FALSE, marginalMean = 
FALSE, name = "Model 1", summary = FALSE, syntax = "")))
```

*ANCOVA - RSem14*

| Cases | Sum of Squares | df | Mean Square | F | p | $\eta^2_p$ |
|---|---|---|---|---|---|---|
| Group | 0.255 | 1 | 0.255 | 0.065 | 0.799 | $5.531\times10^{-4}$ |
| Gender | $3.946\times10^{-4}$ | 1 | $3.946\times10^{-4}$ | $1.011\times10^{-4}$ | 0.992 | $8.568\times10^{-7}$ |
| Age | 6.717 | 1 | 6.717 | 1.721 | 0.192 | 0.014 |
| Education | 3.125 | 1 | 3.125 | 0.801 | 0.373 | 0.007 |
| Handedness | 6.839 | 1 | 6.839 | 1.752 | 0.188 | 0.015 |
| Sem_Dec_Acc | 11.934 | 1 | 11.934 | 3.057 | 0.083 | 0.025 |
| Residuals | 460.593 | 118 | 3.903 | | | |

*Note.* Type III Sum of Squares

#### Assumption Checks

*Test for Equality of Variances (Levene's)*

| F | df1 | df2 | p |
|---|---|---|---|
| 2.548 | 3.000 | 121.000 | 0.059 |

#### Marginal Means

*Marginal Means - Group*

| | | 95% CI for Mean Difference | | | | | |
|---|---|---|---|---|---|---|---|
| Group | Marginal Mean | Lower | Upper | SE | t | df | p |
| Control | 0.783 | 0.168 | 1.397 | 0.310 | 2.523 | 118 | 0.013 |
| Stroke | 0.675 | 0.195 | 1.156 | 0.243 | 2.783 | 118 | 0.006 |

### Supplementary Table 30- Mixed-effects model examining effects of lesion size and time-since-stroke on RH Language Network activity

```
jaspMixedModels::MixedModelsLMM(
        version = "0.19.2",
        formula = RH_Activity ~ LogLesionSize + LogTimeSinceStroke + Age + Education +
Handedness + Sem_Dec_Acc + Gender + (1 + LogTimeSinceStroke | Node) + (1 | Participant),
        contrasts = NULL,
        fixedEffectEstimate = TRUE,
        modelSummary = TRUE,
        trendsContrasts = NULL)
```

*ANOVA Summary*

| Effect | df | F | p |
|---|---|---|---|
| LogLesionSize | 1, 68.00 | 0.039 | 0.845 |
| LogTimeSinceStroke | 1, 64.38 | 4.425 | 0.039 |
| Age | 1, 68.00 | 7.985 | 0.006 |
| Education | 1, 68.00 | 4.644 | 0.035 |
| Handedness | 1, 68.00 | 5.992 | 0.017 |
| Sem_Dec_Acc | 1, 68.00 | 1.847 | 0.179 |
| Gender | 1, 68.00 | 0.081 | 0.777 |

*Note.* Model terms tested with Satterthwaite testMethod.
*Note.* The following variables are used as random effects grouping factors: 'Node', 'Participant'.
*Note.* Type III Sum of Squares

#### Model summary

*Fit statistics*

| Deviance (REML) | log Lik. | df | AIC | BIC |
|---|---|---|---|---|
| 2094.228 | -1047.114 | 13 | 2120.228 | 2181.701 |

*Note.* The model was fitted using restricted maximum likelihood. Please note that models with different fixed effects cannot be compared when REML is used. To use ML, switch 'Test method' to 'Likelihood ratio tests'.

*Sample sizes*

| | Levels of RE grouping factors | |
|---|---|---|
| Observations | Participant | Node |
| 836 | 76 | 11 |

*Fixed Effects Estimates*

| Term | Estimate | SE | df | t | p |
|---|---|---|---|---|---|
| Intercept | -1.356 | 1.410 | 68.868 | -0.962 | 0.339 |
| LogLesionSize | -0.026 | 0.130 | 68.001 | -0.196 | 0.845 |
| LogTimeSinceStroke | 0.438 | 0.208 | 64.384 | 2.104 | 0.039 |
| Age | -0.021 | 0.008 | 68.001 | -2.826 | 0.006 |
| Education | 0.065 | 0.030 | 68.001 | 2.155 | 0.035 |
| Handedness | -0.004 | 0.002 | 68.001 | -2.448 | 0.017 |
| Sem_Dec_Acc | 1.774 | 1.306 | 68.001 | 1.359 | 0.179 |
| Gender (1) | -0.023 | 0.081 | 68.001 | -0.285 | 0.777 |

*Note.* The intercept corresponds to the (unweighted) grand mean; for each factor with k levels, k - 1 parameters are estimated with sum contrast coding. Consequently, the estimates cannot be directly mapped to factor levels. Use estimated marginal means for obtaining estimates for each factor level/design cell or their differences.

#### Supplementary Table 31- Mixed-effects model examining effects of nodewise homotopic LH lesions and LH activity on RH Language Network activity

*ANOVA Summary*

| Effect | df | F | p |
|---|---|---|---|
| LesionLoad | 1, 13.43 | 21.388 | $4.379\times10^{-4}$ |
| LH_Activity | 1, 44.22 | 42.640 | $5.487\times10^{-8}$ |
| LogLesionSize | 1, 70.59 | 0.041 | 0.840 |
| LogTimeSinceStroke | 1, 65.22 | 1.959 | 0.166 |
| Sem_Dec_Acc | 1, 68.39 | 0.347 | 0.558 |
| Age | 1, 67.95 | 5.732 | 0.019 |
| Education | 1, 68.46 | 2.417 | 0.125 |
| Handedness | 1, 67.39 | 6.162 | 0.016 |
| Gender | 1, 67.98 | 0.015 | 0.903 |

*Note.* Model terms tested with Satterthwaite testMethod.
*Note.* The following variables are used as random effects grouping factors: 'Node', 'Participant'.
*Note.* Type III Sum of Squares

#### Model summary

*Fit statistics*

| Deviance (REML) | log Lik. | df | AIC | BIC |
|---|---|---|---|---|
| 2049.499 | -1024.750 | 20 | 2089.499 | 2184.072 |

*Note.* The model was fitted using restricted maximum likelihood. Please note that models with different fixed effects cannot be compared when REML is used. To use ML, switch 'Test method' to 'Likelihood ratio tests'.

*Sample sizes*

| | Levels of RE grouping factors | |
|---|---|---|
| Observations | Participant | Node |
| 836 | 76 | 11 |

*Fixed Effects Estimates*

| Term | Estimate | SE | df | t | p |
|---|---|---|---|---|---|
| Intercept | -0.162 | 1.379 | 69.708 | -0.117 | 0.907 |
| LesionLoad | 0.637 | 0.138 | 13.427 | 4.625 | $4.379\times10^{-4}$ |
| LH_Activity | 0.276 | 0.042 | 44.217 | 6.530 | $5.487\times10^{-8}$ |
| LogLesionSize | -0.026 | 0.128 | 70.594 | -0.203 | 0.840 |
| LogTimeSinceStroke | 0.282 | 0.202 | 65.224 | 1.400 | 0.166 |
| Sem_Dec_Acc | 0.758 | 1.286 | 68.393 | 0.589 | 0.558 |
| Age | -0.018 | 0.007 | 67.945 | -2.394 | 0.019 |
| Education | 0.046 | 0.030 | 68.463 | 1.555 | 0.125 |
| Handedness | -0.004 | 0.001 | 67.390 | -2.482 | 0.016 |
| Gender (1) | 0.010 | 0.079 | 67.984 | 0.122 | 0.903 |

*Note.* The intercept corresponds to the (unweighted) grand mean; for each factor with k levels, k - 1 parameters are estimated with sum contrast coding. Consequently, the estimates cannot be directly mapped to factor levels. Use estimated marginal means for obtaining estimates for each factor level/design cell or their differences.

#### Supplementary Table 32- Mixed-effects model examining effects of nodewise LH lesions on residual LH Language Network activity

```
jaspMixedModels::MixedModelsLMM(
        version = "0.19.2",
        formula = LH_Activity ~ LesionLoad + LogLesionSize + LogTimeSinceStroke + 
Sem_Dec_Acc + Age + Education + Handedness + Gender + (1 + LesionLoad + Sem_Dec_Acc | 
Node) + (1 + LesionLoad | Participant),
        contrasts = NULL,
        fixedEffectEstimate = TRUE,
        modelSummary = TRUE,
        trendsContrasts = NULL)
```

*ANOVA Summary*

| Effect | df | F | p |
|---|---|---|---|
| LesionLoad | 1, 15.47 | 31.204 | $4.639\times10^{-5}$ |
| LogLesionSize | 1, 82.82 | 3.673 | 0.059 |
| LogTimeSinceStroke | 1, 43.86 | 10.048 | 0.003 |
| Sem_Dec_Acc | 1, 28.36 | 14.039 | $8.128\times10^{-4}$ |
| Age | 1, 33.85 | 0.290 | 0.594 |
| Education | 1, 52.56 | 5.089 | 0.028 |
| Handedness | 1, 38.43 | 0.009 | 0.923 |
| Gender | 1, 38.29 | 2.961 | 0.093 |

*Note.* Model terms tested with Satterthwaite testMethod.
*Note.* The following variables are used as random effects grouping factors: 'Node', 'Participant'.
*Note.* Type III Sum of Squares

#### Model summary

*Fit statistics*

| Deviance (REML) | log Lik. | df | AIC | BIC |
|---|---|---|---|---|
| 1698.487 | -849.243 | 19 | 1736.487 | 1826.331 |

*Note.* The model was fitted using restricted maximum likelihood. Please note that models with different fixed effects cannot be compared when REML is used. To use ML, switch 'Test method' to 'Likelihood ratio tests'.

*Sample sizes*

| | Levels of RE grouping factors | |
|---|---|---|
| Observations | Participant | Node |
| 836 | 76 | 11 |

*Fixed Effects Estimates*

| Term | Estimate | SE | df | t | p |
|---|---|---|---|---|---|
| Intercept | -3.096 | 0.943 | 58.381 | -3.285 | 0.002 |
| LesionLoad | -1.143 | 0.205 | 15.469 | -5.586 | $4.639\times10^{-5}$ |
| LogLesionSize | -0.193 | 0.101 | 82.819 | -1.916 | 0.059 |
| LogTimeSinceStroke | 0.383 | 0.121 | 43.859 | 3.170 | 0.003 |
| Sem_Dec_Acc | 3.044 | 0.812 | 28.357 | 3.747 | $8.128\times10^{-4}$ |
| Age | -0.002 | 0.004 | 33.849 | -0.539 | 0.594 |
| Education | 0.042 | 0.018 | 52.561 | 2.256 | 0.028 |
| Handedness | $-8.844\times10^{-5}$ | $9.104\times10^{-4}$ | 38.427 | -0.097 | 0.923 |

*Fit statistics*

| Deviance (REML) | log Lik. | df | AIC | | BIC |
|---|---|---|---|---|---|
| Gender (1) | -0.081 | 0.047 | 38.290 | -1.721 | 0.093 |

#### Supplementary Table 33- Mixed-effects model examining effects of nodewise homotopic LH lesions on RH Language Network activity

```
jaspMixedModels::MixedModelsLMM(
        version = "0.19.2",
        formula = RH_Activity ~ LesionLoad + LogLesionSize + LogTimeSinceStroke + 
Sem_Dec_Acc + Age + Education + Handedness + Gender + (1 + LogLesionSize | Node) + (1 + 
LesionLoad | Participant),
        contrasts = NULL,
        fixedEffectEstimate = TRUE,
        modelSummary = TRUE,
        trendsContrasts = NULL)
```

*ANOVA Summary*

| Effect | df | F | p |
|---|---|---|---|
| LesionLoad | 1, 42.88 | 5.194 | 0.028 |
| LogLesionSize | 1, 62.89 | 0.255 | 0.615 |
| LogTimeSinceStroke | 1, 68.59 | 5.280 | 0.025 |
| Sem_Dec_Acc | 1, 69.09 | 4.015 | 0.049 |
| Age | 1, 71.14 | 6.248 | 0.015 |
| Education | 1, 68.23 | 3.498 | 0.066 |
| Handedness | 1, 68.87 | 5.252 | 0.025 |
| Gender | 1, 68.45 | 0.003 | 0.955 |

*Note.* Model terms tested with Satterthwaite testMethod.
*Note.* The following variables are used as random effects grouping factors: 'Node', 'Participant'.
*Note.* Type III Sum of Squares

#### Model summary

*Fit statistics*

| Deviance (REML) | log Lik. | df | AIC | BIC |
|---|---|---|---|---|
| 2077.928 | -1038.964 | 16 | 2109.928 | 2185.586 |

*Note.* The model was fitted using restricted maximum likelihood. Please note that models with different fixed effects cannot be compared when REML is used. To use ML, switch 'Test method' to 'Likelihood ratio tests'.

*Sample sizes*

| | Levels of RE grouping factors | |
|---|---|---|
| Observations | Participant | Node |
| 836 | 76 | 11 |

*Fixed Effects Estimates*

| Term | Estimate | SE | df | t | p |
|---|---|---|---|---|---|
| Intercept | -1.854 | 1.322 | 67.308 | -1.402 | 0.166 |
| LesionLoad | 0.317 | 0.139 | 42.877 | 2.279 | 0.028 |
| LogLesionSize | -0.064 | 0.126 | 62.887 | -0.505 | 0.615 |
| LogTimeSinceStroke | 0.442 | 0.192 | 68.589 | 2.298 | 0.025 |
| Sem_Dec_Acc | 2.481 | 1.238 | 69.091 | 2.004 | 0.049 |
| Age | -0.018 | 0.007 | 71.136 | -2.500 | 0.015 |
| Education | 0.053 | 0.029 | 68.233 | 1.870 | 0.066 |
| Handedness | -0.003 | 0.001 | 68.871 | -2.292 | 0.025 |
| Gender (1) | 0.004 | 0.077 | 68.450 | 0.056 | 0.955 |

*Note.* The intercept corresponds to the (unweighted) grand mean; for each factor with k levels, k - 1 parameters are estimated with sum contrast coding. Consequently, the estimates cannot be directly mapped to factor levels. Use estimated marginal means for obtaining estimates for each factor level/design cell or their differences.

#### Supplementary Table 34- Mixed-effects model examining effects of nodewise homotopic LH activity on RH Language Network activity in the Stroke group

```
jaspMixedModels::MixedModelsLMM(
        version = "0.19.2",
        formula = RH_Activity ~ LH_Activity + LogLesionSize + LogTimeSinceStroke + 
Sem_Dec_Acc + Age + Education + Handedness + Gender + (1 + LH_Activity + Sem_Dec_Acc | 
Node) + (1 + LH_Activity | Participant),
        contrasts = NULL,
        fixedEffectEstimate = TRUE,
        modelSummary = TRUE,
        trendsContrasts = NULL)
```

*ANOVA Summary*

| Effect | df | F | p |
|---|---|---|---|
| LH_Activity | 1, 29.93 | 16.745 | $2.974 \times 10^{-4}$ |
| LogLesionSize | 1, 68.13 | 0.057 | 0.813 |
| LogTimeSinceStroke | 1, 67.53 | 2.880 | 0.094 |
| Sem_Dec_Acc | 1, 67.15 | 0.482 | 0.490 |
| Age | 1, 68.99 | 6.691 | 0.012 |
| Education | 1, 69.18 | 3.314 | 0.073 |
| Handedness | 1, 66.99 | 5.517 | 0.022 |
| Gender | 1, 68.41 | 0.001 | 0.973 |

*Note.* Model terms tested with Satterthwaite testMethod.
*Note.* The following variables are used as random effects grouping factors: 'Node', 'Participant'.
*Note.* Type III Sum of Squares

##### Model summary

*Fit statistics*

| Deviance (REML) | log Lik. | df | AIC | BIC |
|---|---|---|---|---|
| 2063.903 | -1031.951 | 19 | 2101.903 | 2191.747 |

*Note.* The model was fitted using restricted maximum likelihood. Please note that models with different fixed effects cannot be compared when REML is used. To use ML, switch 'Test method' to 'Likelihood ratio tests'.

*Sample sizes*

| | Levels of RE grouping factors | |
|---|---|---|
| Observations | Participant | Node |
| 836 | 76 | 11 |

*Fixed Effects Estimates*

| Term | Estimate | SE | df | t | p |
|---|---|---|---|---|---|
| Intercept | -0.650 | 1.431 | 69.452 | -0.454 | 0.651 |
| LH_Activity | 0.184 | 0.045 | 29.926 | 4.092 | $2.974 \times 10^{-4}$ |
| LogLesionSize | 0.031 | 0.129 | 68.129 | 0.238 | 0.813 |
| LogTimeSinceStroke | 0.342 | 0.202 | 67.534 | 1.697 | 0.094 |
| Sem_Dec_Acc | 0.938 | 1.352 | 67.152 | 0.694 | 0.490 |
| Age | -0.020 | 0.008 | 68.993 | -2.587 | 0.012 |
| Education | 0.055 | 0.030 | 69.184 | 1.820 | 0.073 |
| Handedness | -0.004 | 0.001 | 66.990 | -2.349 | 0.022 |
| Gender (1) | -0.003 | 0.081 | 68.406 | -0.033 | 0.973 |

*Fit statistics*

| Deviance (REML) | log Lik. | df | AIC | BIC |
|---|---|---|---|---|

*Note.* The intercept corresponds to the (unweighted) grand mean; for each factor with k levels, k - 1 parameters are estimated with sum contrast coding. Consequently, the estimates cannot be directly mapped to factor levels. Use estimated marginal means for obtaining estimates for each factor level/design cell or their differences.

#### Supplementary Table 35- RLang01 linear regression on effects of homotopic LH lesions and activity

```
jaspRegression::RegressionLinear(
        version = "0.19.2",
        collinearityStatistic = TRUE,
        covariates = ~ LLang01_lesion + LLang01 + LogLesionSize + LogTimeSinceStroke + Sem_Dec_Acc + Age +
Education + Handedness,
        dependent = ~ RLang01,
        factors = ~ Gender,
        modelTerms = list(list(components = list("LLang01_lesion", "LLang01", "LogLesionSize",
"LogTimeSinceStroke", "Sem_Dec_Acc", "Age", "Education", "Handedness", "Gender"), name = "model0", title = "Model
0")))
```

*Model Summary - RLang01*

| Model | R | $R^2$ | Adjusted $R^2$ | RMSE |
|---|---|---|---|---|
| $M_0$ | 0.534 | 0.285 | 0.188 | 0.925 |

| Model | | Sum of Squares | df | Mean Square | F | p |
|---|---|---|---|---|---|---|
| $M_0$ | Regression | 22.548 | 9 | 2.505 | 2.926 | 0.006 |
| | Residual | 56.517 | 66 | 0.856 | | |
| | Total | 79.065 | 75 | | | |

| Model | | Unstandardized | Standard Error | Standardized[a] | t | p | Collinearity Statistics | |
|---|---|---|---|---|---|---|---|---|
| | | | | | | | Tolerance | VIF |
| $M_0$ | (Intercept) | -1.317 | 2.133 | | -0.618 | 0.539 | | |
| | LLang01_lesion | 1.250 | 0.611 | 0.262 | 2.046 | 0.045 | 0.660 | 1.516 |
| | LLang01 | 0.222 | 0.134 | 0.216 | 1.657 | 0.102 | 0.640 | 1.561 |
| | LogLesionSize | -0.012 | 0.196 | -0.008 | -0.060 | 0.952 | 0.694 | 1.440 |
| | LogTimeSinceStroke | 0.165 | 0.325 | 0.063 | 0.509 | 0.612 | 0.698 | 1.432 |
| | Sem_Dec_Acc | 2.937 | 1.957 | 0.175 | 1.501 | 0.138 | 0.797 | 1.255 |
| | Age | -0.026 | 0.011 | -0.290 | -2.367 | 0.021 | 0.723 | 1.384 |
| | Education | 0.043 | 0.043 | 0.121 | 0.985 | 0.328 | 0.723 | 1.383 |
| | Handedness | -0.004 | 0.002 | -0.225 | -2.007 | 0.049 | 0.862 | 1.160 |
| | Gender (M) | -0.037 | 0.228 | | -0.161 | 0.872 | 0.880 | 1.137 |

[a] Standardized coefficients can only be computed for continuous predictors.

#### Supplementary Table 36- RLang02 linear regression on effects of homotopic LH lesions and activity

```
jaspRegression::RegressionLinear(
        version = "0.19.2",
        collinearityStatistic = TRUE,
        covariates = ~ LLang02_lesion + LLang02 + LogTimeSinceStroke + Sem_Dec_Acc + Age + Education + Handedness
+ LogLesionSize,
        dependent = ~ RLang02,
        factors = ~ Gender,
        modelTerms = list(list(components = list("LLang02_lesion", "LLang02", "LogLesionSize",
"LogTimeSinceStroke", "Sem_Dec_Acc", "Age", "Education", "Handedness", "Gender"), name = "model0", title = "Model
0")))
```

*Model Summary - RLang02*

| Model | R | $R^2$ | Adjusted $R^2$ | RMSE |
|---|---|---|---|---|
| $M_0$ | 0.549 | 0.301 | 0.206 | 0.947 |

| Model | | Sum of Squares | df | Mean Square | F | p |
|---|---|---|---|---|---|---|
| $M_0$ | Regression | 25.536 | 9 | 2.837 | 3.162 | 0.003 |
| | Residual | 59.227 | 66 | 0.897 | | |
| | Total | 84.763 | 75 | | | |

| | | | | | | | Collinearity Statistics | |
|---|---|---|---|---|---|---|---|---|
| Model | | Unstandardized | Standard Error | Standardized[a] | t | p | Tolerance | VIF |
| $M_0$ | (Intercept) | 1.969 | 2.128 | | 0.925 | 0.358 | | |
| | LLang02_lesion | 1.468 | 0.581 | 0.366 | 2.529 | 0.014 | 0.507 | 1.974 |
| | LLang02 | 0.217 | 0.171 | 0.183 | 1.268 | 0.209 | 0.510 | 1.960 |
| | LogLesionSize | -0.133 | 0.211 | -0.082 | -0.629 | 0.532 | 0.624 | 1.603 |
| | LogTimeSinceStroke | 0.202 | 0.320 | 0.075 | 0.632 | 0.530 | 0.753 | 1.328 |
| | Sem_Dec_Acc | 0.111 | 1.953 | 0.006 | 0.057 | 0.955 | 0.839 | 1.192 |
| | Age | -0.032 | 0.011 | -0.348 | -2.817 | 0.006 | 0.694 | 1.441 |
| | Education | 0.048 | 0.046 | 0.132 | 1.044 | 0.300 | 0.660 | 1.516 |
| | Handedness | -0.005 | 0.002 | -0.250 | -2.300 | 0.025 | 0.893 | 1.120 |
| | Gender (M) | -0.108 | 0.237 | | -0.455 | 0.651 | 0.855 | 1.169 |

[a] Standardized coefficients can only be computed for continuous predictors.

#### Supplementary Table 37- RLang03 linear regression on effects of homotopic LH lesions and activity

```
jaspRegression::RegressionLinear(
        version = "0.19.2",
        collinearityStatistic = TRUE,
        covariates = ~ LLang03_lesion + LLang03 + LogTimeSinceStroke + Sem_Dec_Acc + Age + Education + Handedness
+ LogLesionSize,
        dependent = ~ RLang03,
        factors = ~ Gender,
        modelTerms = list(list(components = list("LLang03_lesion", "LLang03", "LogLesionSize",
"LogTimeSinceStroke", "Sem_Dec_Acc", "Age", "Education", "Handedness", "Gender"), name = "model0", title = "Model
0")))
```

*Model Summary - RLang03*

| Model | R | $R^2$ | Adjusted $R^2$ | RMSE |
|---|---|---|---|---|
| $M_0$ | 0.458 | 0.210 | 0.102 | 0.872 |

| Model | | Sum of Squares | df | Mean Square | F | p |
|---|---|---|---|---|---|---|
| $M_0$ | Regression | 13.354 | 9 | 1.484 | 1.951 | 0.060 |
| | Residual | 50.188 | 66 | 0.760 | | |
| | Total | 63.542 | 75 | | | |

| Model | | Unstandardized | Standard Error | Standardized[a] | t | p | Collinearity Statistics | |
|---|---|---|---|---|---|---|---|---|
| | | | | | | | Tolerance | VIF |
| $M_0$ | (Intercept) | 0.189 | 2.046 | | 0.092 | 0.927 | | |
| | LLang03_lesion | 1.171 | 0.652 | 0.275 | 1.794 | 0.077 | 0.510 | 1.962 |
| | LLang03 | 0.124 | 0.156 | 0.131 | 0.799 | 0.427 | 0.445 | 2.249 |
| | LogLesionSize | -0.017 | 0.193 | -0.012 | -0.087 | 0.931 | 0.631 | 1.585 |
| | LogTimeSinceStroke | 0.191 | 0.292 | 0.082 | 0.652 | 0.516 | 0.766 | 1.306 |
| | Sem_Dec_Acc | 0.567 | 1.910 | 0.038 | 0.297 | 0.768 | 0.743 | 1.346 |
| | Age | -0.021 | 0.011 | -0.267 | -2.014 | 0.048 | 0.682 | 1.466 |
| | Education | 0.058 | 0.044 | 0.184 | 1.325 | 0.190 | 0.623 | 1.606 |
| | Handedness | -0.002 | 0.002 | -0.133 | -1.143 | 0.257 | 0.883 | 1.132 |
| | Gender (M) | -0.302 | 0.219 | | -1.379 | 0.173 | 0.850 | 1.176 |

[a] Standardized coefficients can only be computed for continuous predictors.

#### Supplementary Table 38- RLang04 linear regression on effects of homotopic LH lesions and activity

```
jaspRegression::RegressionLinear(
        version = "0.19.2",
        collinearityStatistic = TRUE,
        covariates = ~ LLang04_lesion + LLang04 + LogTimeSinceStroke + Sem_Dec_Acc + Age + Education + Handedness
+ LogLesionSize,
        dependent = ~ RLang04,
        factors = ~ Gender,
        modelTerms = list(list(components = list("LLang04_lesion", "LLang04", "LogLesionSize",
"LogTimeSinceStroke", "Sem_Dec_Acc", "Age", "Education", "Handedness", "Gender"), name = "model0", title = "Model
0"), list(components = list("LLang04_lesion", "LLang04", "LogLesionSize", "LogTimeSinceStroke", "Sem_Dec_Acc",
"Age", "Education", "Handedness", "Gender", list("LLang04_lesion", "LLang04")), name = "model1", title = "Model
1")),
        rSquaredChange = TRUE)
```

*Model Summary - RLang04*

| Model | R | $R^2$ | Adjusted $R^2$ | RMSE | $R^2$ Change | df1 | df2 | p |
|---|---|---|---|---|---|---|---|---|
| $M_0$ | 0.518 | 0.269 | 0.169 | 0.997 | 0.269 | 9 | 66 | 0.010 |
| $M_1$ | 0.523 | 0.273 | 0.161 | 1.001 | 0.004 | 1 | 65 | 0.529 |

*Note.* $M_0$ includes LLang04_lesion, LLang04, LogLesionSize, LogTimeSinceStroke, Sem_Dec_Acc, Age, Education, Handedness, Gender
*Note.* $M_1$ includes LLang04_lesion, LLang04, LogLesionSize, LogTimeSinceStroke, Sem_Dec_Acc, Age, Education, Handedness, Gender, LLang04_lesion:LLang04

*ANOVA*

| Model | | Sum of Squares | df | Mean Square | F | p |
|---|---|---|---|---|---|---|
| $M_0$ | Regression | 24.083 | 9 | 2.676 | 2.693 | 0.010 |
| | Residual | 65.590 | 66 | 0.994 | | |
| | Total | 89.673 | 75 | | | |
| $M_1$ | Regression | 24.484 | 10 | 2.448 | 2.441 | 0.015 |
| | Residual | 65.189 | 65 | 1.003 | | |
| | Total | 89.673 | 75 | | | |

*Note.* $M_0$ includes LLang04_lesion, LLang04, LogLesionSize, LogTimeSinceStroke, Sem_Dec_Acc, Age, Education, Handedness, Gender
*Note.* $M_1$ includes LLang04_lesion, LLang04, LogLesionSize, LogTimeSinceStroke, Sem_Dec_Acc, Age, Education, Handedness, Gender, LLang04_lesion:LLang04

*Coefficients*

| | | | | | | | Collinearity Statistics | |
|---|---|---|---|---|---|---|---|---|
| Model | | Unstandardized | Standard Error | Standardized[a] | t | p | Tolerance | VIF |
| $M_0$ | (Intercept) | -0.274 | 2.325 | | -0.118 | 0.907 | | |

*Coefficients*

| Model | | Unstandardized | Standard Error | Standardized[a] | t | p | Collinearity Statistics | |
|---|---|---|---|---|---|---|---|---|
| | | | | | | | Tolerance | VIF |
| | LLang04_lesion | 0.926 | 0.627 | 0.244 | 1.477 | 0.144 | 0.405 | 2.469 |
| | LLang04 | 0.192 | 0.180 | 0.167 | 1.065 | 0.291 | 0.451 | 2.219 |
| | LogLesionSize | 0.044 | 0.218 | 0.026 | 0.200 | 0.842 | 0.646 | 1.549 |
| | LogTimeSinceStroke | 0.260 | 0.332 | 0.094 | 0.785 | 0.435 | 0.777 | 1.287 |
| | Sem_Dec_Acc | 1.619 | 2.085 | 0.091 | 0.777 | 0.440 | 0.814 | 1.228 |
| | Age | -0.031 | 0.012 | -0.329 | -2.577 | 0.012 | 0.681 | 1.469 |
| | Education | 0.064 | 0.049 | 0.170 | 1.300 | 0.198 | 0.646 | 1.549 |
| | Handedness | -0.005 | 0.002 | -0.241 | -2.144 | 0.036 | 0.877 | 1.140 |
| | Gender (M) | -0.046 | 0.247 | | -0.185 | 0.854 | 0.869 | 1.151 |
| $M_1$ | (Intercept) | -0.477 | 2.357 | | -0.203 | 0.840 | | |
| | LLang04_lesion | 0.198 | 1.311 | 0.052 | 0.151 | 0.880 | 0.093 | 10.696 |
| | LLang04 | 0.232 | 0.192 | 0.202 | 1.210 | 0.231 | 0.401 | 2.494 |
| | LogLesionSize | 0.051 | 0.220 | 0.031 | 0.234 | 0.816 | 0.644 | 1.553 |
| | LogTimeSinceStroke | 0.267 | 0.333 | 0.096 | 0.802 | 0.425 | 0.776 | 1.288 |
| | Sem_Dec_Acc | 1.730 | 2.102 | 0.097 | 0.823 | 0.413 | 0.809 | 1.237 |
| | Age | -0.028 | 0.013 | -0.296 | -2.138 | 0.036 | 0.584 | 1.712 |
| | Education | 0.057 | 0.051 | 0.152 | 1.130 | 0.263 | 0.616 | 1.623 |
| | Handedness | -0.005 | 0.002 | -0.234 | -2.063 | 0.043 | 0.869 | 1.151 |
| | Gender (M) | -0.046 | 0.249 | | -0.184 | 0.855 | 0.869 | 1.151 |
| | LLang04_lesion ✻ LLang04 | -0.478 | 0.755 | -0.232 | -0.633 | 0.529 | 0.083 | 11.983 |

[a] Standardized coefficients can only be computed for continuous predictors.

#### Supplementary Table 39- RLang05 linear regression on effects of homotopic LH lesions and activity

```
jaspRegression::RegressionLinear(
        version = "0.19.2",
        collinearityStatistic = TRUE,
        covariates = ~ LLang05_lesion + LLang05 + LogTimeSinceStroke + Sem_Dec_Acc + Age + Education + Handedness
+ LogLesionSize,
        dependent = ~ RLang05,
        factors = ~ Gender,
        modelTerms = list(list(components = list("LLang05_lesion", "LLang05", "LogLesionSize",
"LogTimeSinceStroke", "Sem_Dec_Acc", "Age", "Education", "Handedness", "Gender"), name = "model0", title = "Model
0"), list(components = list("LLang05_lesion", "LLang05", "LogLesionSize", "LogTimeSinceStroke", "Sem_Dec_Acc",
"Age", "Education", "Handedness", "Gender", list("LLang05_lesion", "LLang05")), name = "model1", title = "Model
1")),
        rSquaredChange = TRUE)
```

*Model Summary - RLang05*

| Model | R | $R^2$ | Adjusted $R^2$ | RMSE | $R^2$ Change | df1 | df2 | p |
|---|---|---|---|---|---|---|---|---|
| $M_0$ | 0.502 | 0.252 | 0.150 | 0.870 | 0.252 | 9 | 66 | 0.017 |
| $M_1$ | 0.565 | 0.319 | 0.215 | 0.836 | 0.067 | 1 | 65 | 0.014 |

*Note.* $M_0$ includes LLang05_lesion, LLang05, LogLesionSize, LogTimeSinceStroke, Sem_Dec_Acc, Age, Education, Handedness, Gender
*Note.* $M_1$ includes LLang05_lesion, LLang05, LogLesionSize, LogTimeSinceStroke, Sem_Dec_Acc, Age, Education, Handedness, Gender, LLang05_lesion:LLang05

*ANOVA*

| Model | | Sum of Squares | df | Mean Square | F | p |
|---|---|---|---|---|---|---|
| $M_0$ | Regression | 16.820 | 9 | 1.869 | 2.471 | 0.017 |
| | Residual | 49.921 | 66 | 0.756 | | |
| | Total | 66.741 | 75 | | | |
| $M_1$ | Regression | 21.311 | 10 | 2.131 | 3.049 | 0.003 |
| | Residual | 45.430 | 65 | 0.699 | | |
| | Total | 66.741 | 75 | | | |

*Note.* $M_0$ includes LLang05_lesion, LLang05, LogLesionSize, LogTimeSinceStroke, Sem_Dec_Acc, Age, Education, Handedness, Gender
*Note.* $M_1$ includes LLang05_lesion, LLang05, LogLesionSize, LogTimeSinceStroke, Sem_Dec_Acc, Age, Education, Handedness, Gender, LLang05_lesion:LLang05

*Coefficients*

| | | | | | | | Collinearity Statistics | |
|---|---|---|---|---|---|---|---|---|
| Model | | Unstandardized | Standard Error | Standardized[a] | t | p | Tolerance | VIF |
| $M_0$ | (Intercept) | 1.378 | 1.962 | | 0.702 | 0.485 | | |

*Coefficients*

| Model | | Unstandardized | Standard Error | Standardized[a] | t | p | Collinearity Statistics | |
|---|---|---|---|---|---|---|---|---|
| | | | | | | | Tolerance | VIF |
| | LLang05_lesion | 0.130 | 0.430 | 0.055 | 0.301 | 0.764 | 0.345 | 2.901 |
| | LLang05 | 0.050 | 0.173 | 0.051 | 0.290 | 0.773 | 0.367 | 2.723 |
| | LogLesionSize | 0.002 | 0.199 | 0.002 | 0.011 | 0.991 | 0.593 | 1.686 |
| | LogTimeSinceStroke | 0.549 | 0.293 | 0.229 | 1.875 | 0.065 | 0.759 | 1.317 |
| | Sem_Dec_Acc | -0.800 | 1.794 | -0.052 | -0.446 | 0.657 | 0.837 | 1.195 |
| | Age | -0.026 | 0.010 | -0.319 | -2.533 | 0.014 | 0.715 | 1.398 |
| | Education | 0.064 | 0.041 | 0.198 | 1.568 | 0.122 | 0.710 | 1.407 |
| | Handedness | -0.006 | 0.002 | -0.326 | -2.868 | 0.006 | 0.875 | 1.142 |
| | Gender (M) | -0.147 | 0.216 | | -0.680 | 0.499 | 0.865 | 1.156 |
| $M_1$ | (Intercept) | 0.077 | 1.955 | | 0.040 | 0.969 | | |
| | LLang05_lesion | -1.768 | 0.855 | -0.746 | -2.068 | 0.043 | 0.080 | 12.423 |
| | LLang05 | 0.225 | 0.180 | 0.229 | 1.252 | 0.215 | 0.313 | 3.193 |
| | LogLesionSize | 0.096 | 0.195 | 0.067 | 0.494 | 0.623 | 0.572 | 1.750 |
| | LogTimeSinceStroke | 0.572 | 0.281 | 0.239 | 2.032 | 0.046 | 0.759 | 1.318 |
| | Sem_Dec_Acc | -0.066 | 1.749 | -0.004 | -0.038 | 0.970 | 0.814 | 1.228 |
| | Age | -0.021 | 0.010 | -0.252 | -2.036 | 0.046 | 0.683 | 1.464 |
| | Education | 0.061 | 0.039 | 0.187 | 1.535 | 0.130 | 0.709 | 1.409 |
| | Handedness | -0.005 | 0.002 | -0.273 | -2.456 | 0.017 | 0.845 | 1.184 |
| | Gender (M) | -0.103 | 0.209 | | -0.496 | 0.622 | 0.859 | 1.163 |
| | LLang05_lesion ✻ LLang05 | -1.189 | 0.469 | -0.951 | -2.535 | 0.014 | 0.074 | 13.427 |

[a] Standardized coefficients can only be computed for continuous predictors.

#### Supplementary Table 40- RLang06 linear regression on effects of homotopic LH lesions and activity

```
jaspRegression::RegressionLinear(
        version = "0.19.2",
        collinearityStatistic = TRUE,
        covariates = ~ LLang06_lesion + LLang06 + LogTimeSinceStroke + Sem_Dec_Acc + Age + Education + Handedness
+ LogLesionSize,
        dependent = ~ RLang06,
        factors = ~ Gender,
        modelTerms = list(list(components = list("LLang06_lesion", "LLang06", "LogLesionSize",
"LogTimeSinceStroke", "Sem_Dec_Acc", "Age", "Education", "Handedness", "Gender"), name = "model0", title = "Model
0")))
```

*Model Summary - RLang06*

| Model | R | $R^2$ | Adjusted $R^2$ | RMSE |
|---|---|---|---|---|
| $M_0$ | 0.623 | 0.388 | 0.305 | 0.995 |

| Model | | Sum of Squares | df | Mean Square | F | p |
|---|---|---|---|---|---|---|
| $M_0$ | Regression | 41.513 | 9 | 4.613 | 4.658 | $8.642 \times 10^{-5}$ |
| | Residual | 65.362 | 66 | 0.990 | | |
| | Total | 106.875 | 75 | | | |

| | | | | | | | Collinearity Statistics | |
|---|---|---|---|---|---|---|---|---|
| Model | | Unstandardized | Standard Error | Standardized[a] | t | p | Tolerance | VIF |
| $M_0$ | (Intercept) | -4.034 | 2.382 | | -1.694 | 0.095 | | |
| | LLang06_lesion | 1.162 | 0.446 | 0.279 | 2.604 | 0.011 | 0.810 | 1.235 |
| | LLang06 | 0.333 | 0.130 | 0.300 | 2.555 | 0.013 | 0.671 | 1.489 |
| | LogLesionSize | 0.101 | 0.205 | 0.056 | 0.495 | 0.622 | 0.734 | 1.362 |
| | LogTimeSinceStroke | 0.802 | 0.324 | 0.265 | 2.475 | 0.016 | 0.810 | 1.235 |
| | Sem_Dec_Acc | 2.427 | 2.164 | 0.124 | 1.122 | 0.266 | 0.753 | 1.327 |
| | Age | -0.006 | 0.012 | -0.055 | -0.487 | 0.628 | 0.733 | 1.364 |
| | Education | 0.050 | 0.047 | 0.123 | 1.072 | 0.288 | 0.709 | 1.411 |
| | Handedness | -0.004 | 0.002 | -0.157 | -1.532 | 0.130 | 0.882 | 1.133 |
| | Gender (M) | 0.102 | 0.252 | | 0.404 | 0.688 | 0.834 | 1.199 |

[a] Standardized coefficients can only be computed for continuous predictors.

#### Supplementary Table 41- RLang07 linear regression on effects of homotopic LH lesions and activity

```
jaspRegression::RegressionLinear(
        version = "0.19.2",
        collinearityStatistic = TRUE,
        covariates = ~ LLang07_lesion + LLang07 + LogTimeSinceStroke + Sem_Dec_Acc + Age + Education + Handedness
+ LogLesionSize,
        dependent = ~ RLang07,
        factors = ~ Gender,
        modelTerms = list(list(components = list("LLang07_lesion", "LLang07", "LogLesionSize",
"LogTimeSinceStroke", "Sem_Dec_Acc", "Age", "Education", "Handedness", "Gender"), name = "model0", title = "Model
0")))
```

*Model Summary - RLang07*

| Model | R | $R^2$ | Adjusted $R^2$ | RMSE |
|---|---|---|---|---|
| $M_0$ | 0.637 | 0.406 | 0.325 | 0.599 |

| Model | | Sum of Squares | df | Mean Square | F | p |
|---|---|---|---|---|---|---|
| $M_0$ | Regression | 16.153 | 9 | 1.795 | 5.009 | $3.842 \times 10^{-5}$ |
| | Residual | 23.650 | 66 | 0.358 | | |
| | Total | 39.803 | 75 | | | |

| | | | | | | | Collinearity Statistics | |
|---|---|---|---|---|---|---|---|---|
| Model | | Unstandardized | Standard Error | Standardized[a] | t | p | Tolerance | VIF |
| $M_0$ | (Intercept) | -1.574 | 1.370 | | -1.149 | 0.255 | | |
| | LLang07_lesion | 0.760 | 0.265 | 0.309 | 2.866 | 0.006 | 0.773 | 1.294 |
| | LLang07 | 0.247 | 0.107 | 0.261 | 2.316 | 0.024 | 0.711 | 1.406 |
| | LogLesionSize | -0.273 | 0.125 | -0.245 | -2.174 | 0.033 | 0.706 | 1.416 |
| | LogTimeSinceStroke | 0.262 | 0.189 | 0.141 | 1.388 | 0.170 | 0.867 | 1.153 |
| | Sem_Dec_Acc | 2.164 | 1.257 | 0.182 | 1.721 | 0.090 | 0.808 | 1.238 |
| | Age | -0.002 | 0.007 | -0.034 | -0.310 | 0.758 | 0.738 | 1.356 |
| | Education | 0.045 | 0.028 | 0.181 | 1.625 | 0.109 | 0.728 | 1.374 |
| | Handedness | -0.003 | 0.001 | -0.221 | -2.198 | 0.031 | 0.894 | 1.119 |
| | Gender (M) | 0.180 | 0.154 | | 1.172 | 0.245 | 0.812 | 1.232 |

[a] Standardized coefficients can only be computed for continuous predictors.

#### Supplementary Table 42- RLang08 linear regression on effects of homotopic LH lesions and activity

```
jaspRegression::RegressionLinear(
        version = "0.19.2",
        collinearityStatistic = TRUE,
        covariates = ~ LLang08_lesion + LLang08 + LogTimeSinceStroke + Sem_Dec_Acc + Age + Education + Handedness
+ LogLesionSize,
        dependent = ~ RLang08,
        factors = ~ Gender,
        modelTerms = list(list(components = list("LLang08_lesion", "LLang08", "LogLesionSize",
"LogTimeSinceStroke", "Sem_Dec_Acc", "Age", "Education", "Handedness", "Gender"), name = "model0", title = "Model
0")))
```

*Model Summary - RLang08*

| Model | R | $R^2$ | Adjusted $R^2$ | RMSE |
|---|---|---|---|---|
| $M_0$ | 0.566 | 0.321 | 0.228 | 0.918 |

| Model | | Sum of Squares | df | Mean Square | F | p |
|---|---|---|---|---|---|---|
| $M_0$ | Regression | 26.248 | 9 | 2.916 | 3.461 | 0.002 |
| | Residual | 55.611 | 66 | 0.843 | | |
| | Total | 81.859 | 75 | | | |

| | | | | | | | Collinearity Statistics | |
|---|---|---|---|---|---|---|---|---|
| Model | | Unstandardized | Standard Error | Standardized[a] | t | p | Tolerance | VIF |
| $M_0$ | (Intercept) | -1.208 | 2.138 | | -0.565 | 0.574 | | |
| | LLang08_lesion | 1.490 | 0.397 | 0.443 | 3.748 | $3.776\times10^{-4}$ | 0.738 | 1.355 |
| | LLang08 | 0.550 | 0.166 | 0.468 | 3.311 | 0.002 | 0.516 | 1.939 |
| | LogLesionSize | 0.040 | 0.201 | 0.025 | 0.197 | 0.844 | 0.649 | 1.540 |
| | LogTimeSinceStroke | -0.104 | 0.306 | -0.039 | -0.340 | 0.735 | 0.772 | 1.295 |
| | Sem_Dec_Acc | 1.620 | 2.013 | 0.095 | 0.805 | 0.424 | 0.741 | 1.350 |
| | Age | -0.002 | 0.011 | -0.024 | -0.203 | 0.840 | 0.730 | 1.370 |
| | Education | -0.011 | 0.044 | -0.031 | -0.251 | 0.803 | 0.680 | 1.470 |
| | Handedness | $-7.596\times10^{-4}$ | 0.002 | -0.039 | -0.357 | 0.722 | 0.874 | 1.144 |
| | Gender (M) | 0.091 | 0.229 | | 0.396 | 0.693 | 0.858 | 1.166 |

[a] Standardized coefficients can only be computed for continuous predictors.

#### Supplementary Table 43- RLang09 linear regression on effects of homotopic LH lesions and activity

```
jaspRegression::RegressionLinear(
        version = "0.19.2",
        collinearityStatistic = TRUE,
        covariates = ~ LLang09_lesion + LLang09 + LogTimeSinceStroke + Sem_Dec_Acc + Age + Education + Handedness
+ LogLesionSize,
        dependent = ~ RLang09,
        factors = ~ Gender,
        modelTerms = list(list(components = list("LLang09_lesion", "LLang09", "LogLesionSize",
"LogTimeSinceStroke", "Sem_Dec_Acc", "Age", "Education", "Handedness", "Gender"), name = "model0", title = "Model
0")))
```

*Model Summary - RLang09*

| Model | R | $R^2$ | Adjusted $R^2$ | RMSE |
|---|---|---|---|---|
| $M_0$ | 0.461 | 0.212 | 0.105 | 1.089 |

| Model | | Sum of Squares | df | Mean Square | F | p |
|---|---|---|---|---|---|---|
| $M_0$ | Regression | 21.073 | 9 | 2.341 | 1.975 | 0.056 |
| | Residual | 78.251 | 66 | 1.186 | | |
| | Total | 99.324 | 75 | | | |

| | | | | | | | Collinearity Statistics | |
|---|---|---|---|---|---|---|---|---|
| Model | | Unstandardized | Standard Error | Standardized[a] | t | p | Tolerance | VIF |
| $M_0$ | (Intercept) | 2.842 | 2.345 | | 1.212 | 0.230 | | |
| | LLang09_lesion | 0.855 | 0.434 | 0.277 | 1.971 | 0.053 | 0.604 | 1.657 |
| | LLang09 | 0.440 | 0.190 | 0.339 | 2.311 | 0.024 | 0.556 | 1.798 |
| | LogLesionSize | -0.157 | 0.238 | -0.089 | -0.660 | 0.512 | 0.649 | 1.540 |
| | LogTimeSinceStroke | 0.481 | 0.343 | 0.165 | 1.403 | 0.165 | 0.868 | 1.152 |
| | Sem_Dec_Acc | -2.195 | 2.297 | -0.117 | -0.956 | 0.343 | 0.801 | 1.249 |
| | Age | -0.023 | 0.013 | -0.237 | -1.811 | 0.075 | 0.699 | 1.431 |
| | Education | 0.040 | 0.050 | 0.102 | 0.800 | 0.427 | 0.739 | 1.352 |
| | Handedness | -0.003 | 0.003 | -0.148 | -1.273 | 0.208 | 0.880 | 1.137 |
| | Gender (M) | 0.069 | 0.271 | | 0.253 | 0.801 | 0.862 | 1.160 |

[a] Standardized coefficients can only be computed for continuous predictors.

#### Supplementary Table 44- RLang10 linear regression on effects of homotopic LH lesions and activity

```
jaspRegression::RegressionLinear(
        version = "0.19.2",
        collinearityStatistic = TRUE,
        covariates = ~ LLang10_lesion + LLang10 + LogTimeSinceStroke + Sem_Dec_Acc + Age + Education + Handedness
+ LogLesionSize,
        dependent = ~ RLang10,
        factors = ~ Gender,
        modelTerms = list(list(components = list("LLang10_lesion", "LLang10", "LogLesionSize",
"LogTimeSinceStroke", "Sem_Dec_Acc", "Age", "Education", "Handedness", "Gender"), name = "model0", title = "Model
0")))
```

*Model Summary - RLang10*

| Model | R | $R^2$ | Adjusted $R^2$ | RMSE |
|---|---|---|---|---|
| $M_0$ | 0.402 | 0.162 | 0.047 | 1.204 |

| Model | | Sum of Squares | df | Mean Square | F | p |
|---|---|---|---|---|---|---|
| $M_0$ | Regression | 18.446 | 9 | 2.050 | 1.414 | 0.200 |
| | Residual | 95.683 | 66 | 1.450 | | |
| | Total | 114.128 | 75 | | | |

*Coefficients*

| | | | | | | | Collinearity Statistics | |
|---|---|---|---|---|---|---|---|---|
| Model | | Unstandardized | Standard Error | Standardized[a] | t | p | Tolerance | VIF |
| $M_0$ | (Intercept) | 0.500 | 2.593 | | 0.193 | 0.848 | | |
| | LLang10_lesion | 0.674 | 0.523 | 0.180 | 1.290 | 0.202 | 0.656 | 1.525 |
| | LLang10 | 0.309 | 0.201 | 0.205 | 1.540 | 0.128 | 0.717 | 1.395 |
| | LogLesionSize | -0.370 | 0.257 | -0.197 | -1.444 | 0.153 | 0.683 | 1.464 |
| | LogTimeSinceStroke | 0.177 | 0.381 | 0.056 | 0.464 | 0.644 | 0.857 | 1.166 |
| | Sem_Dec_Acc | 1.455 | 2.428 | 0.072 | 0.599 | 0.551 | 0.876 | 1.141 |
| | Age | -0.017 | 0.014 | -0.156 | -1.194 | 0.237 | 0.741 | 1.350 |
| | Education | 0.071 | 0.056 | 0.168 | 1.277 | 0.206 | 0.735 | 1.360 |
| | Handedness | -0.004 | 0.003 | -0.167 | -1.372 | 0.175 | 0.856 | 1.168 |
| | Gender (M) | 0.078 | 0.299 | | 0.260 | 0.796 | 0.869 | 1.151 |

[a] Standardized coefficients can only be computed for continuous predictors.

#### Supplementary Table 45- RLang11 linear regression on effects of homotopic LH lesions and activity

```
jaspRegression::RegressionLinear(
        version = "0.19.2",
        collinearityStatistic = TRUE,
        covariates = ~ LLang11_lesion + LLang11 + LogTimeSinceStroke + Sem_Dec_Acc + Age + Education + Handedness
+ LogLesionSize,
        dependent = ~ RLang11,
        factors = ~ Gender,
        modelTerms = list(list(components = list("LLang11_lesion", "LLang11", "LogLesionSize",
"LogTimeSinceStroke", "Sem_Dec_Acc", "Age", "Education", "Handedness", "Gender"), name = "model0", title = "Model
0")))
```

*Model Summary - RLang11*

| Model | R | $R^2$ | Adjusted $R^2$ | RMSE |
|---|---|---|---|---|
| $M_0$ | 0.381 | 0.145 | 0.028 | 0.681 |

| Model | | Sum of Squares | df | Mean Square | F | p |
|---|---|---|---|---|---|---|
| $M_0$ | Regression | 5.181 | 9 | 0.576 | 1.242 | 0.286 |
| | Residual | 30.603 | 66 | 0.464 | | |
| | Total | 35.784 | 75 | | | |

*Coefficients*

| | | | | | | | Collinearity Statistics | |
|---|---|---|---|---|---|---|---|---|
| Model | | Unstandardized | Standard Error | Standardized[a] | t | p | Tolerance | VIF |
| $M_0$ | (Intercept) | 1.978 | 1.452 | | 1.362 | 0.178 | | |
| | LLang11_lesion | -0.311 | 0.271 | -0.148 | -1.146 | 0.256 | 0.780 | 1.283 |
| | LLang11 | -0.081 | 0.114 | -0.092 | -0.716 | 0.477 | 0.792 | 1.263 |
| | LogLesionSize | -0.013 | 0.143 | -0.013 | -0.093 | 0.926 | 0.703 | 1.422 |
| | LogTimeSinceStroke | 0.040 | 0.215 | 0.023 | 0.186 | 0.853 | 0.864 | 1.158 |
| | Sem_Dec_Acc | -1.298 | 1.360 | -0.115 | -0.954 | 0.343 | 0.893 | 1.120 |
| | Age | -0.006 | 0.008 | -0.097 | -0.730 | 0.468 | 0.741 | 1.349 |
| | Education | 0.006 | 0.031 | 0.026 | 0.194 | 0.847 | 0.737 | 1.357 |
| | Handedness | -0.005 | 0.002 | -0.371 | -2.929 | 0.005 | 0.810 | 1.235 |
| | Gender (M) | -0.130 | 0.169 | | -0.772 | 0.443 | 0.873 | 1.146 |

[a] Standardized coefficients can only be computed for continuous predictors.

### Supplementary Table 46- Mixed-effects model examining effects of nodewise homotopic LH activity on RH Language Network activity in the Control group

```
jaspMixedModels::MixedModelsLMM(
        version = "0.19.2",
        formula = RHActivity ~ semantic_acc + Handedness + Gender + Education + Age + 
LHActivity + (1 | Node) + (1 | Participant),
        contrasts = NULL,
        fixedEffectEstimate = TRUE,
        modelSummary = TRUE,
        plotBackgroundData = ~ Node + Participant,
        trendsContrasts = NULL)
```

*ANOVA Summary*

| Effect | df | F | p |
|---|---|---|---|
| semantic_acc | 1, 44.12 | 1.086 | 0.303 |
| Handedness | 1, 43.07 | 4.958 | 0.031 |
| Gender | 1, 42.63 | 0.038 | 0.846 |
| Education | 1, 43.10 | 5.243 | 0.027 |
| Age | 1, 42.50 | 0.828 | 0.368 |
| LHActivity | 1, 479.67 | 21.132 | $5.490\times10^{-6}$ |

*Note.* Model terms tested with Satterthwaite testMethod.
*Note.* The following variables are used as random effects grouping factors: 'Node', 'Participant'.
*Note.* Type III Sum of Squares

#### Model summary

*Fit statistics*

| Deviance (REML) | log Lik. | df | AIC | BIC |
|---|---|---|---|---|
| 2123.562 | -1061.781 | 10 | 2143.562 | 2186.459 |

*Note.* The model was fitted using restricted maximum likelihood. Please note that models with different fixed effects cannot be compared when REML is used. To use ML, switch 'Test method' to 'Likelihood ratio tests'.

*Sample sizes*

| | Levels of RE grouping factors | |
|---|---|---|
| Observations | Participant | Node |
| 539 | 49 | 11 |

*Fixed Effects Estimates*

| Term | Estimate | SE | df | t | p |
|---|---|---|---|---|---|
| Intercept | 2.484 | 3.166 | 44.689 | 0.784 | 0.437 |
| semantic_acc | -3.941 | 3.782 | 44.118 | -1.042 | 0.303 |
| Handedness | -0.007 | 0.003 | 43.071 | -2.227 | 0.031 |
| Gender (1) | -0.026 | 0.131 | 42.635 | -0.196 | 0.846 |
| Education | 0.116 | 0.051 | 43.096 | 2.290 | 0.027 |
| Age | -0.011 | 0.012 | 42.499 | -0.910 | 0.368 |
| LHActivity | 0.135 | 0.029 | 479.670 | 4.597 | $5.490\times10^{-6}$ |

*Note.* The intercept corresponds to the (unweighted) grand mean; for each factor with k levels, k - 1 parameters are estimated with sum contrast coding. Consequently, the estimates cannot be directly mapped to factor levels. Use estimated marginal means for obtaining estimates for each factor level/design cell or their differences.

#### Supplementary Table 47- Significant lesion-symptom mapping results

| Node | Location of LSM result | Cluster p-value | Volume (mm3) | MNI Center of Mass x | y | z | MNI Bounding Box Min x | Min y | Min z | Max x | Max y | Max z |
|---|---|---|---|---|---|---|---|---|---|---|---|---|
| RLang01 | IFG pars triangulars, pars orbitalis, anterior insula, putamen | 0.009 | 11031 | -39.6 | 22.9 | -3.8 | -25 | -10 | -21 | -55 | 43 | 12 |
| RLang02 | Anterior insula, IFG pars orbitalis | 0.0436 | 4391 | -35.4 | 22.3 | -3.5 | -25 | 5 | -16 | -53 | 38 | 12 |
| RLang03 | IFG pars triangulars, pars orbitalis, pars opercularis, posterior IFS, anterior insula | 0.0134 | 9250 | -39 | 21 | 4 | -23 | -3 | -16 | -58 | 43 | 34 |
| RLang06 | Anterior STG/STS/MTG | 0.045 | 4594 | -44 | 9.9 | -30.3 | -30 | -10 | -44 | -53 | 23 | -9 |
| RLang07 | Anterior and mid STG/STS/MTG, insula, putamen | 0.0002 | 24547 | -47.9 | -5.5 | -14.7 | -15 | -45 | -44 | -68 | 25 | 22 |
| RLang08 | Anterior and mid STG/STS/MTG | 0.0014 | 19922 | -52.6 | -6 | -16.3 | -33 | -45 | -44 | -68 | 23 | 4 |
| RLang09 | IFG pars triangulars, pars orbitalis, anterior MFG, anterior insula | 0.0204 | 7266 | -43.4 | 30.6 | -1.7 | -25 | 10 | -16 | -55 | 48 | 14 |
| RLang10 | IFG pars triangulars, pars orbitalis, anterior insula | 0.0462 | 4297 | -42.7 | 27.9 | -2.8 | -28 | 10 | -14 | -53 | 45 | 9 |

**Supplementary Table 48- Partial Spearman correlations of behavioral scores with RH language node activity**

Partial correlations control for age, education, handedness, gender, log-time-since-stroke, log-lesion size, and lesion load in LH nodes with adequate lesion coverage (all 11 LLang nodes, LSem04, LSem05, LSem12)

P<.05 corrected for 66 comparison (11 nodes x 6 behaviors)
P<.05 corrected for 11 comparisons
Uncorrected P < .01
Uncorrected P < .05

| | Western Aphasia Battery | | | | | |
|---|---|---|---|---|---|---|
| Node | Spontaneous Speech | Naming & Word Finding | Repetition | Auditory Verbal Comprehension | Oral Word Reading | Semantic Judgement Composite |
| RLang01 | 0.228 | 0.43 | 0.103 | 0.214 | 0.364 | 0.369 |
| RLang02 | 0.097 | 0.287 | 0.005 | 0.175 | 0.255 | 0.088 |
| RLang03 | -0.037 | 0.097 | 0.01 | -0.167 | 0.251 | 0.006 |
| RLang04 | 0.065 | 0.107 | -0.056 | 0.069 | 0.112 | 0.099 |
| RLang05 | -0.068 | 0.148 | -0.138 | 0.134 | 0.031 | 0.089 |
| RLang06 | 0.075 | 0.254 | 0.045 | 0.106 | 0.194 | 0.115 |
| RLang07 | 0.096 | 0.318 | 0.009 | 0.297 | 0.318 | 0.205 |
| RLang08 | 0.19 | 0.366 | 0.221 | 0.125 | 0.448 | 0.134 |
| RLang09 | -0.09 | 0.252 | 0.024 | -0.051 | 0.279 | -0.011 |
| RLang10 | -0.021 | 0.28 | 0.02 | -0.015 | 0.377 | 0.131 |
| RLang11 | -0.123 | -0.095 | -0.034 | -0.083 | -0.026 | -0.181 |

**Supplementary Table 49- Partial Spearman correlations of behavioral scores with LH language and semantic node activity**

Partial correlations control for age, education, handedness, gender, log-time-since-stroke, log-lesion size, and lesion load in LH nodes with adequate lesion coverage (all 11 LLang nodes, LSem04, LSem05, LSem12)
Corrected significant thresholds are the same as those used for RLang nodes to facilitate comparison.

P<.05 corrected for 66 comparison (11 nodes x 6 behaviors)

P<.05 corrected for 11 comparisons

Uncorrected P < .01

Uncorrected P < .05

| | Western Aphasia Battery | | | | | |
|---|---|---|---|---|---|---|
| | Spontaneous Speech | Naming & Word Finding | Repetition | Auditory Verbal Comprehension | Oral Reading | Semantic Judgement Composite |
| LLang01 | 0.138 | 0.461 | -0.021 | 0.268 | 0.351 | 0.349 |
| LLang02 | 0.095 | 0.276 | -0.106 | 0.206 | 0.123 | 0.221 |
| LLang03 | 0.209 | 0.439 | 0.094 | 0.274 | 0.305 | 0.464 |
| LLang04 | 0.261 | 0.299 | 0.07 | 0.212 | 0.109 | 0.398 |
| LLang05 | 0.13 | 0.262 | 0.067 | 0.229 | 0.065 | 0.169 |
| LLang06 | 0.051 | 0.15 | -0.025 | 0.064 | 0.04 | 0.162 |
| LLang07 | 0.096 | 0.383 | 0.104 | 0.262 | 0.273 | 0.19 |
| LLang08 | 0.228 | 0.438 | 0.099 | 0.293 | 0.284 | 0.26 |
| LLang09 | 0.108 | 0.419 | 0.14 | 0.097 | 0.315 | 0.15 |
| LLang10 | -0.014 | 0.22 | -0.02 | -0.052 | 0.222 | 0.023 |
| LLang11 | -0.075 | -0.001 | 0.064 | -0.047 | -0.029 | -0.133 |
| LSem01 | 0.044 | 0.269 | -0.123 | 0.096 | 0.21 | 0.34 |
| LSem02 | 0.315 | 0.389 | 0.194 | 0.335 | 0.292 | 0.243 |
| LSem03 | 0.201 | 0.282 | 0.118 | 0.224 | 0.316 | 0.296 |
| LSem04 | 0.03 | 0.116 | -0.083 | -0.109 | -0.065 | -0.035 |
| LSem05 | 0.066 | 0.256 | 0.14 | -0.002 | 0.191 | -0.019 |
| LSem06 | 0.085 | 0.299 | 0.085 | 0.184 | 0.409 | 0.299 |
| LSem07 | 0.057 | 0.32 | 0.011 | 0.176 | 0.395 | 0.338 |
| LSem08 | 0.03 | 0.147 | -0.101 | 0.105 | 0.28 | 0.334 |
| LSem09 | 0.054 | 0.343 | -0.087 | 0.224 | 0.124 | 0.318 |
| LSem10 | 0.163 | 0.393 | 0.154 | 0.287 | 0.279 | 0.126 |
| LSem11 | 0.128 | 0.322 | 0.024 | 0.27 | 0.279 | 0.321 |
| LSem12 | -0.032 | 0.04 | 0.306 | -0.047 | 0.185 | -0.131 |
| LSem13 | -0.08 | 0.152 | -0.09 | 0.021 | 0.09 | 0.117 |
| LSem14 | -0.049 | 0.059 | -0.038 | -0.108 | 0.126 | 0.057 |
| RSem01 | 7.44E-04 | 0.108 | -0.048 | 0.14 | 0.332 | 0.366 |
| RSem02 | 0.262 | 0.396 | 0.09 | 0.305 | 0.305 | 0.255 |

| | | | | | | |
|---|---|---|---|---|---|---|
| RSem03 | 0.147 | 0.301 | 0.074 | 0.148 | 0.257 | 0.252 |
| RSem04 | -0.11 | -0.15 | -0.031 | -0.06 | -0.177 | -0.284 |
| RSem05 | -0.092 | 0.112 | -0.114 | -0.066 | -0.12 | -0.019 |
| RSem06 | 0.13 | 0.312 | 0.148 | 0.019 | 0.343 | 0.168 |
| RSem07 | 0.122 | 0.309 | 0.046 | 0.137 | 0.266 | 0.145 |
| RSem08 | -0.001 | 0.09 | 0.065 | 0.05 | 0.137 | 0.073 |
| RSem09 | 0.086 | 0.199 | 0.125 | 0.143 | 0.076 | 0.144 |
| RSem10 | 0.027 | 0.096 | 0.178 | 0.09 | 0.129 | 0.043 |
| RSem11 | 0.186 | 0.332 | 0.054 | 0.281 | 0.187 | 0.191 |
| RSem12 | -0.021 | 0.041 | 0.034 | 0.074 | 0.01 | -0.081 |
| RSem13 | -0.27 | 0.077 | -0.185 | -0.087 | 0.066 | 0.009 |
| RSem14 | -0.061 | 0.086 | -0.026 | -0.113 | 0.192 | 0.043 |
